# Effect of the Friendship Bench intervention on antiretroviral therapy outcomes and mental health symptoms in rural Zimbabwe: A cluster randomized trial

**DOI:** 10.1101/2023.01.21.23284784

**Authors:** Andreas D Haas, Cordelia Kunzekwenyika, Josphat Manzero, Stefanie Hossmann, Andreas Limacher, Janneke H van Dijk, Ronald Manhibi, Per von Groote, Michael A Hobbins, Ruth Verhey, Matthias Egger, the Friendship Bench ART trial group

## Abstract

**Importance:** Common mental disorders (CMD) are prevalent in people living with HIV and associated with suboptimal antiretroviral therapy (ART) adherence.

**Objective:** To assess the effect of a lay health worker-led psychological intervention on adherence to ART, virologic suppression and mental health symptoms.

**Design:** Pragmatic cluster trial with block randomization of health facilities. Treatment assignment was known to participants, providers, evaluators, and data analysts. Recruitment started in October 2018 and the last follow-up visit was done in December 2020. Participants were followed up for 12 months.

**Setting:** Sixteen public health care facilities in Bikita, a rural district in Masvingo Province, about 300 km south of Harare.

**Participants:** Men and non-pregnant women aged 18 years or older who spoke English or Shona, screened positive for CMD (Shona Symptoms Questionnaire [SSQ]-14 score ≥9), had received first-line ART for at least six months, had no WHO clinical stage 4 disease, no psychotic symptoms, and gave informed consent.

**Intervention:** The Friendship Bench, a lay health worker-led intervention consisting of six weekly individual counselling sessions of problem-solving therapy and optional peer-led group support.

**Main Outcomes and Measures:** The primary outcome was Medication Event Monitoring System (MEMS) mean adherence between 2-6 months of follow-up. Secondary outcomes included mean adherence between 1-12 months, change from baseline SSQ-14 and Patient Health Questionnaire (PHQ-9) score at 3, 6, 9, and 12 months and change in viral load suppression (viral load <1000 copies per mL) at months 6 and 12.

**Results:** We recruited 516 participants, 244 in Friendship bench and 272 in standard care facilities. The mean age was 45.6 years (SD 10.9), and most participants were women (84.9%). In the Friendship Bench group, 88.1% of participants attended all six individual counselling sessions. Rates of adherence (>85%) and virologic suppression (>90%) were high in both groups. The intervention had no statistically significant effect on adherence or viral suppression. Declines in SSQ-14 scores from baseline to 3 months (-1.65, 95% CI -3.07 to -0.24), 6 months (-1.57, 95% CI -2.98 to -0.15), and 9 months (-1.63, 95% CI -3.05 to -0.22) were greater in the Friendship Bench than the standard care group (p<0.05). There were no differences in the decline in the SSQ-14 scores from baseline to 12 months and in declines in PHQ-9 scores from baseline to 3, 6, 9, and 12 months.

**Conclusions and Relevance:** The Friendship Bench intervention is a feasible and acceptable approach to closing the treatment gap in mental health care in rural Zimbabwe. The intervention improved CMD symptoms but the intervention effect was smaller than previously shown in an urban setting. The intervention had no effect on adherence and viral suppression, possibly due to the absence of skill-based adherence training and ceiling effect.

**Trial registration:** ClinicalTrials.gov Identifier: NCT03704805

**Key points:** *Question:*

- Does the Friendship Bench intervention improve antiretroviral therapy (ART) adherence, viral suppression and mental health symptoms in people living with HIV in rural Zimbabwe?

*Findings:*

- In this cluster-randomized trial, participants in the intervention group had a significantly greater decrease in symptoms of common mental disorders than those in the control group, but the intervention showed no significant effect on antiretroviral therapy (ART) adherence or viral suppression.

*Meaning:*

- The intervention did not affect adherence and viral suppression and the effect of the intervention on mental health symptoms was smaller than previously shown.

## Introduction

Zimbabwe carries a high HIV burden, with an estimated 1.2 million adults living with HIV in 2020.^1^ Most of them receive antiretroviral treatment (ART),^1^ which can dramatically improve the life expectancy of people living with HIV.^2^ The long-term effectiveness of ART depends on lifelong retention on ART and high levels of adherence.^3,4^ Mental health problems often create challenges in treating HIV. They are associated with adverse HIV treatment outcomes, including suboptimal adherence,^5–7^ inadequate viral suppression,^8–12^ low retention in care,^13^ and premature mortality.^12^ Systematic reviews and meta-analyses consistently found strong associations between depression and anxiety and low adherence.^5–7^ The relationship between mental health problems and virologic treatment outcomes is less clear. Some studies showed associations between mental health disorders and poor viral suppression,^8–12^ while others found no association.^14–18^

Effective treatment of mental disorders among people living with HIV is critical to both improving mental health^19–22^ and managing HIV.^22–26^ However, there is a treatment gap in mental health care: mental disorders in people living with HIV in low- and middle-income countries often remain undiagnosed and untreated.^27,28^ In many settings, access to mental health services decreased during the COVID pandemic.^29,30^ The Friendship Bench (FB) intervention is a lay health worker-led psychological intervention developed to close the treatment gap for common mental disorders (CMD) such as anxiety and depression in Zimbabwe.^31^ The intervention reduced CMD symptoms in a cluster-randomized controlled trial in urban Zimbabwe,^31^ where the prevalence of depression or anxiety is high.^32,33^ However, the effectiveness of the FB intervention in rural settings and in improving adherence to ART and virologic suppression is unknown. In this cluster-randomized trial, we assessed the effect of the FB intervention on mental health symptoms and ART outcomes among adults living with HIV in the rural district of Bikita, where one in five adults living with HIV screened positive for CMD.^34^

## Methods

We conducted a pragmatic cluster-randomized open-label superiority trial in 16 health facilities in Bikita district in rural Zimbabwe (see <u>Supplement 1</u> for study protocol, <u>Supplement 2</u> for statistical analysis plan and ClinicalTrials.gov for trial registration NCT03704805). Participants were followed up every three months for one year between Oct 5, 2018, and Dec 18, 2020.

### Setting

Bikita is a rural district in Masvingo Province, about 300 km south of Harare. We selected 15 public facilities from the pool of 18 facilities participating in the International epidemiology Databases to Evaluate AIDS Southern Africa (IeDEA-SA).^35^ Three facilities were excluded because of low patient numbers, and one hospital site was added to balance intervention and control sites.

ART followed national treatment guidelines, except for one additional CD4 count at enrolment (baseline) and additional viral load testing (baseline, 6 and 12 months).^36^ Until April 2019, first-line regimens contained one non-nucleoside reverse transcriptase inhibitor (NNRTI) (efavirenz or nevirapine) and two nucleoside/nucleotide reverse transcriptase inhibitors (NRTI).^36^ Subsequently, individuals on ART with viral loads below 1000 copies/mL were switched to a dolutegravir-based regimen. Participants with a viral load > 1000 copies/mL received enhanced adherence counselling and an additional viral load test to confirm virologic failure.^37^ See <u>Supplement 3</u> and <u>Supplement 4</u> for further details.

### Randomization and masking

We randomized the 16 health facilities in a 1:1 ratio to intervention or control group. Randomization was stratified by the size of the health facility (three strata). We used block randomization with a block size of two within each stratum. Treatment assignment was known to participants, providers, evaluators, and data analysts. See <u>Supplement 4</u> for further details.

### Participants

The eligibility of interested individuals waiting for their ART clinic appointment was assessed using standardized questionnaires. Men and non-pregnant women aged 18 years or older from Bikita who spoke English or Shona, screened positive for CMD (Shona Symptoms Questionnaire [SSQ]-14 score ≥9), had received first-line ART for at least six months, had no WHO clinical stage 4 disease, no psychotic symptoms, and gave informed consent were eligible.

### Interventions

Participants in the intervention arm were offered the FB intervention in addition to enhanced standard of care (SC). Participants in the control arm received SC only.

The FB intervention consisted of weekly individual counselling sessions over six weeks and optional peer-led group support. Trained lay health workers delivered sessions, following a structured approach to identify problems, including adherence issues, and foster a positive attitude towards resolving them.

After four sessions, participants were invited to join a peer-led group activity where they were trained in income-generating skills (e.g. producing bags from recycled plastic). The FB intervention is described in detail elsewhere.^31,38^

SC consisted of nurse-led brief counselling, education, and support regarding CMD. Prescription of an antidepressant (fluoxetine) or referral to a psychiatric facility followed standard operating procedures (see <u>Supplement 3</u>). The nurses were trained in managing mental, neurological and substance use disorders.^39^

### Data collection

We used the Medication Event Monitoring System (MEMS) electronic pill box (AARDEX, Sion, Switzerland) to measure adherence. We calculated monthly mean adherence as the percentage of days participants opened the box once or twice (depending on the regimen). We treated adherence <10% as missing, assuming participants did not use their MEMS device. We assessed self-reported baseline adherence based on 30-day recall. <u>Supplement 3</u> provides further details.

Research assistants administered the SSQ-14^32,40^ and the Patient Health Questionnaire (PHQ-9)^32,41^ at the eligibility assessment and the 3, 6, 9, and 12-month visits. The SSQ-14 is a locally developed brief screening tool for CMD symptoms, including sleep disturbance, suicidal ideations, tearfulness, perceptual symptoms, and impaired functioning in the past seven days,^32^ with the score ranging from 0 (no symptom) to 14 (all symptoms present).^40^ The PHQ-9 is a brief screening tool for depression symptoms over the past two weeks. The SSQ-14 and the PHQ-9 have good psychometric properties in Zimbabwe’s HIV-positive and HIV-negative urban populations.^32,34^

At baseline, research assistants administered structured questionnaires on participants’ sociodemographic characteristics, perceived general health perception, ART knowledge, alcohol use (Alcohol Use Disorders Identification Test C [AUDIT-C]^42^), food insecurity (Household Food Insecurity Scale [HFIS]), and social support (Medical Outcomes Study Social Support Survey [MOS-SS]).

### Outcomes

The primary outcome was mean adherence between 2-6 months of follow-up. Secondary outcomes included mean adherence 1-12 months, change from baseline SSQ-14 and PHQ-9 score at 3, 6, 9, and 12 months and change in viral suppression (viral load <1000 copies per mL) at months 6 and 12, and positive screening for CMD (SSQ-14 score ≥9) and depression (PHQ-9 score ≥11) at 3, 6, 9, and 12 months.

### Sample size

The study was powered to detect a 10% difference in mean adherence^25^ between months 2-6, assuming a standard deviation of 20, an intra-cluster coefficient (ICC) of 0.05, and a type I error of 5% (see <u>Supplement 1</u>). Sixteen clusters with 25 participants provide 92% power to detect such a difference. To allow for attrition, we set the target sample size at 480 participants (16 clusters with 30 participants).

### Statistical analysis

The primary analysis was per intention to treat (ITT). We used linear mixed-effects models to assess the difference in mean adherence. Models included a random intercept and slope at the participant level to account for the correlation within participants. A random intercept accounted for the clustering of individuals in health facilities, and indicators defined treatment assignment, the month of analysis time, and interactions between the two. We estimated marginal odds ratios for the difference in the proportion of participants with viral suppression at 6 and 12 months using logistic mixed-effect models. We conducted prespecified adjusted analyses of mean adherence and viral suppression, controlling for facility size, age, and sex. In posthoc sensitivity analyses, we adjusted for facility size, age, sex, self-reported baseline adherence, baseline SSQ-14 score, ART regimen, PHQ-9 score, WHO clinical stage, CD4 cell count, viral suppression, AUDIT-C score, MOS-SS score, and travel cost. We also assessed the difference in change from baseline in SSQ-14 and PHQ-9 scores. We used the same mixed-effect models to assess the difference in the proportion of participants with common-mental disorders (SSQ-14 >9) and with depression (PHQ-9>11) at 3, 6, 9, and 12 months. We repeated analyses of primary and secondary outcomes using a per-protocol analysis, excluding participants who did not receive the allocated intervention and those with missing adherence data between months 2 to 6. Missing data were imputed using multiple imputation by chained equations.^43^ Analyses were run on 30 imputed datasets, and results combined using Rubin’s rule.^43^ <u>Supplement 3</u> provides further details.

### Ethics statement

The ethics committees of the Medical Research Council of Zimbabwe (MRCZ/A/2287), the Research Council of Zimbabwe, and the Canton of Bern, Switzerland (2018-00396) approved the study protocol. Individuals provided written informed consent to participate in the study.

## Results

Between Oct 5, 2018, and Dec 19, 2019, we assessed the eligibility of 3,706 individuals and excluded 3,190 participants (86.1%) (Figure 1). The most common reason for exclusion was an SSQ-14 score <9. Of the 595 participants who met inclusion criteria, 79 (13.3%) refused to participate or were lost to follow-up before the baseline visit (Table 1). The remaining 516 individuals (244 participants in eight clusters in the FB and 272 participants in eight clusters in the control group) were included. The last 12-month follow-up visit was on Dec 18, 2020.

**Table 1:**
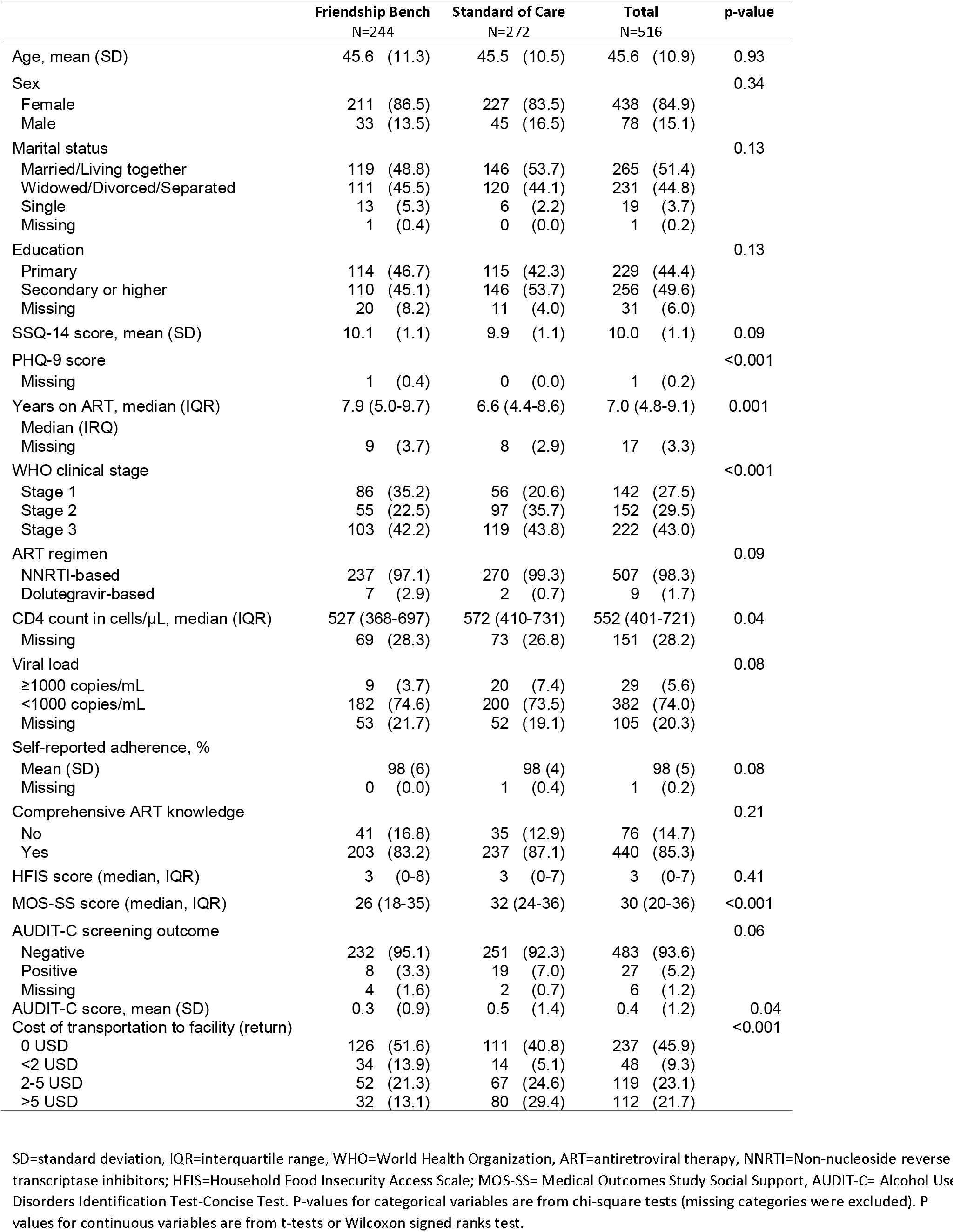
Sociodemographic and clinical characteristics of participants at baseline.

**Figure 1.**
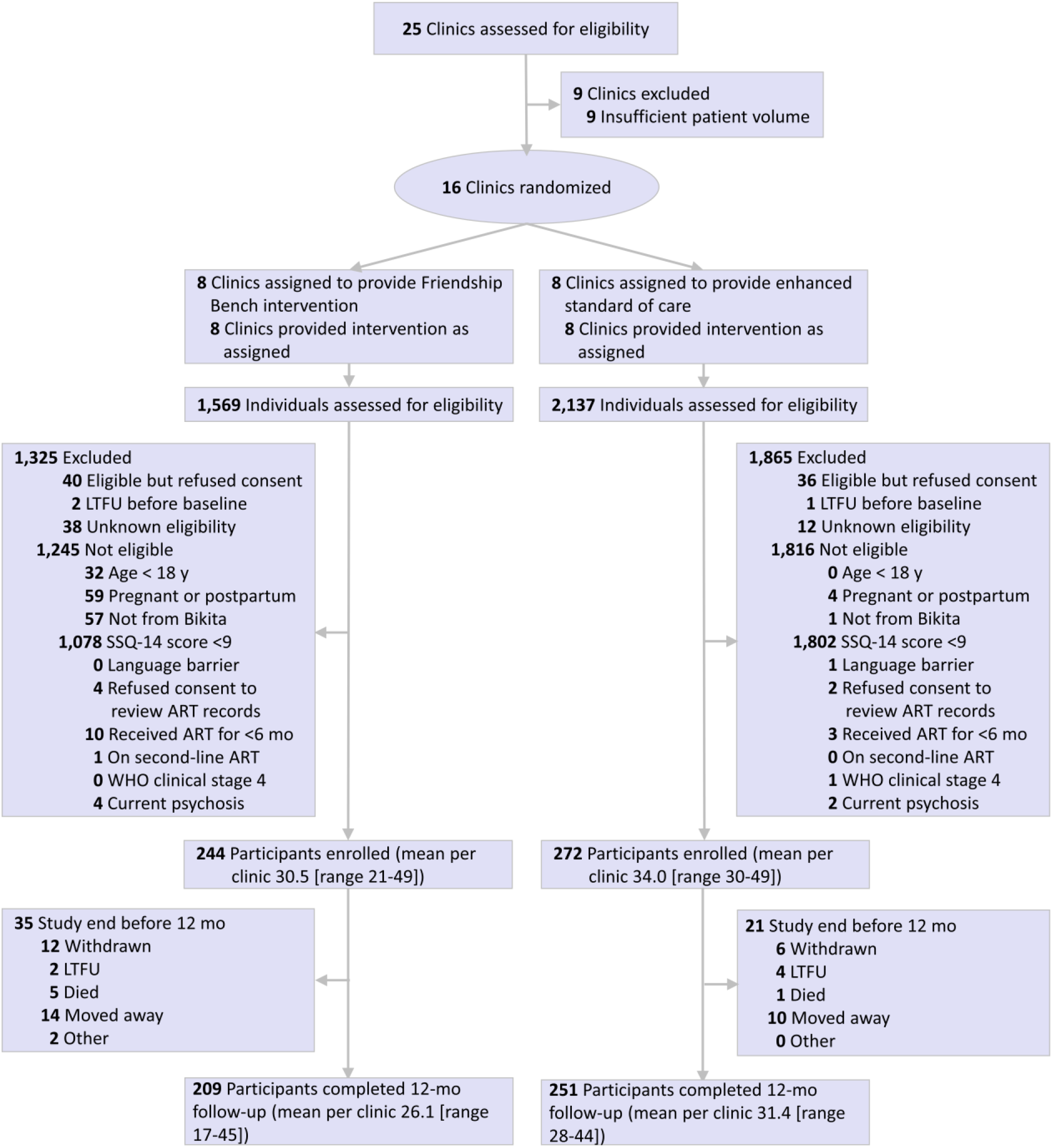
Trial profile.

**Figure 2.**
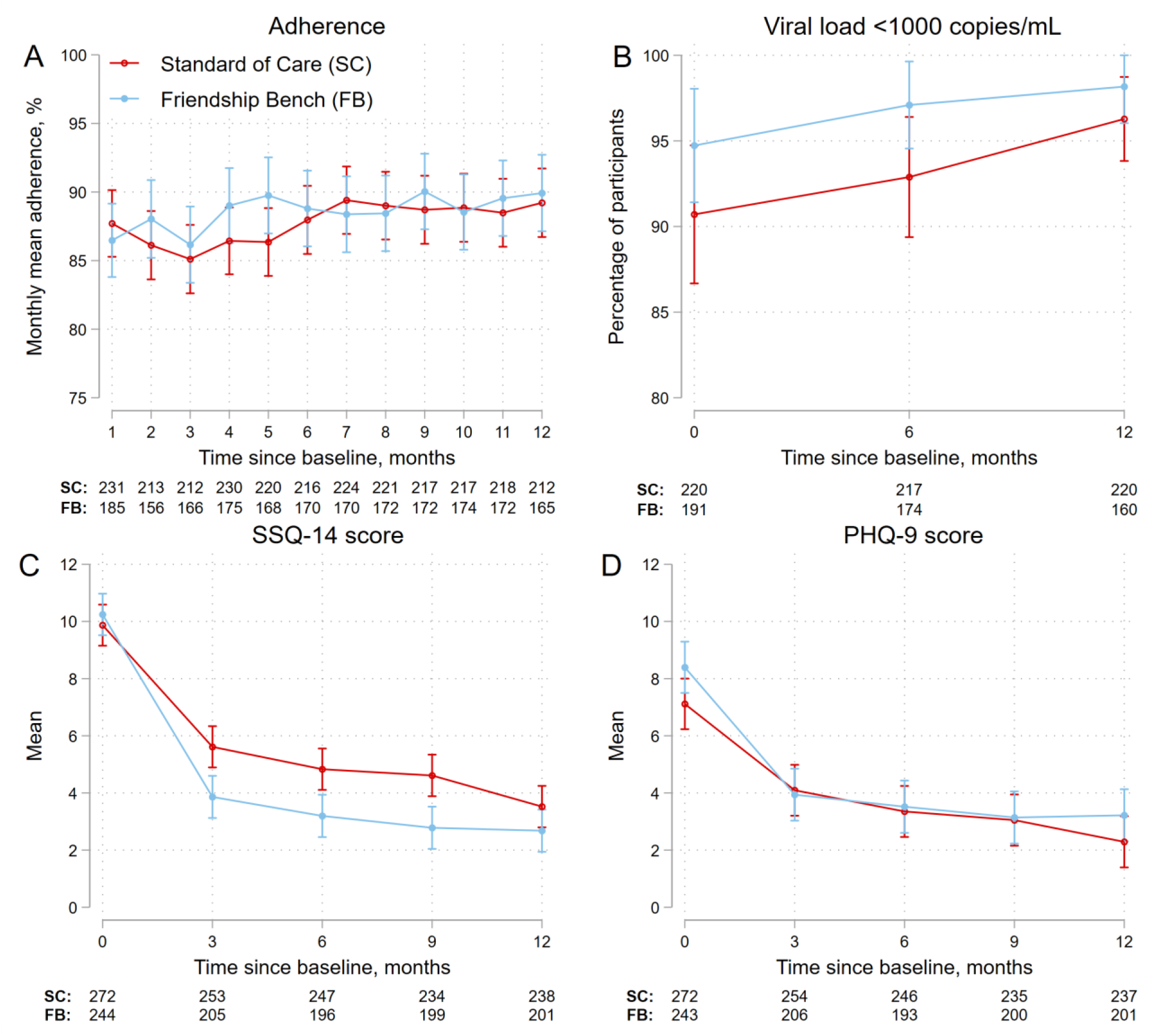
Adherence, viral load suppression, SSQ-14 score and PHQ-9 score by group. Figure shows mean monthly adherence scores (A), proportions of participants with viral load <1000 copies/mL (B), mean SSQ-14 scores, and mean PHQ-9 scores. Error bars represent 95% confidence intervals for means and proportions.

### Baseline characteristics

The mean age was 45.6 years (SD 10.9), and most participants were women (84.9%), married or cohabitating (51.4%) and had completed primary (44.4%) or primary and secondary education (46.7%). The mean SSQ-14 score at baseline was 10.0 (SD 1.1), and the mean PHQ-9 was 7.7 (SD 3.5). At enrolment, participants had been on ART for a median duration of 7.0 years (interquartile range [IQR] 4.8-9.1). Most (97.1%) were on an ART regimen containing tenofovir, lamivudine and efavirenz and in WHO clinical stage 1 (27.5%) or 2 (29.5%). The median CD4 cell count was 544 cells/µL (IQR 399-715), few participants (5.8%) had a viral load of ≥1000 copies/mL, and most (78.5%) reported optimal adherence (i.e. not having missed a single dose) in the 30 days before enrolment.

Participants in the intervention group had a higher baseline mean PHQ-9 score, were more likely to be in WHO clinical stage 1, had lower MOS-SS and AUDIT-C scores, and had lower transportation costs than participants in the control group (Table 1).

### Retention

Retention in the FB and adherence interventions was high (Table 2). All except two participants in the intervention group received the nurse-led brief support counselling. In both groups, the median time from the baseline visit to the date of the counselling session was 0 days (IQR 0-0). Few participants (2.5% in the FB arm and 6.3% in the SC arm) received antidepressants. The median time from baseline to the first FB session was 3 days (IQR 0-8). The median number of sessions received among participants in the intervention group was 6 (IQR, 6-6); 88.1% of participants received all six sessions. In the intervention group, the proportion of participants who attended at least one group support session within a 3-month interval decreased from 67.6% to 51.2% from the first to the last interval. Most participants with ≥1000 copies/mL at the baseline visit (72.7% and 76.9% in FB and SC groups, respectively) attended all three adherence counselling sessions.

**Table 2:** Fidelity to mental health and adherence interventions.

|  | **Friendship Bench** |  | **Standard of Care** |  | **Total** |  |
| --- | --- | --- | --- | --- | --- | --- |
|  | N=244 |  | N=272 |  | N=516 |  |
| Received allocated intervention | 223 | (91.4) | 272 | (100) | 495 | (95.9) |
| **Friendship Bench intervention** |  |  |  |  |  |  |
| Individual counseling session 1 attended | 235 | (96.3) | - | - | 235 | (45.5) |
| Individual counseling session 2 attended | 230 | (94.3) | - | - | 230 | (44.6) |
| Individual counseling session 3 attended | 229 | (93.9) | - | - | 29 | (44.4) |
| Individual counseling session 4 attended | 224 | (91.8) | - | - | 224 | (43.4) |
| Individual counseling session 5 attended | 219 | (89.8) | - | - | 219 | (42.4) |
| Individual counseling session 6 attended | 215 | (88.1) | - | - | 215 | (41.7) |
| Group support attended |  |  |  |  |  |  |
| Between months 0 and 3 | 165 | (67.6) | - | - | 165 | (32.0) |
| Between months 4 and 6 | 146 | (59.8) | - | - | 146 | (28.3) |
| Between months 7 and 9 | 141 | (57.8) | - | - | 141 | (27.3) |
| Between months 10 and 12 | 125 | (51.2) | - | - | 125 | (24.2) |
| **Standard of care** |  |  |  |  |  |  |
| Nurse-led counseling attended | 242 | (99.2) | 272 | (100) | 514 | (99.6) |
| Antidepressant prescribed | 6 | (2.5) | 17 | (6.3) | 23 | (4.5) |
| **Enhanced adherence counselling** |  |  |  |  |  |  |
| VL ≥1000 copies/mL at baseline visit | 11 | (100) | 26 | (100) | 37 | (100) |
| 1-2 sessions attended | 2 | (18.2) | 5 | (19.2) | 7 | (18.9) |
| 3 sessions attended | 8 | (72.7) | 20 | (76.9) | 28 | (75.7) |
| No session attended | 1 | (9.1) | 0 | (0.0) | 1 | (2.7) |
| Unknown | 0 | (0.0) | 1 | (3.8) | 1 | (2.7) |
| VL ≥1000 copies/mL 6-month visit | 6 | (100) | 17 | (100) | 23 | (100) |
| 1-2 sessions attended | 0 | (0.0) | 5 | (29.4) | 5 | (21.7) |
| 3 sessions attended | 5 | (83.3) | 10 | (58.8) | 15 | (65.2) |
| No session attended | 1 | (16.7) | 1 | (5.9) | 2 | (8.7) |
| Unknown | 0 | (0.0) | 1 | (5.9) | 1 | (4.3) |
| VL ≥1000 copies/mL at 12-month visit | 3 | (100) | 9 | (100) | 12 | (100) |
| 1-2 sessions attended | 0 | (0.0) | 2 | (22.2) | 2 | (16.7) |
| 3 sessions attended | 0 | (0.0) | 2 | (22.2) | 2 | (16.7) |
| Unknown | 3 | (100) | 5 | (55.6) | 8 | (66.7) |

### Outcomes

In unadjusted analyses, mean ART adherence between months 2 and 6 was slightly higher in the FB than in the SC group: mean difference 1.93%, 95% CI -1.20 to 5.06, P=0.23, but there was little difference in mean ART adherence after month 6 (Figure 1A, Table 3). Overall, between months 1 and 12, mean ART adherence was similar (mean difference 0.79%, 95% CI -2.14 to 3.71, P=0.60) (Table 3). The odds of virologic suppression at 6 months (OR 2.20, 95% CI 0.79 to 6.14, P=0.13) and 12 months (OR 1.60, 95% CI 0.42 to 6.05, P=0.49) were higher in the FB than in the SC group, but differences failed to reach conventional levels of statistical significance (Figure 1B, Table 3). Declines in SSQ-14 scores were more pronounced (P<0.05) in the FB than in the SC group up to month 9, with scores converging by month 12 (Figure 1C, Table 3) whereas declines in PHQ-9 scores were similar (Figure 1D, Table 3). There was little evidence for a difference in the odds of screening positively for common mental disorders (SSQ-14 >9) or depression (PHQ-9>11) at 3, 6, 9 or 12 months (Table 3). Results were similar in the prespecified adjusted (Table 3) and post hoc sensitivity analyses for the ITT and per-protocol populations (<u>Supplement 3</u>).

**Table 3.**
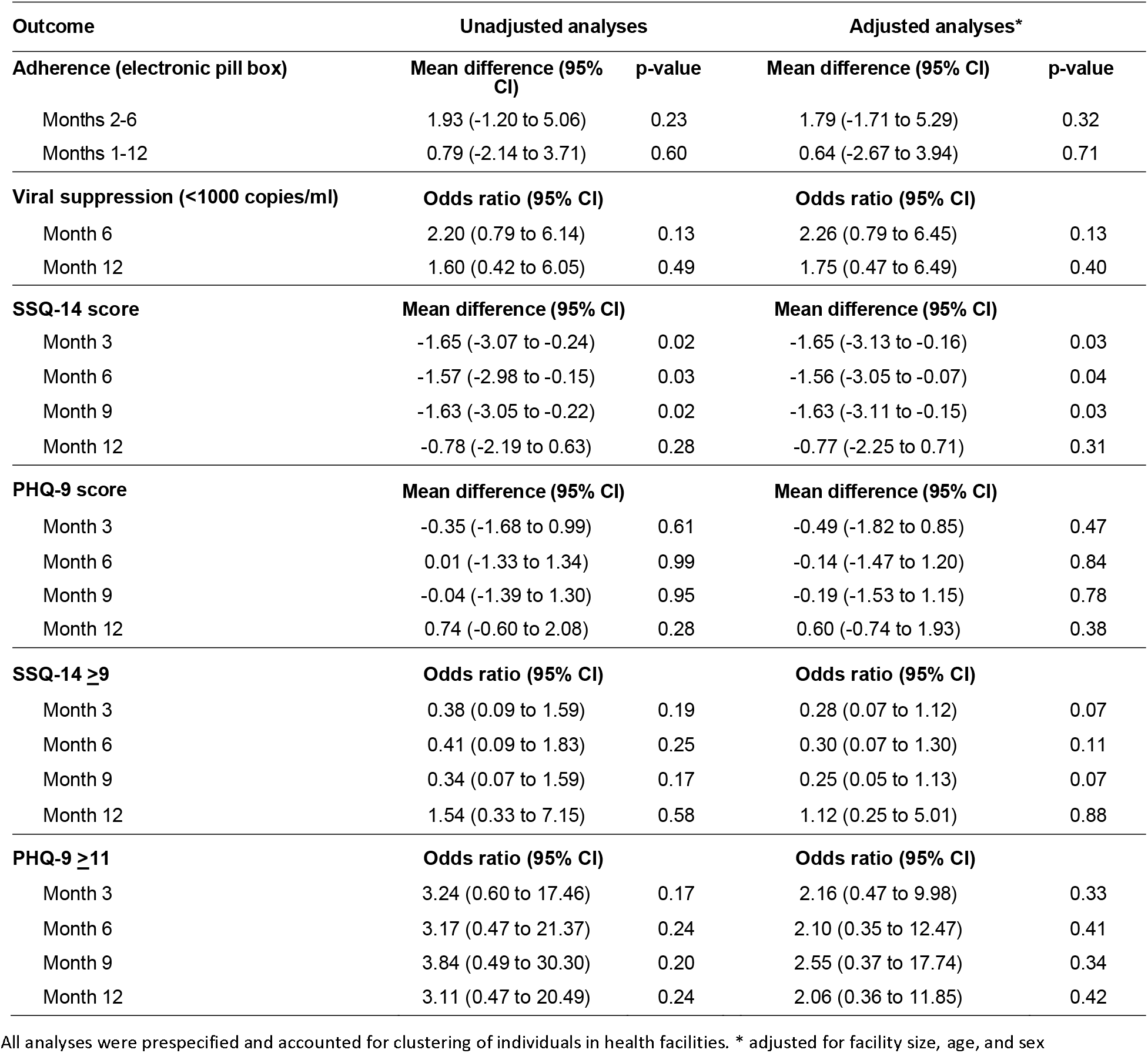
Effect of the friendship bench intervention on adherence, viral load and mental health.

In total, 16 participants reported self-harm or attempted self-harm: 11 (4.5%) in the FB arm and 5 (1.8%) in the SC arm. In the FB group, 9 (3.7%) participants reported a history of self-harm before baseline, and 2 (0.8%) participants reported a new incident self-harm event after baseline. No new incident self-harm event after baseline was reported in the SC group. No psychiatric hospitalizations occurred. Five participants died in the FB arm and one in the SC arm. All deaths were unrelated to study procedures, and no suicides were recorded or suspected (<u>Supplement 3</u>).

## Discussion

In this cluster-randomized trial, we examined the effect of the FB intervention on ART outcomes and mental health symptoms in adults living with HIV in rural Zimbabwe. Implementing this psychological intervention led by lay health workers in rural health facilities proved feasible. The intervention was well accepted by participants, with almost 90% attending all six individual counselling sessions. Participants in the FB group had a more pronounced decrease in symptoms of CMD (SSQ-14 scores) than those in the control group. However, the intervention showed no clear effect on symptoms of depression (PHQ-9 scores), ART adherence or viral suppression. Our study documents a substantial need for mental health services among people living with HIV in rural Zimbabwe. About one in five adults living with HIV in our study had significant symptoms of CMDs.^34^

With the present study, three trials now show that the FB intervention can reduce symptoms of CMDs (<u>eFigure 1 in Supplement 3</u>). The first was a cluster-randomized trial in primary care facilities in Harare.^31^ Participants in the intervention group had a greater decrease in CMD and depression symptoms from baseline to 6 months than participants in the enhanced usual care group.^31^ In our trial in rural Zimbabwe, the difference in CMD symptoms was less pronounced, and there was no effect on symptoms of depression. The first trial was done in an urban setting and included people with and without HIV: 42% were known to be HIV-negative.^31^ Also, the Harare population was about 10 years younger than the rural study population. Interestingly, decreases in the intervention group in symptoms of CMD and depression (SSQ-14 or PHQ-9 scores) between baseline and 6 months were similar in the two studies. The lower effectiveness in the present study was due to differences in the rate of change in the control group: in the urban trial,^31^ the scores remained high at the 6-month assessment, whereas in the rural setting, the mean scores decreased both in the intervention and control arms. In the rural population, enhanced usual care appears to have been more effective than in the urban setting. The third trial included adolescents living with HIV and symptoms of CMDs in Zimbabwe.^44^ The intervention improved symptoms of CMDs and depression compared to standard care among adolescents living with HIV in Zimbabwe.^44^ Of note, differences in SSQ-14 and PHQ-9 scores were relatively small, with about 1 score point difference favoring the intervention at 48 weeks.^44^

Two of the three trials examined virologic suppression (this study and the trial among adolescents^44^), and both failed to demonstrate any statistically significant effect (<u>eFigure 1 in Supplement 3</u>). The present study also examined adherence and equally showed no robust intervention effect. A ceiling effect may have played a role in our study, as over 90% of participants had viral suppression and adherence was close to 90%. Given the well-established association between CMD and poor HIV outcomes,^5–12^ we expected lower baseline viral suppression and adherence rates. The large proportion of women in our study and the relatively old age will have contributed to these high rates. In a recent analysis of a large South African ART program, we found considerably higher viral suppression and adherence rates in women than men and higher rates in older than in younger people.^45^ A ceiling effect is less likely for the trial among adolescents where more than one-third of participants had a viral load of 1000 copies/ml or above.^44^ Indeed, the nature of the FB intervention, which does not include skill-based adherence training, might be a more pervasive explanation for the lack of effect on viral suppression. Mental health interventions with integrated skill-based adherence training have been effective.^45^ For example, a recent randomized controlled trial of South African adults with depression and unsuppressed viral load showed that nurse-delivered cognitive-behavioral therapy improved depression scores, ART adherence, and viral suppression.^26^ Of note, a small pilot trial in Zimbabwe of a version of the FB intervention that incorporated skill-based training in adherence showed promising results regarding improved adherence, viral load suppression and depression.^46^

The large sample size and pragmatic study design, which closely resembled real-world conditions in rural Zimbabwe, are strengths of our study that add to the generalisability of findings. Apart from the initial support provided by the study team during the training, the lay health workers operated under real-world conditions. Further strengths of our study include using electronic pill bottles for adherence monitoring and using the SSQ-14, a screening tool to measure CMD symptoms in Shona people.^32^ There is concern that electronic monitoring may affect adherence, but little evidence supports this contention.^47,48^ Men were underrepresented in our study (and the Harare study^31^), and it remains unclear how well the FB intervention works for them. The poor representation of men may reflect a lack of interest in mental health interventions. Finally, the mental health screening tools used in this study (SSQ-14 and PHQ-9) have been validated in people living with HIV in Zimbabwe’s capital Harare but not in the rural study setting.^32^

## Conclusion

The FB intervention is feasible and acceptable in rural Zimbabwe to closing the treatment gap in mental health care in rural Zimbabwe. The FB intervention improved CMD symptoms but the intervention effect was smaller than previously shown in an urban setting. We could not demonstrate an effect of the intervention on adherence and viral suppression. More work is needed to evaluate the effect of the approach on HIV outcomes in different populations, including young adults and men, and populations with more severe symptoms at higher risk of non-adherence and virologic failure. The further development of the FB intervention to incorporate adherence training may be a promising approach to reach those at high risk of poor HIV outcomes.

## Data Availability

Data cannot be made available online because of legal and ethical restrictions. To request data, readers may contact IeDEA-SA for consideration.

## Article information

### Open Access

This is an open access article distributed under the terms of the CC-BY License. © Haas AD et al. JAMA Network Open.

### Members of the FB-ART Trial Group

Study sponsor: Prof Matthias Egger; Principal investigators: Dr Andreas D Haas and Dr Cordelia Kunzekwenyika; Project manager: Stefanie Hossmann; Study coordinator: Josphat Manzero; Senior statistician: Dr Andreas Limacher; Statistician: Dr Andreas Haas; Friendship Bench: Dr Ruth Verhey (Lead of Global Training & Research); SolidarMed: Dr Janneke H van Dijk (Country Director, Zimbabwe), Dr Michael André Hobbins (Head of Research), Dr Ronald Manhibi (IT manager), Joseph Bishi (IT manager); IeDEA Southern Africa: Dr Per von Groote (Program Manager); Research assistants (RAs): Amos Kateta, Cuthbert Matonhodza, Favourite Machiha, Ntandoyenkosi Mhlanga, Shingai Matuturu, Tatenda Gombirwo, Millicent Gweredza, Josephine Saide, Beauty Muchakubvura, Kudzai Mhlanga, Dennis Mwakasa, Fungai Nyikadzino Zvekare, Yvonne Maumbe, Boldwin Maposa, Sibongile Gumbo, Shingai Matutururu, Japhet Kamusha, Morris Tshuma, Rejoyce Runyowa, Abigail Pikai, Fredrick Mbiva, Elizabeth Mutungama, Waraidzo Mukuwapasi, Rudorwashe Mandabva

### Authors and contributors

AH, SH, JvD, RV and ME conceptualized the study. ME acquired the funding. AH, JM, SH and RM developed electronic case report forms. CK, JM, SH, RM, and RV developed SOPs and training materials. RAs enrolled study participants and collected data. AH, CK, JM, SH and RM curated the dataset. CK, JM, SH, JvD, PvG, and MH contributed to project administration. AH, CK, JM, SH, JvD, RV, PvG, MH, and ME trained or supervised members of the study team or lay health workers. AH did statistical analysis. AL advised on statistical methods and independently reproduced statistical analysis. AH, and ME wrote the original draft. All authors critically revised the manuscript for important intellectual content. AH had full access to all data in the study and takes responsibility for the integrity of the data and the accuracy of the data analysis.

### Conflict of Interest Disclosures

No disclosures were reported.

### Funding/Support

The research reported in this publication was supported by the U.S. National Institutes of Health’s National Institute of Allergy and Infectious Diseases, the Eunice Kennedy Shriver National Institute of Child Health and Human Development, the National Cancer Institute, the National Institute of Mental Health, the National Institute on Drug Abuse, the National Heart, Lung, and Blood Institute, the National Institute on Alcohol Abuse and Alcoholism, the National Institute of Diabetes and Digestive and Kidney Diseases and the Fogarty International Center under Award Number U01AI069924. AH was supported by an Ambizione fellowship (193381) and ME by special project funding (189498) from the Swiss National Science Foundation. The content is solely the responsibility of the authors and does not necessarily represent the official views of the National Institutes of Health or the Swiss National Science Foundation.

#### Role of the Funder/Sponsor

The funders had no role in the design and conduct of the study; collection, management, analysis, and interpretation of the data; preparation, review, or approval of the manuscript; and decision to submit the manuscript for publication.

### Disclaimer

The views and conclusions contained herein are those of the authors and should not be interpreted as necessarily representing the official policies or endorsements, either expressed or implied, of the NIH or the US government.

## SUPPLEMENTARY APPENDIX

### Supplement 1

**Trial protocol**

### Supplement 2

**Statistical analysis plan**

### Supplement 3

**eMethods** Additional information on standard care, randomization, adherence measures, and statistical analyses

**eTable 1** Effect of the Friendship Bench intervention on adherence, viral load and mental health: results from prespecified and posthoc sensitivity intention to treat analyses

**eTable 2** Effect of the Friendship Bench intervention on adherence, viral load and mental health: results from prespecified and posthoc sensitivity per protocol analyses

**eTable 3** Number of participants with safety-relevant outcomes by trial arm

**eFigure 1** Forest plot of available evidence of the effect of the Friendship Bench intervention on SSQ-14 scores, PHQ-9 scores, and viral suppression from three trials

### Supplement 4

Operational and service delivery Manual for the prevention, care and treatment of HIV in Zimbabwe

## Notes

### Competing Interest Statement

The authors have declared no competing interest.

### Clinical Trial

ClinicalTrials.gov Identifier: NCT03704805

