## Supplement 2 for "Effect of the Friendship Bench intervention on antiretroviral therapy outcomes and mental health symptoms in rural Zimbabwe: A cluster randomized trial"

### Statistical Analysis Plan (SAP)

#### Effect of a psychological intervention on antiretroviral therapy outcomes and symptoms of common mental disorders in HIV-positive adults in rural Zimbabwe: cluster-randomized trial

#### Friendship Bench Trial

### Administrative Information

| | |
|---|---|
| Project number: | 794 |
| Trial registration number: | NCT03704805 |
| SAP version: | Version 1.0, April 29, 2019 |
| Protocol version: | Version 4, April 05, 2018 |

#### Contributors

| Name | Affiliation | Role in SAP writing |
|---|---|---|
| Andreas Limacher | CTU Bern | Author |
| Stefanie Hossmann | CTU Bern | Author |
| Andreas Haas | ISPM Bern | Reviewer |

#### Revision history

| Revision | Justification | Timing |
|---|---|---|
| n.a. (first version) | | |

#### Approved by

| Name | Affiliation | Study Role | Date and Signature (wet ink) |
|---|---|---|---|
| Andreas Haas | ISPM Bern | Principal Investigator, Trial Statistician | 06/05/19 |
| Andreas Limacher | CTU Bern | Senior Statistician | 6.5.19 |
| Stefanie Hossmann | CTU Bern | Clinical Trial Manager | 6.5.19 |

6.4 Missing data ... 19

6.5 Safety evaluation ... 20

6.6 Statistical software ... 20

6.7 Quality control ... 20

**7. Changes from the protocol ... 21**

**8. References ... 22**

### 1. Introduction

#### 1.1 Background and rationale/

Common mental disorders are highly prevalent among people living with HIV. Left untreated, common mental disorders cause substantial disability and undermine individuals' ability to adhere to antiretroviral therapy (ART), leading to poor HIV treatment outcomes. Management of mental disorders is a promising strategy to improve both mental health and HIV treatment outcomes among people on antiretroviral therapy who also suffer from common mental disorders. A recent cluster-randomized controlled trial from Harare, Zimbabwe showed that problem-solving therapy delivered by lay health workers effectively reduced symptoms of common mental disorders, but the effect of the intervention on HIV treatment outcomes and its effectiveness in the rural setting has not been studied. We aim to examine the effect of problem-solving therapy delivered by lay health workers on HIV treatment outcomes in rural Zimbabwe.

#### 1.2 Objectives

Primary objective: To assess the effect of problem-solving therapy on HIV treatment outcomes.

Secondary objectives: To assess the effectiveness of problem-solving therapy on symptoms of common mental disorders among rural HIV-infected populations and to assess the prevalence of common-mental disorders among rural HIV-infected population on ART.

### 2. Study methods

#### 2.1 Trial design

The study is designed as a cluster-randomized, controlled, parallel, two-arm multicenter trial with 1:1 allocation ratio.

#### 2.2 Randomization

Health facilities will be assigned in a 1:1 ratio to the intervention or the control group using a computer-generated, stratified blocked randomization. Randomization will be stratified by clinic size (<280, 280-1000, >1000 active ART patients). To avoid imbalance in the size of the groups, we will use blocked randomization within each stratum, with blocks consisting of two health facilities. The unit of random allocation will be the health facility and not the individual participant. This will minimize the risk of contamination between study arms. Randomization of health facilities was done by a statistician who is not part of the study team at CTU Bern, Switzerland. Randomization will be done prior to initiation of the recruitment phase.

#### 2.3 Sample size

We will recruit 480 participants (16 clusters with 30 participants each). See section 11.2 in the study protocol (version 4) for determination of sample size.

#### 2.4 Framework

Trial to show superiority of problem-solving therapy (intervention) over enhanced standard of care (control).

#### 2.5 Statistical interim analyses and stopping guidance

There is no interim analysis planned.

#### 2.6 Timing of final analysis

Final analysis will take place once all follow-up activities are completed, all data is entered in the database, and all data is statistically validated and cleaned in the database. Before starting the analysis the database will be locked and a final export of study data will be done.

#### 2.7 Timing of outcome assessments

Study outcomes are assessed at 3, 6, 9, and 12 months after recruitment.

#### 2.8 Blinding

Treatment assignment is unblinded to participants, lay health workers, health care providers, evaluators, data managers, and trial statistician.

The senior statistician authoring the SAP is blinded while writing the SAP, during the whole conduct of the study and final analysis. The senior statistician is only unblinded before quality control (i.e. double programming) of the primary analysis.

### 3. Data management

#### 3.1 Data export

The CRFs in this trial are implemented electronically using a dedicated electronic data capturing (EDC) system (REDCap, https://www.project-redcap.org/). All data entered in the eCRFs are stored on a Linux server in a dedicated mySQL database. Responsibility for hosting the EDC system and the database lies with CTU Bern.

Data on medication adherence is collected with the Medication Event Monitoring System ([MEMS], AARDEX, Sion, Switzerland). Adherence is measured with patient-held electronic pill bottles (MEMS caps) that record the dosing history of a patient (i.e. the time of each bottle opening). Research assistants download data recorded on MEMS caps at 3-monthly follow-up visits using Android tablets and synchronize the tablets with the medAmigo web platform. MedAmigo is an online web-solution to retrieve and transfer dosing history data from MEMS caps through the internet to dedicated servers. The dosing history data are stored on centralized, secured servers and regular backups are scheduled. Responsibility for hosting the medAmigo system and the database lies with the manufacturer of the MEMS (AARDEX, Sion, Switzerland). At final analyses, data files will be extracted from the database and imported into a statistical software package according to the SOP for data preparation and programming[i].

#### 3.2 Data validation

First line data validation is performed by the online eCRF system at real-time as defined in the data dictionary. Second line data validation and cleaning will be performed after completion of data entry but before database lock according to the SOP for data validation[ii].

Data checks are described in the central data-monitoring plan.

#### 3.3 Data preparation

##### 3.3.1 Adherence data (MEMS data)

Participants' 1-monthly (30-days) mean adherence score is calculated as the percentage of doses correctly taken per interval. For regimens that have to be taken once daily, a dose is considered as taken if the participant opens the electronic pill bottle within -8 to +16 hours of the designated dosing time. For regimens that have to be taken twice daily, a dose was considered as taken if the participant opens the electronic pill bottle within -4 to +8 hours of the designated dosing time. The designated dosing time is specific to each patient and recorded in the eCRF.

##### 3.3.2 SSQ-14

The SSQ-14 is a dichotomous 14-item questionnaire. Each item where the answer is yes is scored as 1, providing a score ranging from 0 to 14.

##### 3.3.3 PHQ-9

Nine items, each of which is scored 0 to 3 on a Likert scale, providing a severity score ranging from 0 to 27.

##### 3.3.4 Viral load suppression

Will be assessed as dichotomous considering <1000 copies per milliliter as suppressed.

###### 3.3.5 Safety outcomes

Psychiatric hospitalization and suicide attempts are reported in variables rf_1 and rf_2, which are assessed at each visit.

Suicides are assessed based on the unscheduled serious adverse event form, using the variables sae_type, sae_description, sae_type_fup1, sae_description_fup1, sae_type_fup2, and sae_description_fup2. Sae_description are free text fields and will be assessed by an appropriately trained person.

###### 3.3.6 Data sharing

Data sharing is described in the data management plan.

### 4. Statistical principles

#### 4.1 Confidence intervals and *P* values

All applicable statistical tests will be 2-sided and will be performed using a 5% significance level. 95% confidence intervals will be reported.

#### 4.2 Analysis populations

##### 4.2.1 Full analysis set (FAS)

The full analysis set (FAS) will include all eligible participants who consented to participate in the study and attended the baseline visit. Following the intent-to-treat principle, subjects will be analyzed according to the treatment which was assigned to the site they are enrolled at.

Table 1: Violation of eligibility criteria

| Protocol deviation | eCRF visit | Variable | Variable type | Violation |
|---|---|---|---|---|
| Violation of inclusion or exclusion criteria | | | | |
| Age >18 years | Recruitment | es_2 | Continuous: years | es_2<18 |
| On first-line antiretroviral therapy for at least 6 months | Recruitment | es_4 and<br>es_14 – es_3_date and<br>es_18 | Dichotomous: multiple choice<br>Continuous: days<br>Dichotomous: multiple choice | es_4 = 2, -8, -9<br>es_14 – es_3_date < 180<br>es_18 = 7, -7 |
| Resident in Bikita District | Recruitment | es_6 | Dichotomous: multiple choice | es_6 = 2, -8, -9 |
| Speak and understand English or Shona | Recruitment | es_8 | Dichotomous: multiple choice | es_6 = 0 |
| Able to comprehend the information on the study | Recruitment | es_21 | Dichotomous: multiple choice | es_21 = 0 |
| Screened positive for CMDs (Shona Symptoms Questionnaire ≥9) | Recruitment | ssq14_1_score | Continuous: score | ssq14_1_score < 9 |
| Provided written informed consent (consent by thumbprint) to participate study | Recruitment | es_21 | Dichotomous: multiple choice | es_21 = 0 |
| Current psychosis / cognitive impairment | Recruitment | ssq14_1_5 and<br>es_27 | Dichotomous: multiple choice<br>Dichotomous: multiple choice | ssq14_1_5 = 1<br>es_27 = 1, -7 |
| Clinical AIDS (WHO clinical stage 4) | Recruitment | es_23 | Dichotomous: multiple choice | es_23 = 4, -7 |
| Known pregnancy or ≤3 months postpartum | Recruitment | es_5 | Dichotomous: multiple choice | es_5 = 1, -8, -9 |

##### 4.2.2 Per-protocol (PP)

Per-protocol population consists of all subjects in the FAS who do not have any protocol deviations that could confound the interpretation of analyses conducted on the FAS. The following are common major protocol deviations:

- Not receiving allocated intervention
- Completely missing primary outcome data (2-6 months)

Table 2: Derivation of protocol deviations.

| Protocol deviation | eCRF visit | Variable | Variable type | Derivation |
|---|---|---|---|---|
| Not receiving allocated intervention | | | | |
| Attending less than four friendship bench individual counselling sessions for participants in the intervention group. | Friendship Bench 1 - 4 | fb_date | Continuous: date | fb_date = less than 4 completed |
| Attending no nurse-led mental health counselling for participants in the control and intervention group. | Nurse-led mental health counselling | nlc_yn | Dichotomous: multiple choice | nlc_yn = 2, -8 or -7 |
| Receiving un-allocated intervention | | | | |
| Attending one or more friendship bench individual or group counselling sessions for participants in the control group. | Any Friendship Bench Visit | fb_date | Continuous: date | fb_date = any completed |
| Completely missing primary outcome data (percentage adherence at month 2 to month 6). | MEMS data | adh2 – adh6 | Continuous | adh2-adh6 all missing |

##### 4.2.3 Safety population

The safety population consists of all subjects in the FAS. Subjects will be analyzed according to the treatment they actually received.

#### 4.3 Estimands

The ICH E9 (R1) addendum on estimands and sensitivity analyses defines different treatment estimators that are of interest in clinical trials (EMA 2017). An estimand is the target of estimation to address the scientific question of interest posed by the trial objective. Attributes of an estimand include the population of interest, the outcome of interest, the specification of how intercurrent events such as cross-overs and non-compliance are reflected in the scientific question of interest, and the population-level summary for the outcome.

##### 4.3.1 Treatment policy estimand

The estimand that addresses the main objective of this trial is based on the treatment policy strategy. The value for the outcome of interest is used regardless of whether or not the intervention was performed and/or recommendations war applied. The primary intention-to-treat analysis of the primary as well as secondary outcomes will be based on this estimand using the FAS.

- Primary outcome of interest: Mean adherence between 2-6 months
- Patient-set of interest: FAS
- Handling of intercurrent events: Outcome data will be used anyway; if missing, outcome data will be imputed
- Population-level summary measure of outcome: Difference in mean adherence between 2-6 months

##### 4.3.2 Per Protocol analysis

We will perform a per-protocol analysis. Subjects with major protocol deviations will be excluded from this analysis.

#### 5. Trial Population

##### 5.1 Screening data

We will draw a flow diagram following the CONSORT extension for Cluster Trials 2012 (http://www.consort-statement.org/extensions/overview/cluster-trials). In addition to the number, average cluster size, and variance of cluster size, we will report number and percentage of patients.

Flowchart 1: CONSORT

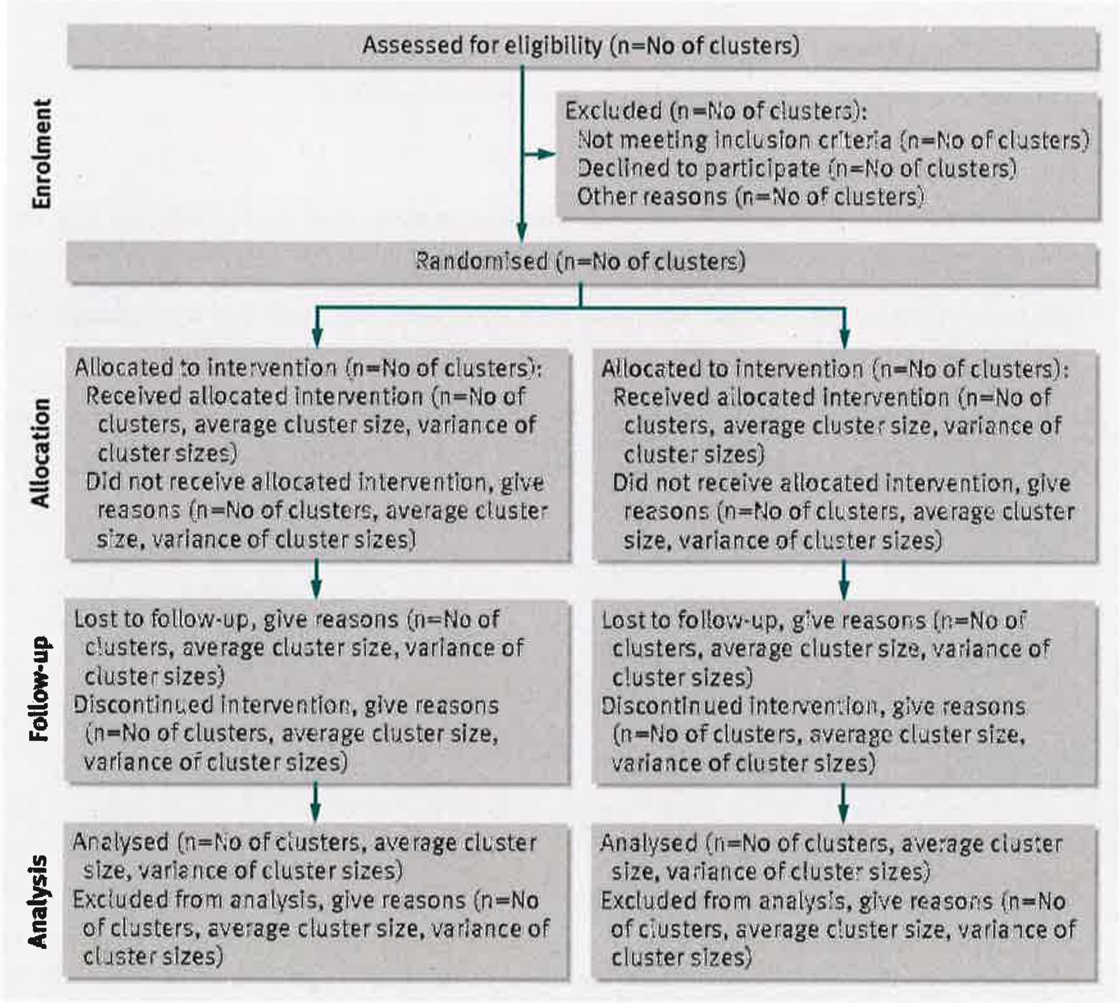

##### 5.2 Eligibility

Inclusion criteria

- Age >18 years
- On first-line antiretroviral therapy for at least 6 months
- Resident in Bikita District
- Speak and understand English or Shona
- Able to comprehend the information on the study

- Screened positive for CMDs (Shona Symptoms Questionnaire ≥9)
- Provided written informed consent (consent by thumbprint) to participate study

Exclusion criteria

- Current psychosis / cognitive impairment
- Clinical AIDS (WHO clinical stage 4)
- Known pregnancy or ≤3 months postpartum

##### 5.3 Recruitment

Information is included in the CONSORT flow diagram (flowchart 1).

##### 5.4 Cluster characteristics

Cluster characteristics at baseline will be presented in a table, stratified by treatment arm, as number and percentage or mean and standard deviation for categorical and normally distributed continuous variables, respectively. For data severely deviating from a normal distribution, we will present median and interquartile ranges. Groups will not be compared statistically because any significant difference can be explained by the play of chance if the randomization was performed properly.

Table 2: Cluster characteristics

| Description | Variable | Type |
|---|---|---|
| Type of clinic | cluster_type | Categorical: multiple choice |
| Clinic size (active ART patients) | cluster_size | Categorical: multiple choice |
| Average travel time to reach facility | b_hs_4 | Continuous: Traveling hours |
| Average travel cost to reach facility | b_hs_7 | Continuous: Dollars |
| Average duration of visit at facility | b_hs_6 | Continuous: Hours at clinic |

##### 5.5 Baseline patient characteristics

The patient characteristics of the FAS at baseline will be presented in a table, stratified by treatment arm, as number and percentage or mean and standard deviation for categorical and normally distributed continuous variables, respectively. For data severely deviating from a normal distribution, we will present median and interquartile ranges. We will show p-values for differences between the two groups because cluster-randomized trials lack the balancing properties of classical randomized trails. Intervention groups will be compared by the t-test for continuous data or by the non-parametric Wilcoxon test when normality assumption is not satisfied. For categorical data, Fisher's exact test will be used if expected frequencies are lower than 5 in any cell, else the chi-squared test will be applied.

Table 3: Patient characteristics table.

| Description | Variable | Type |
|---|---|---|
| Age | es_14 | Continuous: dates |

| Description | Variable | Type |
|---|---|---|
| Gender | gender | Binary: male, female |
| Marital status | b_marital | Categorical: married or living together, widowed, divorced, or separated, single. |
| Education | b_school, b_years | Categorical: None or primary, at least secondary |
| Religion | b_religion | Categorical: Christian or other |
| SSQ-14 score | ssq14_1_score | Continuous: score |
| PHQ 9 Score | phq9_1_score | Continuous: score |
| Depression | Phq9_1_score | Categorical: PHI 9 <11 or ≥11 |
| WHO clinical stage | es_23 | Dichotomous: stage 1 - 4 |
| Viral load suppression (<1000 cop-ies/mL) | lab_vls_bl and lab_vl_bl | Dicotomous: Viral load not detectable or below 1000 copies/mL) |
| CD4 cell count | lab_cd4 | Continuous: cell count |
| ART regimen | es_18 | Dichotomous: Efavirenz or other |
| Comprehensive HIV knowledge | b_hiv_3 – b_hiv_7 | Categorical: all five HIV knowledge questions correctly answered? Yes/no |
| Comprehensive ART knowledge | b_art_1 – b_art_4 | Categorical: all four ART knowledge questions correctly answered? Yes/no |
| HIV status disclosure to family or a friend | b_hiv_1 | Categorical: yes, no, refused/don't know |
| Household Food Insecurity scale (HFIS) score | b_food_1 – b_food_14 | Continuous: score |
| Medical Outcomes Study Social Support Survey (MOS-SS) score | b_social_1 – b_social_8 | Continuous: score |
| Alcohol use: AUDIT-C score | b_alc_1 – b_alc_3 | Continuous: score |
| Patient–Provider Relationship Scale (PPRS) score | b_hs_1 – b_hs_9 | Continuous: score |
| General health perception | Medical Outcomes Study Short Form Survey Instrument (MOS SF-36) item | Categorical: |
| Perceived effectiveness of ART | MOS SF-36 item: b_hs_9 | Categorical: |
| Attending support group for PLHIV | b_hs_4 | Categorical: yes, no, refused/don't know |
| Health care decision making | b_gender_1 | Categorical: participant or other |

#### 5.6 Adherence and protocol deviations

Adherence to the interventions and protocol deviations will be assessed based on the extent of exposure to the assigned intervention.

Adherence to the interventions will be defined as:

- Intervention arm: receiving at least four friendship bench individual counselling sessions and nurse-led mental health counselling
- Control arm: receiving nurse-led mental health counselling

Major protocol deviations will be defined as:

- Control arm: not receiving nurse-led mental health counselling or receiving one or more friendship bench individual counselling sessions or friendship bench group support
- Intervention arm: receiving less than four friendship bench individual counselling sessions or not receiving nurse-led mental health counselling

Description of how adherence to the intervention will be presented:

- $N_{int}$ allocated to intervention
- $N_{int}$ received nurse-led mental health counselling & at least four friendship bench individual counselling sessions
- $N_{int}$ did not receive received nurse-led mental health counselling or did not receive at least four friendship bench individual counselling sessions (i.e. premature end of study)
- $N_{int}$ received nurse-led mental health counselling
- $N_{int}$ received individual counselling session for each of the six visits
- $N_{int}$ received friendship bench group counselling
- Median (IQR) number of friendship bench group counselling visits attended

- $N_{cont}$ allocated to control
- $N_{cont}$ received nurse-led mental health counselling
- $N_{cont}$ did not receive nurse-led mental health counselling (premature end of study)

#### 5.7 Withdrawal/follow-up

All withdrawals and losses to follow-up will be listed with the time points and reasons (if available).

### 6. Analysis

#### 6.1 Outcome definitions

##### 6.1.1 Primary outcome

The primary outcome is the mean adherence between 2-6 months (i.e. start of month 2 until end of month 6, total of 5 months (1 month = 30 days)).

We will calculate participants' 1-monthly mean adherence scores as the percentage of doses taken per interval. For regimens that have to be taken once daily, a dose is considered as taken if the participant opens the electronic pill bottle within -8 to +16 hours of the reported usual dosing time (es_24). For regimens that have to be taken twice daily, a dose was considered as taken if the participant opens the electronic pill bottle within -4 to +8 hours of the reported usual dosing time for the morning dose (es_25) or evening dose (es_26), respectively. We will consider 1-monthly mean adherence scores of <10% (i.e. <3 bottle openings) as missing data assuming that participants with very few openings did not consistently use their MEMS caps.

Secondary outcomes

- Mean adherence between 1-12 months (at each month, i.e. for each 1-monthly interval)
- Change from baseline in SSQ-14 score at 3 months (i.e. day 90 ± 45 days), 6 months (i.e. day 180 ± 45 days), 9 months (i.e. day 270 ± 45 days) and 12 months (i.e. day 360 - 45 /+ 60 days).
- Change from baseline in PHQ-9 score at 3 months (i.e. day 90 ± 45 days), 6 months (i.e. day 180 ± 45 days), 9 months (i.e. day 270 ± 45 days) and 12 months (i.e. day 360 - 45 + 60 days), the PHQ score will be calculated based on the test manual.
- Viral load suppression (<1000 copies per milliliter) at 6 months (i.e. day 180 ± 90 days) and 12 months (i.e. day 360 ± 90 days), categorical (yes, no, invalid or missing).

##### 6.1.2 Other outcomes of interest

- Common-mental disorders (SSQ-14 ≥9) at 3 months (i.e. day 90 ±45 days), 6 months (i.e. day 180 ±45 days), 9 months (i.e. day 270 ±45 days) and 12 months (i.e. day 360 – 45 /+ 60 days).
- Depression (PHQ-9 ≥11) at 3 months (i.e. day 90 ±45 days), 6 months (i.e. day 180 ±45 days), 9 months (i.e. day 270 ±45 days) and 12 months (i.e. day 360 – 45 /+ 60 days).

##### 6.1.3 Safety outcomes

- Psychiatric hospitalization up to 12 months (i.e. day 360 + 30 days)
- Suicide attempt up to 12 months (i.e. day 360+ 30 days)
- Suicide up to 12 months (i.e. day 360+ 30 days)

Table 4: Derivation of primary and secondary outcomes.

| Outcome | eCRF sheet | Variable | Outcome type |
|---|---|---|---|
| **Primary**: Mean adherence between 2 – 6 months | Continuous data extracted from medAmigo | adh2 – adh6 | Continuous |
| **Secondary**: Mean adherence at month 1 - 12 | Continuous data extracted from medAmigo | adh1 – adh12 | Continuous |

| | | | |
|---|---|---|---|
| **Secondary**: Change in SSQ-14 and PHQ-9 scores | Baseline: ssq14 and phq9<br>Follow-ups: ssq14 and phq9 | ssq14_1_score<br>phq9_1_score<br>ssq14_1_score_v2<br>phq9_1_score_v2 | Continuous |
| **Secondary**: Proportion of virologic suppressed | Follow-up: detectable viral load (or below 1000 copies / ml) or not | lab_vls_6<br>lab_vls_12 | Dichotomous |
| **Other**: Common-mental disorders (SSQ-14) | Follow-ups: ssq14 | ssq14_1_score_v2 | Dichotomous: ≥9 or <9 |
| **Other:** Depression (PHQ-9 ≥11) | Follow-ups: phq9 | phq9_1_score_v2 | Dichotomous: ≥11 or <11 |
| **Safety:** Psychiatric hospitalisation | Any visit after recruitment | rf_1 | Dichotomous: yes |
| **Safety:** Suicide attempt | Any visit after recruitment | rf_2 | Dichotomous: yes |
| **Safety:** Suicide | Any visit after recruitment | sae_type and sae_description | Dichotomous: death and described as suicide |

#### 6.2 Analysis methods

Analyses will be done at the Institute of Social and Preventive Medicine with support of CTU Bern.

##### 6.2.1 Primary analysis

The primary analysis will be based on the intention-to-treat principle using the FAS. Missing data will be handled according to section 6.4.

We will depict the mean adherence at baseline and each month of follow-up for both groups in a graph showing the point estimate as well as a 95% confidence interval. We will use linear mixed effect models to assess the difference in mean adherence. Models include a random intercept and slope on the participant-level to account for correlation of measurements within participants, a random intercept on the cluster-level to account for clustering of individuals in health facilities, an indicator for treatment assignment, an indicator for each month of analysis time, and interactions of treatment assignment and each month of analysis time. The primary outcome (mean adherence between 2 and 6 months) as well as secondary adherence outcomes (mean adherence at each month) will be assessed from these models based on contrasts.

We will also use linear mixed effect models to assess the difference (95% confidence interval) in change from baseline in SSQ-14 and PHQ-9 score. Models include a random intercept and slope for participants, a random intercept on the cluster level, an indicator for treatment assignment, the respective baseline value of either SSQ-14 or PHQ-9, an indicator for month of analysis time, and interactions of treatment assignment and analysis time.

We will use logistic mixed effect models to assess the difference in the proportion of participants with viral load suppression, with common-mental disorders (SSQ-14 ≥9), and with depression (PHQ-9 ≥11) at 6 and 12 months. Models include a random intercept on the cluster level, and an indicator for treatment assignment. We will present results as OR with 95% confidence interval.)

##### 6.2.2 Secondary analyses

In a per-protocol analysis, we will evaluate outcomes as described in 6.2.1 using the per-protocol patient population.

Because cluster-randomization may lack the excellent balancing properties of individual-level randomization, we will adjust models in a secondary intention-to-treat analysis using the FAS. We will adjust models of adherence outcome data for the size of the clinic, age, gender, type of ART regimen, and CD4 at baseline. Other models will be controlled for the size of the clinic, age, and gender.

Moreover, we will analyze all outcomes on the cluster level using aggregated data. Continuous data will be averaged on the cluster level. For binary data, the proportion will be calculated for each cluster. Averaged data will be compared between groups using a non-parametric Wilcoxon rank-sum test.

###### 6.2.3 Sensitivity analyses

In a sensitivity analysis, we will analyze primary and secondary adherence outcomes as count data based on the FAS. We will use a generalized mixed-effects linear model with a negative binomial distribution, a log-link, a random intercept on the cluster level, an indicator for treatment assignment, and an offset for exposure time. The exposure time will be the number of days with available adherence data (i.e. a maximum of 5 months for the primary outcome and a maximum of 30 days for secondary outcomes).

We will consider a further sensitivity analysis using the FAS if there are pronounced imbalances in patient- and cluster-level baseline characteristics that were not already considered in the secondary analysis, by further adjusting models for these imbalanced variables.

If multiple imputation is employed in the primary analysis, we will additionally perform a complete case analysis based on the FAS.

To see whether the adherence is sensible to the time window (-8 to +16 hours), we will do the analysis of the outcomes addressing adherence by considering a dose as taken if the participant opens the electronic pill bottle within -6 to +6 hours of the reported usual dosing time (es_24 or es_25 and es_26).

###### 6.2.4 Subgroup analyses

Any planned subgroup analyses for each outcome including how subgroups are defined

Table 5: Derivation of subgroups

| Subgroup | eCRF visit | Variable | Categorization |
|---|---|---|---|
| Disease severity (PHQ-9 ≥11 vs <11) | Follow-ups: phq9 | phq9_1_score_v2 | Dichotomous: ≥11 or <11 |
| Men vs. women | Recruitment | gender | Binary: male, female |
| Young vs. old (median) | Recruitment | es_2 | Continuous: years |

###### 6.2.5 Additional analyses

We will evaluate the intra-class correlation (ICC) of the primary and all secondary outcomes.

###### 6.2.6 Assessment of statistical assumptions

Model assumptions for continuous data will be checked visually using plots of residuals (residuals vs fitted values, QQ-plot). If model assumptions are violated, transformation of the outcome (e.g. log-transformation), more robust methods (e.g. robust standard errors or robust regression) or analysis of aggregated data on the cluster level will be considered.

##### 6.3 Interim analyses

There is no interim analysis planned.

##### 6.4 Missing data

If there are patients with completely missing outcome data during the whole follow-up, we will use multiple imputation assuming missing data to be missing at random. If there is missing data at some but available data at other time points, we will use mixed-effects repeated measures models instead, which account for partially missing data.

If multiple imputation is indicated, each outcome will be imputed separately. We will consider all baseline variables (see Table 5.5 with patient characteristics), outcome measures at all time-points as well as an indicator for the treatment and an indicator for the cluster as predictors in the imputation models. Variables with more than 50% missing values will not be used for the imputation model. Binary variables with a frequency of less than 5% in one category will be omitted, levels of categorical variables with a frequency of less than 5% in one category will be combined with another level in a sensible way. Continuous variables will be log-transformed if it improves normality (checked by Shapiro-Wilks test and QQ plots). We will use multiple imputation by chained equation. Predictive mean matching will be used for continuous and ordinal variables, logistic regression for binary and multinomial regression for categorical variables. In total, fifty imputed data sets will be generated, which will be analyzed using Rubin's rules (Rubin 1987).

##### 6.5 Safety evaluation

Safety outcomes will be summarized descriptively, showing the number and proportion with a 95% Wilson confidence interval separate for both groups.

##### 6.6 Statistical software

All statistical analyses will be done in Stata 15.0 (or a subsequent version).

##### 6.7 Quality control

A senior statistician from CTU Bern will reproduce the primary intention-to-treat analysis of the primary and secondary outcomes. Analyses will be based on aggregated 1-monthly MEMS adherence data and clinical data exported from the database system, i.e. multiple imputation will also be reproduced. If results deviate from the original analysis, the reason for the difference will be determined and a consensus must be reached.

All other analyses will be quality-controlled by the senior statistician performing a review of the statistical report.

### 7. Changes from the protocol

The SAP is consistent with principle features of the statistical methods described in the protocol. Any deviation from the protocol is detailed hereunder with reason.

Table 6: Changes from protocol

| Header | Change | Reason |
|---|---|---|
| 5.2 Secondary outcomes | The outcomes 'change from baseline in SSQ-14 score and PHQ-9 score at 3 months and 9 months' were added. | These questionnaires were also collected at month 3 and 9 (besides 6 and 12) and will therefore also be evaluated. |
| 5.2 Secondary outcomes | The outcome 'change from baseline in proportion of participants with a viral load suppression (<20 copies per milliliter) at 6 months and 12 months' was formulated at the patient level. Moreover the threshold was adapted to <1000 copies. | The detection limit is 1000 copies and collected accordingly in the eCRF. |
| 11.3.2 Primary analysis | We deleted the linear mixed effect model for the change from baseline in the proportion of participants with viral load suppression at 12 month. | This population-level outcome cannot be analyzed at the patient level. |
| 11.3.4 Safety analysis | We will calculate the number and proportion of patients with safety outcomes, not incidence rates and rate ratios. | We only expect few safety outcomes on the patient level and overall and therefore decided to present simple descriptive statistics. |

#### 8. References

European Medicines Agency, Committee for Human Medicinal Products; Draft ICH E9 (R1) addendum on estimands and sensitivity analysis in clinical trials to the guideline on statistical principles for clinical trials, step 2b - Revision 1. 30 August 2017.

SOP Data preparation and data analysis, CS_STA_SOP_05, version 04, 11.09.2018

SOP Statistical data validation, CS_STA_SOP_02, version 03, 11.09.2018

StataCorp. 2017. Stata Statistical Software: Release 15. College Station, TX: StataCorp LLC

Rubin, DB (1987). Multiple Imputation for Nonresponse in Surveys. New York: J Wiley & Sons. 1987
