## Supplement 3 for "Effect of the Friendship Bench intervention on antiretroviral therapy outcomes and mental health symptoms in rural Zimbabwe: A cluster randomized trial"

**eMethods** Additional information on standard care, randomization, adherence measures, and statistical analyses

### Enhanced adherence counselling guidelines

Enhanced adherence counselling sessions were conducted by antiretroviral therapy providers who were not part of the study team. Counselling sessions were conducted according to the Operational and Service Delivery Manual for the Prevention, Care and Treatment of HIV in Zimbabwe.[1] The first session consists of nine steps: viral load education review, discussion of the patient's reason for a high viral load, review of the patient's medication dosing schedule, planning for storing of medications, developing patient's motivation card, discussion of patient's support system, planning for substance use, solving issues related to getting to clinic appointments, and finally to plan the next steps (Supplement 4, pp. 78-81). The second session is held one month after the first session and builds on the content discussed in the first session. The session has the following objectives: identifying any difficulties with plans and solving issues with the plan, discussions on how to learn from mistakes, reviewing and attending to any possible referrals that the patient may have gone to, discuss on how to handle travel while on medication, and finally to plan the next steps (Supplement 4, pp. 82-83). Further counselling sessions will be planned as needed.

### Randomization

Randomization was stratified by the size of the health facility (<280, 280-1000, >1000 retained ART patients). We used block randomization within each stratum, with blocks consisting of two health facilities, to balance the size of the groups. Randomization was done by a statistician not part of the study team.

### Standard operating procedures for follow-up and referral of suicidal or psychotic participants

We used a red flag system to alert us of the possible occurrence of self-harm, suicide, or psychosis. A red flag was present when a participant had responded "yes" to item 5 (hallucinations) or item 11 (suicidality) of the SSQ-14. Participants were assessed for a red flag at every visit by the research assistant, during nurse-led counselling by the nurse and the intervention visits by lay health workers. Identified red flags were referred to the nurse in charge at the clinic, who assessed and referred to the medical officer at the provincial psychiatric unit if needed for further management.

### Adherence measures

We used electronic pill caps (Medication Event Monitoring System [MEMS], AARDEX, Sion, Switzerland) to measure adherence during follow-up. We considered a dose missing if no bottle opening was recorded on a particular day. This definition deviated from the statistical analysis plan, which defined a dose as missing if no bottle opening was recorded within a time window of 8 hours before and 16 hours of the designated dosing time. This deviation was necessary because, in some clusters, in about one-third of participants, the reported designated dosing time was incorrectly recorded as it deviated by 6-12 hours from the observed typical dosing time.

We assessed self-reported baseline adherence using a 30-day recall based on the following item from the AIDS Indicator Survey.[2]

“People sometimes forget to take all their ARVs every day. In the last 30 days, how many days have you missed taking any of your ARV pills?”

**Statistical models**

We used linear mixed-effects models to assess the difference in mean adherence. Models included a random intercept and slope at the participant level to account for the correlation of measurements within participants. A random intercept accounted for the clustering of individuals in health facilities, and indicators defined treatment assignment, the month of analysis time, and interactions between the two. We estimated marginal odds ratios for the difference in the proportion of participants with viral suppression at 6 and 12 months using logistic mixed-effect models. We conducted prespecified adjusted analyses of mean adherence and viral suppression, controlling for facility size, age, and sex. In post hoc sensitivity analyses, we adjusted for facility size, age, sex, self-reported baseline adherence, baseline SSQ-14 score, ART regimen, PHQ-9 score, WHO clinical stage, CD4 cell count, viral suppression, audit score, MOS-SS score, and travel cost. We also assessed the difference in change from baseline in SSQ-14 and PHQ-9 scores.

We used logistic mixed-effect models to assess the difference in the proportion of participants with common-mental disorders (SSQ-14 >9) and with depression (PHQ-9>11) at 3, 6, 9, and 12 months. Models include a random intercept at the cluster level and an indicator for treatment assignment. We controlled for facility size, age, and sex in prespecified adjusted analyses. We did a posthoc sensitivity analysis adjusting for facility size, age, sex, baseline SSQ-14 or PHQ-9 score, WHO clinical stage, CD4 cell count, viral suppression, audit score, and MOS-SS score. We repeated analyses of primary and secondary outcomes using a per-protocol analysis, excluding participants who did not receive the allocated intervention and those with missing adherence data between months 2 to 6.

**Multiple imputation models**

Missing data were imputed using multiple imputation by chained equations [3]. We imputed the four outcomes separately. For adherence, imputation models included monthly mean adherence scores for all time points, an indicator for study facility, age, sex (male or female), marital status (married/living together, or single/widowed/divorced/separated), education (primary or secondary), the baseline SSQ-14 and PHQ-9 scores, the baseline CD4 cell count, viral suppression, WHO clinical stage, ART regimen (NNRTI-based, PI-/II-based), the self-reported baseline adherence score, the time since ART initiation, the HFIS, MOS-SS, and AUDIT-C scores, comprehensive ART knowledge (yes or no) and transportation cost to the clinic (<2 USD, 2-5 USD, >5USD).

For viral suppression, imputation models included viral suppression at all time points, an indicator for treatment assignment, age, sex (male or female), marital status (married/living together, or single/widowed/divorced/separated), education (primary or secondary), the baseline SSQ-14 and PHQ-9 scores, the baseline CD4 cell count, WHO clinical stage, the self-reported baseline adherence score, the ART regimen (NNRTI-based, PI-/II-based), the MOS-SS score, and the AUDIT-C score.

Imputation models for SSQ-9 scores included SSQ-9 scores at all time points, an indicator for study facility, age, sex (male or female), marital status (married/living together, or single/widowed/divorced/separated), education (primary or secondary), the baseline PHQ-9 scores, the baseline CD4 cell count, viral suppression, WHO clinical stage, the self-reported baseline adherence score, the time since ART initiation, the HFIS, MOS-SS, and AUDIT-C scores.

Imputation models for PHQ-9 scores included PHQ-9 scores at all time points, an indicator for study facility, age, sex (male or female), marital status (married/living together, or single/widowed/divorced/separated), education (primary or secondary), the baseline SSQ-14 scores, the baseline CD4 cell count, viral suppression, WHO clinical stage, the self-reported baseline adherence score, the time since ART initiation, the HFIS, MOS-SS, and AUDIT-C scores.

**References**

1. AIDS & TB Programme Ministry of Health and Child Care Zimbabwe. *Operational and Service Delivery Manual for the Prevention, Care and Treatment of HIV in Zimbabwe*.; 2017.

2. AIS. AIDS Indicator Survey (AIS). Published 2017. Accessed January 21, 2021. https://dhsprogram.com/What-We-Do/Survey-Types/AIS.cfm

3. Rubin DBD. *Multiple Imputation for Nonresponse in Surveys*. (Rubin DB, ed.). John Wiley & Sons, Inc.; 1987. doi:10.1002/9780470316696

**eTable 1** Effect of the Friendship Bench intervention on adherence, viral load and mental health: results from prespecified and posthoc sensitivity intention to treat analyses

| | Prespecified analyses | | | | Sensitivity analyses (post hoc) | |
|---|---|---|---|---|---|---|
| **Outcome** | **Unadjusted mean difference (95% CI)** | **p-value** | **Adjusted mean difference (95% CI)** | **p-value** | **Adjusted mean difference (95% CI)** | **p-value** |
| MEMS adherence | | | | | | |
| Months 2-6 | 1.93 (-1.20 to 5.06) | 0.2276 | 1.79 (-1.71 to 5.29) | 0.3160 | 2.05 (-1.91 to 6.01) | 0.3098 |
| Months 1-12 | 0.79 (-2.14 to 3.71) | 0.5969 | 0.64 (-2.67 to 3.94) | 0.7056 | 0.90 (-2.89 to 4.68) | 0.6417 |
| **Outcome** | **Unadjusted mean difference in change from baseline (95% CI)** | **p-value** | **Adjusted mean difference in change from baseline (95% CI)** | **p-value** | **Adjusted mean difference in change from baseline (95% CI)** | **p-value** |
| SSQ-14 score | | | | | | |
| Month 3 | -1.65 (-3.07 to -0.24) | 0.0219 | -1.65 (-3.13 to -0.16) | 0.0295 | -1.60 (-3.08 to -0.11) | 0.0347 |
| Month 6 | -1.57 (-2.98 to -0.15) | 0.0302 | -1.56 (-3.05 to -0.07) | 0.0396 | -1.51 (-3.00 to -0.02) | 0.0466 |
| Month 9 | -1.63 (-3.05 to -0.22) | 0.0233 | -1.63 (-3.11 to -0.15) | 0.0313 | -1.58 (-3.06 to -0.09) | 0.0374 |
| Month 12 | -0.78 (-2.19 to 0.63) | 0.2799 | -0.77 (-2.25 to 0.71) | 0.3068 | -0.72 (-2.20 to 0.76) | 0.3399 |
| PHQ-9 score | | | | | | |
| Month 3 | -0.35 (-1.68 to 0.99) | 0.6130 | -0.49 (-1.82 to 0.85) | 0.4734 | -0.47 (-1.82 to 0.88) | 0.4963 |
| Month 6 | 0.01 (-1.33 to 1.34) | 0.9926 | -0.14 (-1.47 to 1.20) | 0.8412 | -0.12 (-1.47 to 1.23) | 0.8633 |
| Month 9 | -0.04 (-1.39 to 1.30) | 0.9484 | -0.19 (-1.53 to 1.15) | 0.7847 | -0.17 (-1.52 to 1.18) | 0.8062 |
| Month 12 | 0.74 (-0.60 to 2.08) | 0.2778 | 0.60 (-0.74 to 1.93) | 0.3792 | 0.62 (-0.73 to 1.96) | 0.3690 |
| **Outcome** | **Unadjusted odds ratio (95% CI)** | **p-value** | **Adjusted odds ratio (95% CI)** | **p-value** | **Adjusted odds ratio (95% CI)** | **p-value** |
| SSQ-14 ≥9 | | | | | | |
| Month 3 | 0.38 (0.09 to 1.59) | 0.1857 | 0.28 (0.07 to 1.12) | 0.0720 | 0.28 (0.07 to 1.09) | 0.0668 |
| Month 6 | 0.41 (0.09 to 1.83) | 0.2453 | 0.30 (0.07 to 1.30) | 0.1085 | 0.30 (0.07 to 1.27) | 0.1026 |
| Month 9 | 0.34 (0.07 to 1.59) | 0.1701 | 0.25 (0.05 to 1.13) | 0.0716 | 0.25 (0.06 to 1.10) | 0.0668 |
| Month 12 | 1.54 (0.33 to 7.15) | 0.5778 | 1.12 (0.25 to 5.01) | 0.8797 | 1.14 (0.26 to 4.88) | 0.8647 |
| PHQ-9 ≥11 | | | | | | |
| Month 3 | 3.24 (0.60 to 17.46) | 0.1709 | 2.16 (0.47 to 9.98) | 0.3260 | 1.86 (0.49 to 7.05) | 0.3620 |
| Month 6 | 3.17 (0.47 to 21.37) | 0.2369 | 2.10 (0.35 to 12.47) | 0.4137 | 1.84 (0.37 to 9.03) | 0.4524 |
| Month 9 | 3.84 (0.49 to 30.30) | 0.2014 | 2.55 (0.37 to 17.74) | 0.3445 | 2.24 (0.39 to 13.04) | 0.3673 |
| Month 12 | 3.11 (0.47 to 20.49) | 0.2383 | 2.06 (0.36 to 11.85) | 0.4164 | 1.81 (0.38 to 8.57) | 0.4568 |
| Viral load <1000 copies/mL | | | | | | |
| Month 6 | 2.20 (0.79 to 6.14) | 0.1309 | 2.26 (0.79 to 6.45) | 0.1274 | 2.93 (0.73 to 11.79) | 0.1297 |
| Month 12 | 1.60 (0.42 to 6.05) | 0.4850 | 1.75 (0.47 to 6.49) | 0.3989 | 3.41 (0.40 to 29.30) | 0.2603 |

MEMS=Medication Event Monitoring System, SSQ= Shona Symptoms Questionnaire, PHQ=Patient Health Questionnaire, CI=confidence interval

**eTable 2** Effect of the Friendship Bench intervention on adherence, viral load and mental health: results from prespecified and posthoc sensitivity per protocol analyses

| | Prespecified analyses | | | | Sensitivity analyses (post hoc) | |
|---|---|---|---|---|---|---|
| **Outcome** | **Unadjusted mean difference (95% CI)** | **p-value** | **Adjusted mean difference (95% CI)** | **p-value** | **Adjusted mean difference (95% CI)** | **p-value** |
| MEMS adherence | | | | | | |
| Months 2-6 | 2.34 (-0.81 to 5.50) | 0.1458 | 1.97 (-1.53 to 5.47) | 0.2701 | 2.23 (-1.80 to 6.27) | 0.2780 |
| Months 1-12 | 1.35 (-1.60 to 4.30) | 0.3699 | 0.95 (-2.37 to 4.27) | 0.5739 | 1.22 (-2.63 to 5.07) | 0.5358 |
| **Outcome** | **Unadjusted mean difference in change from baseline (95% CI)** | **p-value** | **Adjusted mean difference in change from baseline (95% CI)** | **p-value** | **Adjusted mean difference in change from baseline (95% CI)** | **p-value** |
| SSQ-14 score | | | | | | |
| Month 3 | -1.72 (-3.13 to -0.31) | 0.0166 | -1.70 (-3.18 to -0.23) | 0.0237 | -1.64 (-3.11 to -0.16) | 0.0300 |
| Month 6 | -1.60 (-3.01 to -0.19) | 0.0259 | -1.58 (-3.06 to -0.11) | 0.0356 | -1.52 (-2.99 to -0.04) | 0.0444 |
| Month 9 | -1.72 (-3.12 to -0.31) | 0.0166 | -1.70 (-3.17 to -0.23) | 0.0238 | -1.63 (-3.11 to -0.15) | 0.0303 |
| Month 12 | -0.85 (-2.25 to 0.56) | 0.2374 | -0.83 (-2.30 to 0.64) | 0.2698 | -0.76 (-2.23 to 0.71) | 0.3118 |
| PHQ-9 score | | | | | | |
| Month 3 | -0.36 (-1.71 to 0.98) | 0.5977 | -0.48 (-1.83 to 0.86) | 0.4793 | -0.47 (-1.82 to 0.89) | 0.5015 |
| Month 6 | 0.02 (-1.34 to 1.37) | 0.9787 | -0.10 (-1.45 to 1.25) | 0.8806 | -0.08 (-1.45 to 1.28) | 0.9027 |
| Month 9 | -0.09 (-1.45 to 1.26) | 0.8927 | -0.22 (-1.57 to 1.14) | 0.7549 | -0.20 (-1.56 to 1.16) | 0.7770 |
| Month 12 | 0.66 (-0.69 to 2.01) | 0.3361 | 0.54 (-0.80 to 1.88) | 0.4312 | 0.56 (-0.79 to 1.91) | 0.4179 |
| **Outcome** | **Unadjusted odds ratio (95% CI)** | **p-value** | **Adjusted odds ratio (95% CI)** | **p-value** | **Adjusted odds ratio (95% CI)** | **p-value** |
| SSQ-14 ≥9 | | | | | | |
| Month 3 | 0.34 (0.08 to 1.43) | 0.1412 | 0.25 (0.06 to 1.05) | 0.0576 | 0.25 (0.06 to 1.01) | 0.0522 |
| Month 6 | 0.42 (0.10 to 1.84) | 0.2500 | 0.31 (0.07 to 1.35) | 0.1187 | 0.31 (0.07 to 1.32) | 0.1128 |
| Month 9 | 0.33 (0.07 to 1.55) | 0.1592 | 0.24 (0.05 to 1.13) | 0.0715 | 0.25 (0.05 to 1.10) | 0.0666 |
| Month 12 | 1.50 (0.32 to 6.93) | 0.6039 | 1.11 (0.24 to 5.00) | 0.8960 | 1.13 (0.26 to 4.88) | 0.8749 |
| PHQ-9 ≥11 | | | | | | |
| Month 3 | 3.21 (0.57 to 18.06) | 0.1864 | 2.21 (0.46 to 10.70) | 0.3235 | 1.98 (0.51 to 7.75) | 0.3249 |
| Month 6 | 3.24 (0.47 to 22.40) | 0.2330 | 2.23 (0.37 to 13.54) | 0.3823 | 2.04 (0.41 to 10.19) | 0.3860 |
| Month 9 | 3.75 (0.46 to 30.90) | 0.2192 | 2.58 (0.35 to 18.82) | 0.3495 | 2.37 (0.39 to 14.30) | 0.3454 |
| Month 12 | 2.68 (0.39 to 18.28) | 0.3140 | 1.85 (0.31 to 10.95) | 0.4997 | 1.69 (0.34 to 8.33) | 0.5213 |
| Viral load <1000 copies/mL | | | | | | |
| Month 6 | 2.58 (0.89 to 7.53) | 0.0817 | 2.52 (0.85 to 7.45) | 0.0938 | 2.89 (0.72 to 11.65) | 0.1356 |
| Month 12 | 1.99 (0.46 to 8.53) | 0.3529 | 2.05 (0.49 to 8.65) | 0.3268 | 3.34 (0.36 to 30.70) | 0.2850 |

MEMS=Medication Event Monitoring System, SSQ= Shona Symptoms Questionnaire, PHQ=Patient Health Questionnaire, CI=confidence interval

**eTable 3** Number of participants with safety-relevant outcomes by trial arm

| | **Friendship Bench** N=244 | **Standard of Care** N=272 |
|---|---|---|
| Reported actual or attempted self-harm | 11 (4.5%) | 5 (1.8%) |
| Before or on baseline | 9 (3.7%) | 5 (1.8%) |
| Incident event after baseline | 2 (0.8%) | 0 (0.0%) |
| Psychiatric hospitalizations | 0 (0.0%) | 0 (0.0%) |
| Deaths | | |
| Unrelated to study procedures and intervention | 5 (2.0%) | 1 (0.4%) |
| Related to study procedures or intervention | 0 (0.0%) | 0 (0.0%) |
| Suspected or reported suicide | 0 (0.0%) | 0 (0.0%) |

**eFigure 1** Forest plot of available evidence of the effect of the Friendship Bench intervention on SSQ-14 scores, PHQ-9 scores, and viral suppression from three trials

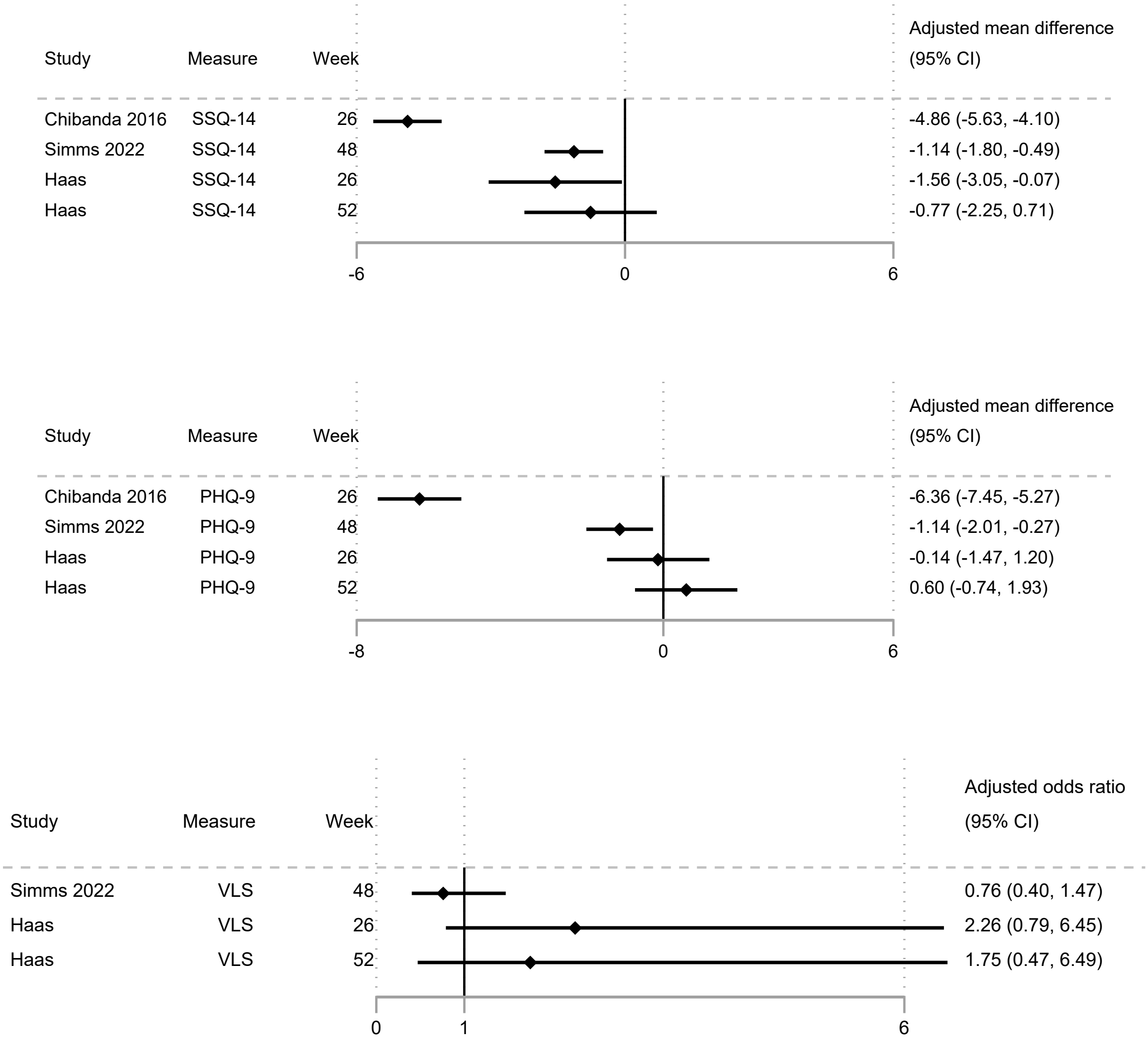


SSQ=Shona Symptoms Questionnaire, PHQ= Patient Health Questionnaire, VLS=viral suppression
