## Supplement 4 for "Effect of the Friendship Bench intervention on antiretroviral therapy outcomes and mental health symptoms in rural Zimbabwe: A cluster randomized trial"

### OPERATIONAL AND SERVICE DELIVERY MANUAL

### FOR THE PREVENTION, CARE AND TREATMENT OF HIV IN ZIMBABWE

**AIDS & TB Programme**

**Ministry of Health and Child Care, Zimbabwe**

### Operational and Service Delivery Manual for the Prevention, Care and Treatment of HIV in Zimbabwe

AIDS & TB Programme - Ministry of Health and Child Care, Zimbabwe
February 2017

### CONTENT

Abbreviations 2
Background and Rationale for the Manual 3
Manual Development Process 3
How to Use this Manual 4

#### Chapter 1: Core Elements of HIV Service Delivery 6

1.1 The minimum package for HIV prevention, care and treatment services 7
1.2 Human resources 16
1.3 Decentralisation of HIV prevention, care and treatment services 22
1.4 Integration of services 24

#### Chapter 2: Differentiated Service Delivery 28

2.1 Introduction to differentiated service delivery 29
2.2 Differentiated HIV testing services 31
2.3 Differentiated HIV prevention strategies 38
2.4 Differentiated ART initiation 40
2.5 Differentiated ART delivery 50
2.6 Differentiated ART delivery for specific sub-populations 68
2.7 Differentiated ART delivery for clients with high viral load 74

#### Chapter 3: Pharmacy, Laboratory and Strategic Information 88

3.1 Pharmacy 89
3.2 Laboratory 91
3.3 Monitoring and evaluation 93
3.4 Quality improvement and implementation research 96

#### Appendices 98

Appendix 1: AIDS and TB referral form 99
Appendix 2: HIV re-testing. Frequently asked questions 100
Appendix 3: Shona symptom questionnaire for the detection of depression and anxiety 102
Appendix 4: Questionnaire to support decision making for differentiated service delivery 103
Appendix 5: SOPs for documentation at clinical and refill visits 106
Appendix 6: M and E forms for ART clubs and CARGs 110

#### Acknowledgements 113

### ABBREVIATIONS

| | |
|---|---|
| ANC | Antenatal care |
| ART | Antiretroviral therapy |
| CARG | Community ART refill group |
| CBO | Community-based organisation |
| CHW | Community health worker |
| CICT | Client-initiated counselling and testing |
| CRAG | Cryptococcal antigen |
| CSF | Cerebral spinal fluid |
| DBS | Dried blood spot |
| DHE | District Health Executive |
| DMO | District medical officer |
| DNO | District nursing officer |
| EAC | Enhanced adherence counselling |
| EDLIZ | Essential Drug List of Zimbabwe |
| EHT | Environmental health technician |
| EID | Early infant diagnosis |
| EPI | Expanded programme of immunisation |
| HIVST | HIV self-testing |
| HTS | HIV testing services |
| ICF | Intensified case finding |
| INH | Isoniazid |
| IPD | Inpatient department |
| M&E | Monitoring and evaluation |
| MNCH | Maternal and child health |
| MoHCC | Ministry of Health and Child Care |
| MSM | Men who have sex with men |
| NCD | Non-communicable disease |
| NVP | Nevirapine |
| OI | Opportunistic infections |
| OPD | Outpatient department |
| PC | Primary counsellor |
| PDSA | Plan-do-study-act |
| PEP | Post-exposure prophylaxis |
| PHE | Provincial Health Executive |
| PITC | Provider-initiated testing and counselling |
| PLHIV | People living with HIV |
| PMTCT | Prevention of mother-to-child transmission |
| PNC | Postnatal care |
| POC | Point of care |
| PrEP | Pre-exposure prophylaxis |
| QC | Quality control |
| QI | Quality improvement |
| SOPs | Standard operating procedures |
| SRH | Sexual and reproductive health |
| STI | Sexually transmitted infection |
| TDF | Tenofovir |
| VIAC | Visual inspection with acetic acid and cervicography |
| VMMC | Voluntary medical male circumcision |
| UNAIDS | Joint United Nations Programme on HIV/AIDS |
| WHO | World Health Organization |
| ZADS | Zimbabwe ART Distribution System |
| ZAPS | Zimbabwe Assisted Pull Systems |

### Background and Rationale for the Manual

By June 2016, 18.2 million people were receiving antiretroviral therapy (ART) in low- and middle-income countries. This increase in the number of clients on ART over the past decade has been achieved through political commitment, community mobilisation and significant domestic and international financial support.

Zimbabwe has been one of the countries worst affected by the HIV epidemic in sub-Saharan Africa. The latest estimates reveal a national HIV prevalence (15-49 years) of 14.7%. About 1.2 million people are estimated to be living with HIV, with 944,614 accessing ART by the end of September 2016. Prevention of mother-to-child transmission (PMTCT) coverage is estimated at 82% of those in need.

Achieving coverage of HIV testing and initiation of ART alone is no longer enough. Ensuring adequate linkages and quality monitoring is now required, with the aim being to reach the 90-90-90 goals set by UNAIDS: 90% of those infected with HIV are identified; 90% of those identified access appropriate antiretroviral therapy (ART); and 90% of those on ART achieve virological suppression.

In December 2016, the Ministry of Health and Child Care (MoHCC) launched its new *Guidelines for Antiretroviral Therapy for the Prevention and Treatment of HIV in Zimbabwe*. These guidelines mark the launch of exciting new initiatives, such as "Treat All", ART for all HIV-positive clients regardless of the degree of immunosuppression, HIV self-testing (HIVST) and pre-exposure prophylaxis (PrEP). In order to achieve the 90-90-90 goals while maintaining a quality service for people living with HIV, innovative programmatic strategies are needed along with active engagement of the community.

To accompany the 2016 clinical guidelines, which outline the **"what to do"**, this *Operational and Service Delivery Manual* is aimed at giving guidance on the **"how to do it" with the aim of increasing retention at all steps of the cascade.** This is the second edition of the manual originally developed in 2014.

Chapter 1 outlines the best practices identified for the organisation of service delivery, defining the minimum package of care, scope of practice, training and mentorship strategies for human resources, decentralisation and integration of services.

Chapter 2 introduces the concept of differentiated service delivery and describes the range of differentiated testing, prevention, initiation and ART delivery strategies that will be considered in Zimbabwe. For each strategy, the four building blocks addressing the "when, where, who and what" are described along with special considerations for specific sub-populations, such as children and adolescents, pregnant and breastfeeding women and key populations.

Chapter 3 provides key messages regarding the essential support services of pharmacy and laboratory and highlights the importance of monitoring and evaluation accompanied by quality improvement projects for the successful functioning of any HIV prevention, care and treatment programme.

### Manual Development Process

The first edition of the Operational and Service Delivery Manual was developed in 2014. The development process was spearheaded by a steering committee composed of representatives of the MoHCC and key stakeholders under the leadership of the Deputy Director for HIV and AIDS of the MoHCC. For the 2016 revision, the following steps were followed:

- Desk review of relevant documents, including background literature and systematic reviews, international guidance, national guidelines, policies and programme reviews
- Discussion and interviews with key stakeholders within the MoHCC, partners, service providers and service users
- Site visits
- Two-day stakeholders' consultative workshop
- Pre-testing of the standard operating procedures.

### How to Use this Manual

This manual is for doctors, clinical officers, nurses, counsellors, pharmacists, health information officers, health promotion officers, community health workers and community-based organisations (CBOs) providing HIV prevention, care and treatment services to children, adolescents and adults (including pregnant and breastfeeding women).

Each section describes a brief background to the topic and highlights key messages and reference materials that should be referred to in combination with the text. Throughout the text, the icons shown in Table 1 point the reader towards considerations for specific sub-populations.

Tables 2-5 highlight the pages where special mention of issues for subpopulations, children and adolescents, pregnant/ breastfeeding women, and key populations are referenced.

#### Table 1 : Key for icons

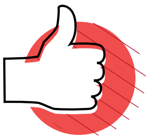

**Key Messages**

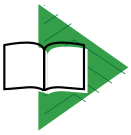

**Reference materials**

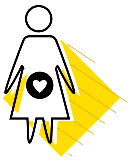

**Special Considerations for Pregnant and Breastfeeding Women**

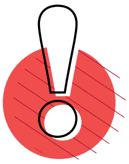

**Very important : Must Implement**

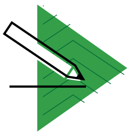

**Resource material (tool or form)**

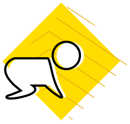

**Special Considerations for Children**

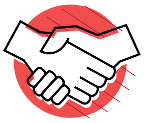

**Opportunity for Community and Facility Linkage**

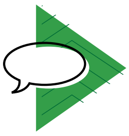

**Counselling activity (Primary counsellor or Nurse)**

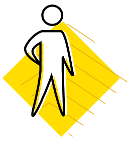

**Special Considerations for Adolescents**

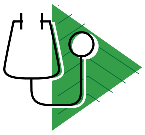

**Activity performed by a clinician**

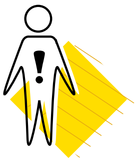

**Special Considerations for Key Populations**

#### Table 2 : Special considerations for children (0–9 years)

| PAGE | TOPIC |
|---|---|
| 33 | HIV testing services |
| 43 | ART Initiation |
| 47-48, 69 | ART Follow Up |
| 85 | Treatment Failure |

#### Table 3 : Special considerations for adolescents (10–19 years)

| PAGE | TOPIC |
|---|---|
| 33 | HIV testing services |
| 43 | ART Initiation |
| 47-48, 69, 72 | ART Follow Up |
| 85 | Treatment Failure |

#### Table 4 : Special considerations for pregnant and breastfeeding women

| PAGE | TOPIC |
|---|---|
| 43 | ART Initiation |
| 47-48, 68 | ART Follow Up |
| 86 | Treatment failure |

#### Table 5 : Special considerations for key populations

| PAGE | TOPIC |
|---|---|
| 35 | HIV testing services |
| 70 | ART Follow Up |

### CHAPTER 1

#### Core elements of HIV service delivery

### 1.1 The minimum package for HIV prevention, care and treatment services

#### 1.1.1 Background

All health facilities in Zimbabwe are expected to provide a minimum package of services for prevention, care and treatment of HIV for children, adolescents and adults (including pregnant and breastfeeding women).

The Zimbabwean health system is a tiered structure, as shown in Figure 1. In addition to the minimum package of services, each successive tier of the health system will have additional responsibilities related to clinical management, mentorship, supportive supervision, pharmacy and laboratory support services and monitoring and evaluation.

##### Figure 1: The Zimbabwean health care system

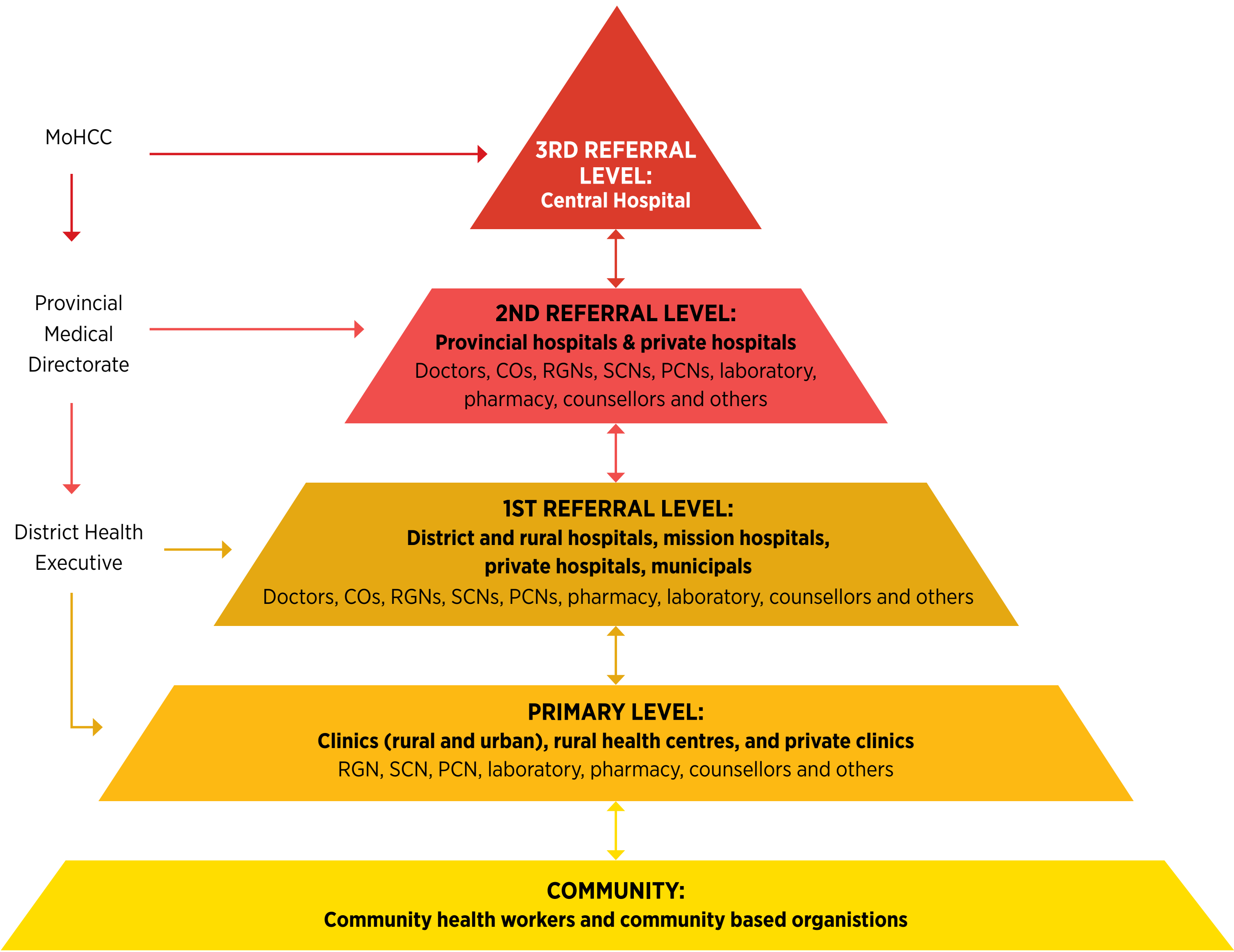

#### 1.1.2 The minimum package for HIV prevention, care and treatment at all facilities

The minimum package of HIV prevention, care and treatment services should be provided at all health facilities from Monday to Friday, 8am-4pm.

**Extended opening hours (early morning, evening or weekend) should be considered during the decision process for differentiating HIV testing and ART service delivery according to the size of the clinic cohort and the needs of specific sub-populations.** Children and adolescents at school will benefit from appointments outside school hours and during school holidays; working adults and at-risk populations may benefit from an early morning clinic at 7-8am or a later clinic at 4-6pm. This could be provided once a week or once a month depending on the cohort size.

##### Table 6: The minimum package of services that should be provided at all tiers of the health care system

| HIV TESTING SERVICES (SEE SECTION 2.2) | |
|---|---|
| Facilities should provide CICT and opt-out PITC for all clients attending the facility (including couples, children and adolescents) at all entry points (OPD, ANC, TB, MNCH/SRH, EPI, IPD) regardless of the purpose of the visit | All facilities should ensure that there are adequate staff members available to provide CICT/PITC (rapid HIV testing and collection of DBS for DNA PCR testing) at all entry points of their facility.<br><br>If there is not sufficient demand to place a dedicated staff member at all entry points, a clear referral system or rota should ensure that there is a trained primary counsellor or nurse available to provide HIV testing services (HTS) for all departments and wards during the day, night and at weekends.<br><br>Re-testing of all HIV-positive diagnoses must be made prior to ART initiation. This should be performed on a different sample and ideally by a different provider.<br><br>Re-testing of HIV-negative clients should be performed according to national recommendations based on the risk of exposure and sub-population (see Job Aide, page 16).<br><br>Facilities should link with their community-based actors to mobilise their communities to attend for HTS. In particular, communities should encourage all pregnant women, children and adolescents and couples to attend for HTS. |
| Index case testing should be provided at the facility and community | Any HIV-positive client is the index case. Family members and all partners of an HIV-positive client should be offered HTS.<br><br>This may be offered :<br>• At the facility (with community-based support to encourage testing)<br>• Where consent is given through community-based index client testing performed by the lay counsellor, nurse or trained community health worker (CHW) or expert client (this may be using HIV self-testing kits)<br>• By offering HIV self-testing kits to the index case |

| | |
|---|---|
| Provider-initiated testing should be incorporated into existing outreach activities | Facilities should incorporate PITC for adults and children (including DBS for DNA PCR) into existing outreach activities.<br>Facilities should link with community-based actors to mobilise the community to attend for HTS on scheduled outreach days. |
| Mobile targeted outreach testing should be organised on a regular basis (at least quarterly) | Facilities should organise targeted mobile outreach testing in order to access those members of the community (men, children and adolescents, artisanal miners and key populations, such as sex workers, MSM, long-distance truck drivers and drug users) who do not regularly visit the health centre. In order to maximise these activities, facilities should analyse their testing data to see which groups are not attending the facility so that they can be targeted during specific outreach testing activities.<br>Facilities should link with community-based actors to mobilise clients to attend for HTS services at planned outreach testing and counselling activities. |
| Role of community cadres (community health workers and expert clients) to perform HTS | Selected cadres will be trained to conduct HTS primarily using HIV self-testing kits. This will be under the supervision of the facility. Linkage between the facility and these community cadres will be essential to maximise the impact of these activities, to ensure quality and data collection. |
| **LINKING CLIENTS TO PREVENTION, CARE AND TREATMENT** | |
| Clients testing negative should be proactively linked to prevention services | Condoms should be distributed alongside HTS.<br>Linkage to voluntary medical male circumcision (VMMC) services should be ensured.<br>Clients at substantial risk of HIV transmission should be assessed for PreP (See Job Aide page 31) |
| Clients testing positive should be proactively linked to the facility ART services for continuing care | If tested positive during outreach or community-based testing, with their consent, the client should be linked to a CHW or another community-based representative working with HIV, who should assist the client to link to appropriate care and treatment.<br>Where mobile testing services are being provided, consider community-based initiation after completing the clinical and psychosocial readiness assessment.<br>POC CD4 should be used during outreach testing activities to identify clients with advanced disease. |

| **PREVENTION (SEE SECTION 2.3)** | |
|---|---|
| Provision of health education on how to prevent HIV transmission | Education on HIV should be regularly included in the facility in daily health talks.<br><br>Community-based organisations must promote prevention of HIV infection through HIV education in the community (e.g., at schools, workplaces, public gatherings, church functions and door-to-door health education). |
| Provision of male and female condoms | Condoms should be easily accessible at the facility, e.g., available in toilets and waiting areas. Condoms should be proactively offered to anyone attending with an STI or for HIV testing services.<br><br>Facilities should link with community health workers, PLHIV, church leaders and CBO members to distribute condoms. Condoms should be available in hotspots, such as beer halls and growth points. |
| Treatment of STIs | All facilities should provide syndromic STI treatment. Where mobile testing services are implemented with a trained nurse within the team, treatment of STIs may be integrated into the package of services. |
| Linkage with VMMC | All facilities should offer or refer clients for VMMC (promote in health talks; have posters of locally available services).<br><br>CHW and community leaders should mobilise their communities to take up VMMC. |
| Post-exposure prophylaxis (PEP) | PEP should be available for all facility staff after an accidental exposure to blood, for anyone presenting after an episode of sexual violence or for anyone assessed to have been exposed to a significant risk of sexual exposure (unprotected sex with known HIV-positive client or high-risk group). |
| Pre-exposure prophylaxis (PrEP) | PrEP should be available in all facilities with trained health care workers. PrEP should be provided to those at substantial risk of HIV infection according to the criteria outlined in the clinical guidelines. PrEP will be introduced in Zimbabwe in a phased manner and guidance will be provided by the MoHCC. |
| **PROVISION OF ART SERVICES (SEE SECTIONS 2.4, 2.5 AND 2.6)** | |
| All facilities should provide the basic package for differentiated ART initiation (see Section 2.4) to all HIV-positive clients | All HIV-positive clients, regardless of age and CD4 count, are eligible to start ART. ART should be initiated as soon as possible after assessing clinical and psychosocial readiness.<br><br>Facilities should provide access to:<br>• Clinical assessment and staging<br>• TB screening, diagnosis and treatment (See Job Aide pages 95-97)<br>• CD4 testing (and other baseline investigations if available)<br>• Cotrimoxazole and TB preventive therapy (IPT)<br>• Cryptococccal antigen screening and fluconazole prophylaxis<br>• Treatment and/or referral for opportunistic infections and cancers<br>• ART adherence preparation and initiation counselling for children, adolescents and adults (including pregnant and breastfeeding women). This should include an ART readiness assessment<br>• Integrated SRH and MCH services, including treatment of STIs and screening for cervical cancer using VIAC<br>• Family planning |

| PROVISION OF ART SERVICES | |
|---|---|
| Facilities should provide a package of differentiated ART delivery for stable clients (see Section 2.5)<br><br>A stable client where there is access to VL<br>• Has no current OIs<br>• Has a VL <1000 copies/ml<br>• Is at least 6 months on their current regimen<br>A stable client where there is no access to VL<br>• Has no current OIs<br>• Has a CD4 >200 cells/mm3<br>• Is at least 6 months on their current regimen<br>Facilities should provide differentiated ART delivery for specific subpopulations (See section 2.6) | All facilities should have an appointment system, and clients should be traced if they do not attend (Section 2.5.2).<br>Facilities should provide the services outlined in the clinical and refill visit details (Section 2.5.3).<br>A 3-month refill of ART and cotrimoxazole should be provided.<br>The choice of refill option available at an individual site should be chosen according to a needs assessment, as outlined in Section 2.5.4. Five options should be considered:<br>• Fast Track: Facility-based individual refill from pharmacy<br>• Facility Club: Facility-based health care worker-led group refill<br>• Outreach: Community individual ART delivery through mobile outreach<br>• CARG: Community-based client-led group refill<br>• Family member refill<br><br>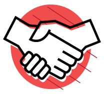 Facilities should link with community health workers and expert clients to strengthen treatment literacy, adherence, raise awareness around options available for ART delivery and to perform defaulter tracing. |
| Facilities should provide the package of differentiated ART delivery for clients with high viral load as outlined in Section 2.7 | Viral load (or CD4 if VL not available) should be used to monitor the client along with clinical assessment. In addition to those who do not attend for ART appointments, defaulter tracing should be performed for anyone whose monitoring tests show signs of treatment failure.<br>Enhanced adherence support (Pages 79-83) for red-flag clients (patients who present with viral load >1000 copies/ml, possible signs or symptoms of treatment failure or attend late for refills) should be provided.<br>Initiation and maintenance of second-line ART should be provided from all facilities with trained health care workers. |

| | |
|---|---|
| **PROVISION OF INTEGRATED TB/HIV SERVICES (SEE SECTION 1.4.2)** | |
| Facilities should provide integrated TB/HIV services as outlined in Section 1.4.2 | All clients with HIV should be screened for TB at every clinical visit and have access to diagnostic services (Xpert MTB/Rif, Culture, CXR).<br><br>HIV-positive clients with no symptoms of TB should be considered for TB preventive therapy according to the national guidelines.<br><br>All TB clients should be screened for HIV.<br><br>If diagnosed with TB and HIV, they should be initiated and followed for both diseases within the same facility, collecting drugs for both TB and ART on the same day.<br><br>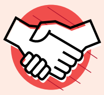 Community health workers and other community-based workers should educate, screen and refer/accompany presumptive TB cases from the community. |
| **PROVISION OF INTEGRATED SRH/MCH AND PMTCT SERVICES (SEE SECTION 1.4.3)** | |
| Facilities should provide integrated ANC/delivery/PNC and PMTCT care as outlined in Section 1.4.3 | PMTCT should be provided as part of MNCH services (antenatally, at delivery and postnatally) so that the woman, child and husband/partner are seen as a family under a one-stop service approach (same nurse, same consultation room, same day). This is known as the family-centred approach.<br><br>Family planning should be available for all HIV-positive clients as a one-stop service.<br><br>Screening for cervical cancer using VIAC should be available within the district.<br><br>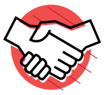 Community health workers and other community-based workers should identify clients who need SRH/MNCH/PMTCT and refer them to the facility. Facilities should link with the community to strengthen treatment literacy education and to perform defaulter tracing of pregnant and breastfeeding women. |
| **PROVISION OF INTEGRATED NCD/HIV SERVICES (SEE SECTION 1.4.4)** | |
| Facilities should provide integrated NCD/HIV services as outlined in Section 1.4.4 | All clients on ART should have their blood pressure and cardiovascular risk assessed annually.<br><br>All clients on ART should be screened for depression and anxiety annually<br><br>Clients with red-flag characteristics (VL >1000 copies/ml, signs of clinical failure, missed appointments) should be screened for symptoms of depression and anxiety. |

| PHARMACY | |
|---|---|
| Facilities are responsible for ensuring a continuous supply of OI and ART medicines (see Section 3.1) | **Access to:**<br>Cotrimoxazole<br>Aciclovir<br>Fluconazole<br>Other essential antibiotics, including treatment of STIs<br>Family planning commodities<br>Adult and paediatric first- and second-line regimens at all facilities<br>Adult and paediatric third-line regimen available after review from tertiary institutions<br>Nevirapine and zidovudine syrup for exposed baby management<br>Anti-TB medicines |
| **LABORATORY** | |
| Facilities should make all efforts to have access to the test kits listed. Lack of access to these investigations does not mean ART therapy should be delayed.<br>Facilities should ensure regular stock management to avoid over/under stocking and expiries.<br>Facilities should ensure regular sample transport, e.g., weekly (see Section 3.2.2) | **At primary health facility:**<br>HIV testing kits<br>DBS kits for DNA PCR<br>Pregnancy tests<br>Syphilis rapid tests<br>Hb meters and test strips<br>Glucometers<br>Urine dipstick<br>Specimen tubes for CD4, FBC and biochemistry<br>DBS kits for viral load testing<br>Point-of-care technologies (e.g., cartridges for CD4) where appropriate<br>**At district level, all of above plus:**<br>TB diagnosis (smear or Xpert MTB/Rif)<br>CRAG testing for blood and CSF (at minimum access to Indian ink)<br>Creatinine (TDF use)<br>ALT (NVP use)<br>CD4 (baseline and when clinically indicated)<br>Hepatitis B and C screening<br>VL and EID cartridges for near POC where appropriate<br>**At provincial level, all of above plus:**<br>Viral load testing<br>EID testing<br>**At tertiary level, all of the above plus:**<br>Genotyping<br>TB culture and drug sensitivity testing |
| **INFRASTRUCTURE AND EQUIPMENT REQUIRED** | |
| Facilities should ensure that there is adequate equipment available to offer the minimum package of services | Running water<br>Well-ventilated room; room with confidentiality for counselling<br>BP machines<br>Stethoscopes<br>Torch and otoscope/auroscope<br>Thermometers<br>Height measuring boards/charts, child health cards for 0-5 and weight-for-age and height-for-age charts for older children, MUAC tapes, weight/salter scales<br>Examination couches |

| **MANAGEMENT OF HIV PREVENTION, CARE AND TREATMENT SERVICES** | |
|---|---|
| Each facility should allocate a focal person for HIV prevention care and treatment services | This person is responsible for ensuring quality provision of HIV prevention, care and treatment services. |
| Current guidelines and Job Aides, as referenced in this document, should be available | Essential in every clinic:<br>Guidelines for Antiretroviral Therapy for the Prevention and Treatment of HIV in Zimbabwe<br>Operational and Service Delivery Manual for the Prevention, Care and Treatment of HIV in Zimbabwe<br>Job Aide for the Prevention, Care and Treatment of HIV in Zimbabwe<br>National Guidelines on HIV Testing and Counselling<br>National Guidelines on HIV Testing for Children and Adolescents<br>Essential Drug List of Zimbabwe (EDLIZ)<br>TB guidelines |
| Organisation of clinic and community meetings | Facilities should organise regular case discussion meetings to review difficult/failing cases (weekly or monthly depending on case load). Where there is only one nurse, regular case discussions must be organised with the district mentoring team.<br>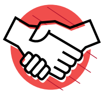 Facilities should organise a regular health centre committee meeting (at least quarterly) with community leaders and community-based organisations engaged in HIV prevention care and treatment activities.<br>Facilities should organise regular meetings (monthly) between the facility and community health workers. |
| Data collection, analysis and reporting (see Section 3.3) | All HIV-related activities, (testing and counselling, ART, PMTCT, TB/HIV, STI management) must be reported according to the national monitoring and evaluation (M&E) standard reporting system. Each level of the health system will have its individual responsibility for reporting and analysis. Data from community-based activities should be reported under their respective facility.<br>Analysis of facility data should be utilised for continuous quality improvement and for decision making at facility level (see Section 3.4). |

##### 1.1.3 Additional responsibilities for district hospitals

In addition to the minimum package of services, the district must also support the primary care clinics that are in its catchment area. The district must ensure there is:

- A referral system for sick/complicated clients to be linked to the hospital for further investigation and management
- Treatment for cryptococcal meningitis as stated in the *Guidelines for Antiretroviral Therapy for the Prevention and Treatment of HIV in Zimbabwe*
- Continuation of any treatments initiated at tertiary level, for example, chemotherapy treatment for Kaposi's sarcoma
- Screening and treatment for cervical cancer
- A pharmacist or pharmacy technician who can support the clinics to strengthen supply chain management and respond in a timely manner to any emergency medicine shortages at the clinics
- A laboratory that provides the basic package of investigations as outlined in the minimum package. Maintenance contracts for all machines (haematology, biochemistry and CD4), along with quality assurance

mechanisms, must be in place. The district laboratory must also ensure that adequate quality assurance (internal and external) is in place for tests that are being performed as point of care at the clinic (Section 3.2)

- A radiology department that provides access to X-ray and, ideally, ultrasound
- A district **multi-disciplinary** mentorship team that visits each clinic (see Section 1.2.3). In addition, telephone and other electronic support should be available to the clinic nurses from the district HIV prevention, care and treatment mentors to assist in the management of complicated clients and cases of treatment failure
- As part of the district clinical mentorship planning, provision of a clinical attachment system for those nurses identified to be in need of clinical skills enhancement in HIV prevention, care and treatment
- A district human resources management plan to ensure adequate trained health care workers are available across all clinics
- Regular assessment of staff training needs at district and clinic level in order to provide the minimum HIV prevention, care and treatment package of services
- Supportive supervision. These visits should be performed by the District Health Executive (DHE) team, but should also include co-opted members as per need, such as the laboratory scientist, district health information officer, OI sister in charge, nutritionist and district AIDS coordinator. Each clinic within a district should be visited quarterly. Following a supervision visit, an action plan should be developed for each site
- District-wide coordination of partners and HIV service delivery organisations
- District-wide coordination of community engagement with HIV activities in line with local health needs
- Consolidation of the monthly reports from all facilities and transmission of data to provincial level.

#### 1.1.4 Additional responsibilities for provincial hospitals

In addition to the minimum package of services, provincial hospitals should:

- Accept referrals from the district facilities to diagnose and treat more complicated clinical cases, including the management of drug-resistant tuberculosis (DRTB)
- Provide laboratory tests that are not available at the district, e.g., viral load
- Provide additional radiological testing that is not available at the district
- Consolidate provincial data for reporting to national level.

#### 1.1.5 Additional responsibilities for tertiary hospitals

These hospitals should function as centres of excellence. In addition to the minimum package of services for their catchment area, they should:

- Accept referrals from district and provincial medical staff to manage complicated cases including cases of second-line treatment failure
- Act as the referral centre for suspected Kaposi's sarcoma and initiate chemotherapy treatment
- Provide laboratory tests that are not available at the district and provincial sites, such as resistance testing, TB culture and drug sensitivity testing

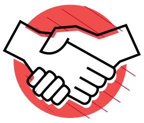

- Provide additional radiological testing that is not available at the district or provincial sites, such as CT and MRI scanning.

#### 1.1.6 The role of the community

It is essential to engage the community in policy development, programming, resource mobilisation, implementation and evaluation of services. Across the HIV care and treatment cascade, community engagement provides the possibility of increasing uptake of HIV testing and treatment and enhancing retention and successful adherence to ART.

To achieve this, there must be improved coordination and monitoring of community-based activities and greater linkage with services provided at the facility. Facility managers, community nurses and district coordinators must ensure that they guide their communities in the activities they choose in order to maximise the health benefits at each step of the cascade.

Developing the capacity of existing community-based workers using harmonised training materials in line with the *Guidelines for Antiretroviral Therapy for the Prevention and Treatment of HIV in Zimbabwe and the Operational and Service Delivery Manual* is the first step. Parallel to this, there is an urgent need to scale up the capacity of community members and volunteers to support community mobilisation and treatment literacy activities.

Community-based organisations should also advocate for increased political commitment and financial support for HIV programming and act as a watchdog, using community monitoring systems to report on the quality of services provided and critical issues, such as drugs supply shortages or stock outs.

### 1.2 Human resources

#### 1.2.1 Background

Decentralisation and integration of HIV prevention, care and treatment activities across all levels of the health system has required a critical review of the roles and responsibilities of health care workers. With the further scale up of ART demanded by the 2016 *Guidelines for Antiretroviral Therapy for the Prevention and Treatment of HIV in Zimbabwe* and the introduction of "Treat All", further review of the tasks and working practices of health workers is required. By reviewing the scope of practice of health workers, not only may access and retention be improved, but it should also allow clinicians (doctors and nurses) to spend more time with clients with more complex medical needs.

Evidence from the literature has demonstrated that there is no difference in clinical outcomes, including mortality or losses to follow up, when nurses initiate or manage people on ART relative to physician-led care. Nurses should therefore be empowered to initiate and follow up children, adolescents and adults (including pregnant and breastfeeding women). Quality of care, however, should be ensured through adequate training, ongoing mentorship, clear indications for referral to higher levels of care, and monitoring and evaluation systems that are utilised for improving client management.

#### 1.2.2 Roles and responsibilities of health care workers

The precise distribution of tasks will depend on the level of the health system, the number of staff available at a facility and the size of the cohort being served. It is the responsibility of the nurse in charge of the clinic or HIV prevention, care and treatment centre to ensure that all tasks have a member of staff clearly identified and responsible for fulfilling it.

Table 7 outlines the scope of practice for different health care workers involved in the provision of HIV prevention, care and treatment services. Each site should use this table to review the scope of practice within their facility and establish responsibilities for each step.

##### District medical officer and district nursing officer

The district medical officer (DMO), along with the district nursing officer (DNO), is responsible for the successful implementation of the minimum package of care for HIV prevention, care and treatment services within their district, including ensuring that facilities have appropriate human resource capacity to provide the services. In addition, they must ensure that the district hospital and DHE fulfil the additional requirements as outlined in Section 1.1.3.

##### TB focal nurse and district TB coordinator

The TB focal nurse should oversee the clinical aspects of TB and TB/HIV activities at facility level. The district TB coordinator, in liaison with the TB focal nurse, should oversee all TB and TB/HIV activities within the district. This must include oversight of community-based activities to support TB case finding, retention in care of TB patients and coordination of all TB and TB/HIV data collection. They should also support implementation of intensified case finding, TB preventive therapy and TB infection control policies within the facilities.

##### Doctors and clinical officers

A doctor or clinical officer based at district level should:

- Assist in the general running of HIV prevention, care and treatment services in collaboration with the focal person/nurse in charge
- Ensure that a thorough history and examination is performed and that appropriate investigations are ordered for clients seen
- Assess complicated cases referred to them by the nurses within the district HIV prevention, care and treatment centre and from referring clinics
- Support the HIV prevention, care and treatment nurse mentor in decisions to switch to second-line ART
- Review cases of second-line failure and refer as appropriate
- Support the management of DRTB cases
- Determine whether referral to a more specialised level of care is appropriate.

A doctor at district level should also be identified to be part of the district mentoring team. As well as participating in clinic visits, a doctor should be available by phone for nurses to contact with clinical queries. This role can be rotated among the doctors within the district.

At provincial and tertiary level, the doctors should be available to discuss complicated cases and accept referral as appropriate. If specialist consultants visit provincial or district sites, referring facilities should be aware of the scheduled dates.

#### Table 7: Health care worker scope of practice for the provision of HIV prevention, care and treatment services

| ACTIVITIES | DOCTOR AND CLINICAL OFFICER | RGN | PCN | NURSE AID | GENERAL HAND | PC | DATA CLERK | CLINIC -BASED MICROSCOPIST | EHT | CHW | EXPERT CLIENTS | CBO MEMBERS, SELECTED COMMUNITY MEMBERS |
|---|---|---|---|---|---|---|---|---|---|---|---|---|
| Registration and filling of appointment diaries | | Yes but should delegate | Yes but should delegate | X | X | X | X | | | | X | |
| Performing vital signs | | X | X | X | | | | | | | | |
| HIV testing and counselling | X | X | X | X* | | X | | X | X | X* | X* | X* |
| DBS for DNA PCR testing | X | X | X | X | | X | | | | | | |
| Management of complicated cases (e.g., CCM, second- line failure treatment failure etc) | X | | | | | | | | | | | |
| ART initiation and follow up for adults and children | X | X | X | | | | | | | | | |
| TB initiation of smear or Xpert positive cases for adults and children | X | X | X | | | | | | | | | |
| TB initiation for adults requiring CXR interpretation and children where no sputum is available | X | | | | | | | | | | | |
| Distribution of refills | | X | X | X | | | | | | X | X | X |
| Management of first -line treatment failure | X | X** | X** | | | | | | | | | |
| ART preparation and adherence counselling for adults, children and pregnant women including treatment failure | X | X | X | X | | X | | | | X | X | X |
| Defaulter tracing | | X | X | X | | X | | | X | X | X | X |
| Data entry (for area of service ) | | X | X | X | | X | X | | | X | X | X |
| Phlebotomy | X | X | X | | | | | | | | | |

* with adequate training

** with discussion with nurse mentor or doctor

#### Nurse in charge of a facility or HIV prevention, care and treatment unit

In addition to the clinical tasks involved in the provision of HIV prevention, care and treatment, the nurse in charge of the facility should ensure that:

- The minimum package of HIV prevention, care and treatment services (Table 6) is available within their facility and staff are allocated their daily duties to provide these services
- They have the necessary skills to manage and refer complicated cases
- Staff within her/his facility are adequately trained
- Pharmacy and laboratory commodities are ordered timeously and stored and utilised correctly
- Accurate and timely monthly data is submitted to the district
- Clinic team meetings are organised to discuss challenging cases, including treatment failure, and to give feedback of analysed data to support quality improvement activities
- Liaise with the community nurse, who will organise meetings with community leaders and CBOs to facilitate linkage between clinic and community-based activities
- Samples are taken, stored correctly and prepared for sample transport.

#### Clinic nurse

A clinic nurse should:

- Provide all clinical services required to implement the minimum package of care for children, adolescents and adults (including pregnant and breastfeeding women)
- Initiate and follow up children, adolescents and adults (including pregnant and breastfeeding women) on ART, including provision of counselling if required
- Initiate and follow up children, adolescents and adults (including pregnant and breastfeeding women) with a positive smear or Xpert MTB/Rif result on TB medication
- Follow up children, adolescents and adults (including pregnant and breastfeeding women) initiated on TB medication by the doctors (those diagnosed on CXR or clinically)
- Ensure that clear documentation of the consultation is made in the client-held notebook and the clinic-held patient care and treatment book
- Ensure that all baseline and follow-up laboratory tests are performed according to the clinical guidelines
- Ensure that medicines are prescribed and dispensed accurately
- Support the provision of refill services (Section 2.5.3-2.5.7)
- Ensure accurate and timely documentation of all activities in the designated registers  or EPMS according to the activity carried out (HTC, ART care)
- Assist the nurse in charge with the compilation of accurate and timely monthly reports.

#### Primary counsellor

The primary counsellor is responsible for providing:

- HTS for children, adolescents and adults (including pregnant and breastfeeding women), including preparation of DBS specimens for DNA PCR testing
- Basic HIV education, ART education and ART initiation counselling for children, adolescents, adults (including pregnant and breastfeeding women) (Job Aide, page 105)
- Follow-up adherence counselling for children, adolescents, adults (including pregnant and breastfeeding women)
- Enhanced adherence counselling for children, adolescents and adults (including pregnant and breastfeeding women) who are failing their treatment (Page 79-83)
- TB and DRTB adherence counselling.

**For all the above tasks, the counsellor must document their findings in the client-held notebook and the notes section of the patient care and treatment booklet.**

For HTS activities, the primary counsellor must also complete the HTS register and, if instructed by the nurse in charge, compile the monthly HTS report.

According to the setting, the primary counsellor could also be assigned tasks, such as registration of clients, updating of the appointment diary, initiation of the defaulter tracing process and acting as focal person for the formation and support of clubs and CARGs (Section 2.5.4). It remains the responsibility of the nurse in charge to clearly allocate duties and ensure that they are carried out.

#### Nurse aid

The nurse aid should assist the clinic nurses in the provision of the minimum package of HIV prevention, care and treatment services. Depending on the setting, some tasks could be assigned to the nurse aid; these include checking of vital signs, registration of clients, ensuring that the diary is accurately updated and ensuring that defaulter tracing is implemented. Nurse aids could be considered as an additional cadre to perform HTS, DBS or facilitate some ART refill strategies after receiving some prerequisite training. It remains the responsibility of the nurse in charge to clearly allocate duties and ensure that they are carried out.

#### Data clerk

The data clerk is part of the medical team at the facility. They are responsible for ensuring that data is accurately entered from the patient care and treatment booklet into the paper-based or electronic patient monitoring system on a daily basis. They should also support analysis of facility-based data, provide feedback to healthcare workers and provide the automatic appointment list on a weekly basis.

Depending on the setting, the data clerk could be assigned certain tasks, such as registration of clients, updating of the appointment diary, and initiation of the defaulter tracing process. It remains the responsibility of the nurse in charge to clearly allocate duties and ensure that they are carried out.

##### Environmental health technician (EHT)

EHTs perform a wide range of activities. Regarding HIV prevention, care and treatment activities, EHTs should:

- Support defaulter tracing under the guidance of the clinic nurse in charge
- Where appropriate, support weekly sample transport under the guidance of a district sample transport strategy.

##### Health promotion officer

The health promotion officer is responsible for activities to support community mobilisation, advocacy and health promotion messages related to the activities described within the minimum package of HIV prevention, care and treatment.

##### Pharmacist and pharmacy technician

The district-level and provincial-level pharmacist or pharmacy technician must ensure that their facility and the clinics they are supporting have an uninterrupted supply of quality medicines (according to the Zimbabwean pharmacy standard operating procedures) and ensure that emergency orders are supplied (Section 3.1). When dispensing OI and ARV medications, they should also advise on treatment literacy. Any serious adverse events should be reported by the clinicians and recorded appropriately.

Pharmacy staff should support, in collaboration with the nurse in charge, the provision of differentiated ART delivery (Section 2.5.4), which for certain models, may include pre- packing of drugs according to the SOPs outlined on pages 56-66.

In addition to ensuring that activities outlined in Section 3.1 are carried out, the pharmacist or pharmacy technician should perform regular (quarterly) supportive supervision visits to all clinics. These visits should be combined with existing visits for mentorship and supportive supervision by the DHE team to avoid the need for additional transport arrangements.

##### Laboratory scientist and technician

The district and provincial level laboratory scientist or technician must ensure that their facility and the clinics they are supporting have access to the minimum package of diagnostic and monitoring tests required for the provision of HIV prevention, care and treatment services as outlined in Section 1.1.2 They must ensure timely ordering of supplies and have a system in place to ensure access to investigations if a machine breakdown occurs. Adequate internal and external quality control must be ensured for all tests, including rapid HIV testing and other POC tests being performed at clinic level. In addition to ensuring that activities outlined in Section 3.2 are carried out, the laboratory scientist or technician should perform regular (quarterly) supportive supervision visits to all clinics. These visits should be combined with existing visits for mentorship and supportive supervision by the DHE team to avoid the need for additional transport arrangements.

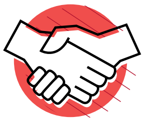

##### Community health workers and expert clients

Community health workers and other community-based cadres should:

- Conduct community mobilisation to increase uptake of HIV testing and counselling, care and treatment
- Act as a link between outreach and community-based activities and the facility to support linkage and retention to treatment services
- Provide community-based treatment literacy to support adherence and retention, including supporting clients with high viral load
- Link with the facility to provide defaulter tracing for clients utilising the AIDS and TB referral form to support effective communication
- Facilitate the formation and running of community-based ART delivery models where implemented (see Section 2.5.4)
- Ensure that monitoring and evaluation of community-based activities is linked to the respective facility.

#### 1.2.3 Capacity building

To provide the minimum package of HIV prevention, care and treatment services, the available human resources must be adequately trained. District health executives should ensure that a **regular assessment of training needs** is carried out and that there is a functioning district clinical mentorship team. Organisation of **clinical attachments** to the district or centre of excellence should be regularly scheduled to support nurses needing clinical skills enhancement in HIV prevention, care and treatment. Rotation of staff must be planned, considering their skills and investment in the staff through trainings, clinical attachments and mentorship, with the aim being to ensure that the appropriately trained and skilled staff members are in the right department at the right time.

In addition to classroom-based training, a national clinical mentorship programme in Zimbabwe was established in 2007. The goal of the programme is to scale up high-quality comprehensive HIV prevention, care and treatment services supporting decentralisation, building capacity in health care workers and motivating them with support. In addition to workshop-based trainings provided through the MoHCC, continuous capacity building and professional development is encouraged through the district mentorship programme. The programme utilises blended learning approaches, e-consultations and teleconsultations within the MoHCC structures. The programme is currently being scaled up with the goal of covering 80% of districts by the end of 2017 and every district by the end of 2018.

Mentorship entails **site visits to provide face-to-face support to the mentees** and also **telephone and other electronic support** for discussion of difficult cases. The province and district management teams must plan for clinical mentorship based on the needs identified through data analysis, supportive supervision and other programme review platforms. Based on this analysis, they should prioritise sites, cadres, focus areas and frequency for mentorship. Initially, sites should be mentored more frequently, and then less frequently as mentoring objectives are met according to established monitoring and evaluation tools. The mentorship team should support all human resources within the facility who are involved with providing HIV prevention care and treatment services. In particular, the primary counsellor should receive ongoing mentorship in order to provide counselling activities utilising the correct tools as developed in this manual and the Job Aide.

The content of the mentorship should be guided by the content of the clinical *Guidelines for Antiretroviral Therapy for the Prevention and Treatment of HIV in Zimbabwe*, the *Operational and Service Delivery Manual* and the accompanying Job Aide. In addition to clinical skills, the scope of the mentorship programme should include knowledge of and implementation of operational strategies across the cascade of care. These include implementation of appointment and defaulter tracing systems, strategies for provision of differentiated testing, ART initiation and ART delivery for both stable and unstable clients. The district mentorship team should be engaged in the decision-making process to determine which strategies are chosen in a district and lead the introduction of differentiated service delivery as outlined in Chapter 2. Ongoing monitoring and evaluation of the mentorship programme at district level should be carried out to determine impact and effectiveness.

Each district should select appropriate mentors. Core competencies of mentors are:

#### A. Expertise in HIV, STI and TB care

a) In-depth knowledge of current national guidelines on HIV, STI and TB prevention, care and treatment services
b) Familiarity with other related guidelines
c) Continuous updating of knowledge, especially on comprehensive HIV, STI and TB care
d) High-quality clinical assessment of patients (history and physical examination) and efficient documentation in the standard documents and M&E tools.

#### B. Knowledge of the Zimbabwe health care delivery system

a) Knowledge of and experience in the Zimbabwe health care delivery system is critical as the mentor is expected to assist mentees in addressing facility challenges and system issues.

#### C. Interpersonal process skills

a) Excellent communication skills
b) Giving non-judgmental and empathetic feedback
c) Self-awareness (awareness of one's own qualities and limitations).

#### D. Mentoring skills

a) Communication with the mentee
  - Demonstrating technical skills and knowledge in interaction with the mentee
  - Ensuring the ongoing development of the mentee
  - Disseminating clinical practice and information updates.

b) Knowledge of ICT to facilitate off-site mentoring and blended learning initiatives.

Mentors must receive mentorship training conducted in line with the standard national clinical mentoring curriculum and training manuals. The training focuses on communication skills, adult educational principles, the national guidelines, protocols and tools relevant in practicing as an HIV, STIs and TB clinical mentor, including the *Operational and Service Delivery Manual*, the health delivery system, the mentoring programme itself, and the details of the mentoring approaches. The mentor training involves an assessment of mentoring competencies, and practicing mentors undergo an appraisal and accreditation process yearly to ensure competency, assure quality and encourage continuous medical education and professional development.

Selection of appropriate and competent mentors is critical to the success of the mentorship programme. Mentors are selected to be part of a **multi-disciplinary team** that collectively has the expertise to address programme needs and gaps identified in the provision of HIV, STIs and TB services and related health delivery system issues. Mentoring multi-disciplinary teams are made up of, but are not limited to, a medical doctor, nurse, pharmacist/pharmacy technician, primary counsellor, laboratory scientist/technician and monitoring and evaluation officer. The respective MoHCC leadership at the different levels (DHE, PHE, City Health Directorate and hospital executive) is responsible for selecting appropriate cadres to be trained and practice as mentors.

Mentorship must be an ongoing process. The development of a trusting relationship between the district mentorship team and the health care workers providing services in the decentralised sites should strengthen the quality of service provision and motivate staff in their daily tasks.

#### 1.2.4 Key messages and reference materials

- Providing the HIV prevention, care and treatment minimum package requires TEAM WORK.
- With the growing cohort of clients, “ who does what” will need ongoing evaluation.
- All doctors, clinical officers, registered nurses and primary care nurses can initiate and follow up adults (including pregnant and breastfeeding women), adolescents and children on ART.
- Community health workers, expert clients and other community workers should be actively involved in prevention, treatment literacy activities and defaulter tracing.
- Clinic and district managers should be clear about who is responsible for each task within their facility.
- All health workers should rotate through departments or activities so they become polyvalent, but rotation of staff must be structured to ensure that adequately trained staff members are available at any point to provide the minimum package of HIV prevention, care and treatment.
- Ongoing capacity building is essential. All DHEs should undertake regular reviews of training needs.
- All DHEs, in collaboration with the national mentorship programme, should plan for implementation and ownership of a multi-disciplinary district mentorship programme for on-site and off-site mentoring to facility staff providing HIV prevention care and treatment services, including counselling and monitoring and evaluation.
- Goals of mentorship should be agreed between the mentor and mentee at the start, but must include both clinical and service delivery components.
- Mentorship should cover clinical and operational and service delivery aspects of care.
- Mentorship should include the whole clinic team engaged in HIV care, including the primary counsellors.
- Mentorship is an ongoing process, which should also be linked with the goals of quality improvement (Section 3.4).

##### Reference materials

Guidelines for Clinical Mentoring of Comprehensive HIV Care and Treatment Services in Zimbabwe.

### 1.3 Decentralisation of HIV prevention, care and treatment services

#### 1.3.1 Background

Access to ART for many clients still remains a challenge and, of those who access it, many find it difficult to remain on treatment due to time- and cost-related constraints. Decentralisation of HIV prevention, care and treatment services, where services are taken closer to the client's place of residence, is a strategy that may both reduce congestion at centralised sites and reduce the burden on clients. Evidence suggests that decentralisation of services can significantly reduce loss-to-follow-up rates.

Decentralisation is a key principle for the organisation and management of the National HIV Care and Treatment Strategic Plan in Zimbabwe. According to two national ART outcomes studies conducted in Zimbabwe in 2010 and 2016, those initiating ART at primary health care facilities had better retention rates compared with those initiated at higher levels of care; this was attributed mainly to decentralisation and nurse-led provision of ART initiation services.

Zimbabwe's ART programme has expanded rapidly from the initial **seven sites in 2004 to 1545 sites (94% of all health facilities) providing ART at the end of 2015**. Of the 1545 sites, 1475 are ART-initiating sites with the remainder providing ART follow-up services. This is a significant milestone for the national ART programme, whose goal was to ensure that all facilities in the country provide the minimum package of HIV prevention, care and treatment services by the end of 2015. Capacity assessments, ongoing trainings, mentorship and supervision support have since been strengthened and decentralised to the district level.

Decentralising HIV care presents an opportunity to strengthen community engagement and linkages across the continuum of the HIV prevention, care and treatment cascade.

#### 1.3.2 Planning for decentralisation

Adequate planning for decentralisation of HIV care, treatment and support services is essential. The planning should be spearheaded by the District Health Executive team and should include the following core staff:

- District medical officer
- District nursing officer
- District HIV focal person
- District TB coordinator
- District pharmacy manager
- District health services administrator
- District laboratory in charge
- District health information officer
- District environmental health officer

Other key stakeholders, such as representatives from the DAAC, community leaders, CBOs, faith-based organisations and people living with HIV/AIDS, should be consulted whenever possible throughout the process.

The planning team should ensure that it has the capacity to assess the ART readiness of a facility by using the Health Facility Comprehensive HIV/AIDS Capacity Assessment Tool.

Key areas for assessment to ensure a functional decentralised site include:

- **Human resources – nurses:** How many nurses have received the HIV integrated training and are able to provide the minimum package of HIV prevention care and treatment services?
- **Human resources – counselling:** How many trained nurses and/or primary counsellors are able to provide HIV testing services, ART preparation, follow up and enhanced adherence counselling for children, adolescents and adults (including pregnant and breastfeeding women)?
- **Human resources – doctor:** Is there a medical doctor available for regular outreach to the decentralised sites and/or available for remote mentorship support?
- **Pharmacy:** Is there capacity for supply to the site; is there capacity for medicine storage; are staff trained to order HIV-related medicines (Section 3.1)?
- **Laboratory:** Is there a regular and reliable sample transport system; is there capacity at district level to process samples and ensure quality assurance from all sites (Section 3.2)?
- **Monitoring and evaluation:** Are all the tools required for HIV prevention, care and treatment services available and are staff adequately trained to use them (Section 3.3)?
- **Community involvement and linkages:** To what extent is the community involved and is there a plan for community linkages to support access and retention as outlined in the minimum package?

- **Physical space:** Is there adequate space to ensure that clients are consulted, counselled and examined confidentially and privately; is there secure and adequate space for storage of medicines?
- **Mentorship:** Is there a district mentorship team formed and adequately resourced to support decentralised sites (Section 1.2.3)?
- **Supportive supervision:** Is there a plan for a district-wide supportive supervision system?

##### 1.3.3 Key messages and reference materials

- The minimum package of HIV prevention, care and treatment services (Section 1.1.2) should be available at all health facilities in Zimbabwe.
- Successful decentralisation requires nurses at primary care level to recognise serious red-flag conditions, including treatment failure, that should be referred when appropriate.
- Successful decentralisation requires a "safety net" referral site with capacity to investigate and manage complicated cases that cannot be managed at primary care level.
- Accreditation for decentralisation of ART services involves assessment of human resource capacity, pharmacy supply chain management system, laboratory services systems, monitoring and evaluation systems, and infrastructure.
- Successful decentralisation requires ongoing mentorship AND ongoing supportive supervision. This is the primary responsibility of the District Health Executive (DHE) with overall oversight provided by the Provincial Health Executive (PHE).

Decentralisation requires a strong link between the facility and community in order to mobilise the community to access HIV prevention, care and treatment services and to support ongoing retention and adherence.

Manual for Primary Health Facility Comprehensive HIV and AIDS Capacity Assessment.

### 1.4 Integration of services

#### 1.4.1 Background

The goal of integration (TB/HIV, SRH/MNCH/PMTCT or NCD/HIV) is to provide a one-stop service for the client:

- Under the same roof
- On the same day
- By the same health care professional.

Integrating services is aimed at reducing missed opportunities for initiation of ART, enhancing long-term adherence support and optimising client retention in care. TB, as the most common co-infection, has raised the challenge of one client simultaneously needing to be treated for two infectious diseases usually managed by two different programmes. With the massive scale up of PMTCT services, the need for integrated MNCH and HIV services has also raised itself as a challenge to existing health care systems. Finally, as people live longer with HIV, the challenge of managing other co-morbidities, such as hypertension, diabetes and mental health conditions, will also need to be incorporated into the service provision for clients on ART.

The challenges for integration of services vary according to the level of the health care system. At primary care clinics, there are often only two or three nurses providing services and, therefore, if the minimum package of HIV prevention, care and treatment services is being offered on all days, integration can be more easily achieved. In larger facilities where services are spread across departments, integration of services requires communication between departments, adequately trained staff and consolidation of monitoring and evaluation tools.

#### 1.4.2 TB/HIV integration

The aim of TB/HIV integration is to:

1. Increase HTS coverage among TB clients as an entry point to HIV care
2. Screen and diagnose active TB disease (including smear negative TB) in HIV-infected persons
3. Reduce the delay in initiating ART in co-infected clients.

The goal should be that all nurses are able to commence both TB and ART medicines and provide a one-stop service for the client. Table 8 outlines the 12 components of TB/HIV collaboration.

In large-volume sites where the TB and OI/ART clinics are separate, the goal is:

- For clients whose first diagnosis is TB and who enter the system through the TB clinic, initiate them on ART within the TB clinic (where possible). After successful completion of TB treatment, they are transferred to the OI/ART clinic.
- For clients on ART who develop TB, they should receive their TB treatment within the OI/ART clinic (where possible) but ensure that they are registered accordingly as per requirements of the TB programme.

For both scenarios, the client should be able to receive their ART and TB treatment on the same day, in the same consultation room from the same clinician.

##### Intensified case finding (ICF)

All HIV-positive clients should be screened for TB at every clinical visit (and all TB clients should be tested for HIV). Another opportunity for TB screening is during HTS. ICF serves two purposes: it identifies presumptive TB cases that need diagnostic work up and possible treatment; and it identifies those HIV-positive clients with no TB symptoms who may benefit from TB preventive therapy. TB screening algorithms can be found on page 95 and 96 of the Job Aide.

##### TB preventive therapy for people living with HIV

TB preventive therapy has been introduced in a phased approach, with the number of sites providing isoniazid (INH) as a preventive intervention increasing from an initial 10 pilot sites in 2013 to 354 high-volume sites by mid-2016. Under the programme, if TB screening is negative and clients meet the eligibility criteria for IPT, a six-month daily course of INH is given. Treatment should then be repeated every three years according to the criteria in the algorithm. For full guidance on TB preventive therapy, see document entitled *Implementation of Intensified TB Case Finding and Isoniazid Preventive Therapy in Zimbabwe: Step by Step Guide* (Collaborative TB/HIV Activities, April 2015).

##### TB infection control

The most infectious patients are those not yet diagnosed, i.e., those coughing in the OPD. Infection control measures can be divided into administrative, environmental and personal. Making sure that simple infection control measures are in place can make a big impact on reducing the transmission of TB for health care workers and patients. Each hospital should have an infection control committee and each department or clinic should have an infection control focal person.

#### Table 8: 12 components of TB HIV collaboration

| | |
|---|---|
| **A. STRENGTHENING THE MECHANISMS FOR DELIVERING INTEGRATED TB AND HIV SERVICES** | |
| A1 | Strengthening coordinating bodies for collaborative TB/HIV activities functional at all levels |
| A2 | Determining HIV prevalence among TB patients and TB prevalence among people living with HIV |
| A3 | Carrying out joint TB/HIV planning to integrate the delivery of TB and HIV services |
| A4 | Monitoring and evaluating collaborative TB/HIV activities |
| **B. REDUCING THE BURDEN OF TB IN PEOPLE LIVING WITH HIV AND INITIATING EARLY ANTIRETROVIRAL THERAPY (THE THREE I'S FOR TB/HIV)** | |
| B1 | Intensifying TB case finding and ensure high-quality anti-tuberculosis treatment |
| B2 | Initiating TB prevention with isoniazid preventive therapy and early antiretroviral therapy |
| B3 | Ensuring control of TB Infection in health care facilities and congregate settings |
| **C. REDUCING THE BURDEN OF HIV IN PATIENTS WITH PRESUMPTIVE AND DIAGNOSED TB** | |
| C1 | Providing HIV testing and counselling to patients with presumptive and diagnosed TB |
| C2 | Providing HIV prevention interventions for patients with presumptive and diagnosed TB |
| C3 | Providing cotrimoxazole preventive therapy for TB patients living with HIV |
| C4 | Ensuring HIV prevention interventions, treatment and care for TB patients living with HIV |
| C5 | Providing antiretroviral therapy for TB patients living with HIV |

**Administrative:** Cough triage should be performed. Any coughing client should be identified and, ideally, given a surgical mask where resources permit. Cough hygiene should be promoted in the waiting area. Coughing clients or any known smear positive client, until converted, should be fast tracked for assessment.

**Environmental:** One of the most effective ways this can be done without costly interventions is to ensure that there is good cross ventilation in a room and existing windows are opened. Position the desk and chairs so that any airflow is from the health care worker toward the client.

**Personal:** Ideally, N95 masks should always be worn when managing a coughing client. However, they are often not available. Health care workers are, however, mandated to wear them if dealing with an MDR SUSPECT (not only a diagnosed patient).

##### 1.4.3 MNCH/HIV integration

The aim of MNCH/SRH/HIV integration is to:

1. Increase HTS among pregnant and breastfeeding women and their partners
2. Reduce the delay in initiating ART for those who test positive
3. Ensure re-testing of those who initially test negative
4. Reduce loss to follow up of the mother and her exposed baby by providing a family-centred approach in the under-five clinic.

All women attending ANC, labour and delivery and PNC services must have access to HTS and PMTCT services as a one-stop service. This means that all nurse midwives must be trained in the provision of the HIV prevention, care and treatment minimum package. The mother, child and, ideally, father should be seen within MNCH services until the exposed or infected child is five years old.

Women receiving HIV care should also be able to receive family planning services as a one-stop service as part of the effort to reduce unintended pregnancies in HIV-positive women of reproductive age.

For full details of all the SRH/HIV linkages, refer to the service guideline on SRHR and HIV linkages in the reference materials.

#### 1.4.4 NCD/HIV integration

In line with the WHO *2016 consolidated guidelines on the use of antiretroviral drugs for treating and preventing HIV infection*.

**To identify cardiovascular risk factors, all HIV-positive patients on ART should have their blood pressure and cardiovascular risk assessed during an annual review.**

HIV itself and a number of the antiretroviral medications are risk factors for cardiovascular disease. In addition, as the cohort ages, clients will increasingly present with other chronic co-morbidities.

For those clients already diagnosed with a chronic co-morbidity, such as hypertension, diabetes, asthma or epilepsy, follow up should, where possible, be integrated with ART clinical follow up (same day, under the same roof, by the same nurse). Once stable, medication refills for their co-morbidities should be offered using the same refill system as their ART, if drug supply allows (see Section 2.5.4).

**All clients, during their annual review, should be screened for anxiety and depression using two initial screening questions:**

- During the past month, have you felt like you were losing interest or pleasure in doing things?
- During the past month, have you felt down, depressed or helpless?

If there is a positive answer to either of these questions or if the client presents with red-flag issues (virological failure, missed appointments, challenging psychosocial issues), a formal mental health assessment using a standardised tool, such as the Shona Symptom Questionnaire, should be conducted (See Appendix 3). Clients should then be managed with counselling interventions or, where appropriate, referred for a formal mental health assessment.

Clients with severe mental illness who are stable and on long-term medication should be able to receive their chronic repeat medication via a similar refill system as their ART while having specialist clinical review as appropriate.

#### 1.4.5 Key messages and reference materials

##### Service providers should aim to provide a one-stop service for clients

- Under the same roof
- By the same health care provider
- On the same day.

##### TB/HIV integration

- At primary care clinics, both diseases should be managed as a one-stop service
- Where there are separate OI and TB clinics, a collaborative approach should be adopted
- All HIV-positive clients should be screened for TB
- All TB clients should be offered HTS.

##### SRH/MNCH/HIV integration

- HIV testing services and prevention (provision of condoms and promotion of VMMC) should be available at all SRH entry points (family planning, ANC, labour and delivery, PNC).
- Access to PMTCT should be available as a one-stop service in ANC, labour and PNC.
- Stand-alone family planning units should be able to test for HIV and refer to facilities providing ART.
- All women on ART should be able to receive their ART and family planning as a one-stop service.
- Provision of the minimum package of HIV prevention care and treatment (Section 1.1) as a family-centred approach should be provided within MNCH until the child (exposed or infected) is five years old. After that, the mother, father and any HIV-positive child is referred back to the OI service.

#### NCD/HIV integration

- All HIV-positive clients should have an annual blood pressure check and assessment of cardiovascular risk during their HIV clinical review.
- All HIV-positive clients should have a brief assessment of depression and anxiety. If positive or if there are other risk factors, such as high viral load or missed appointments, a formal assessment using the Shona symptom screening tool should be carried out.
- Stable clients on ART with other chronic co-morbidities should have their refill medication via the same system where possible.
- Where possible, clinical review of stable clients on ART with other chronic co-morbidities should have their clinical review integrated on the same day, under the same roof, by the same clinician.

#### Reference materials

Service Guideline on SRHR and HIV Linkages, August 2013.

### CHAPTER 2

#### Differentiated Service delivery

### 2.1 Introduction to differentiated service delivery

With the introduction of "Treat All" and the intention to meet the 90-90-90 targets, innovative strategies are needed to identify those who currently do not know their status, link them to care and retain them on ART within a health system that is already over-burdened. Continuing to provide services in the same way for all clients regardless of their differing needs is not only inefficient for the health system, but also places an unnecessary burden on the client.

Differentiated service delivery has been defined as a **client-centred approach** that simplifies and adapts HIV services across the cascade to reflect the preferences and expectations of various groups of people living with HIV (PLHIV) while **reducing unnecessary burdens** on the health system. By providing differentiated care, the health system can refocus resources to those most in need.

The principles of differentiated care are aimed at supporting both the achievement of the 90-90-90 targets and the introduction of "Treat All" while improving the quality of services for clients and responding to the increasing workload faced by health care workers. Describing models of care as differentiated has been driven by the need to provide adapted services according to three elements: the clinical characteristics; the sub-population (e.g., pregnant, adolescents, key population); and context (e.g., urban or rural, conflict or stable setting) (Figure 2).

#### Figure 2: The three elements of differentiated service delivery

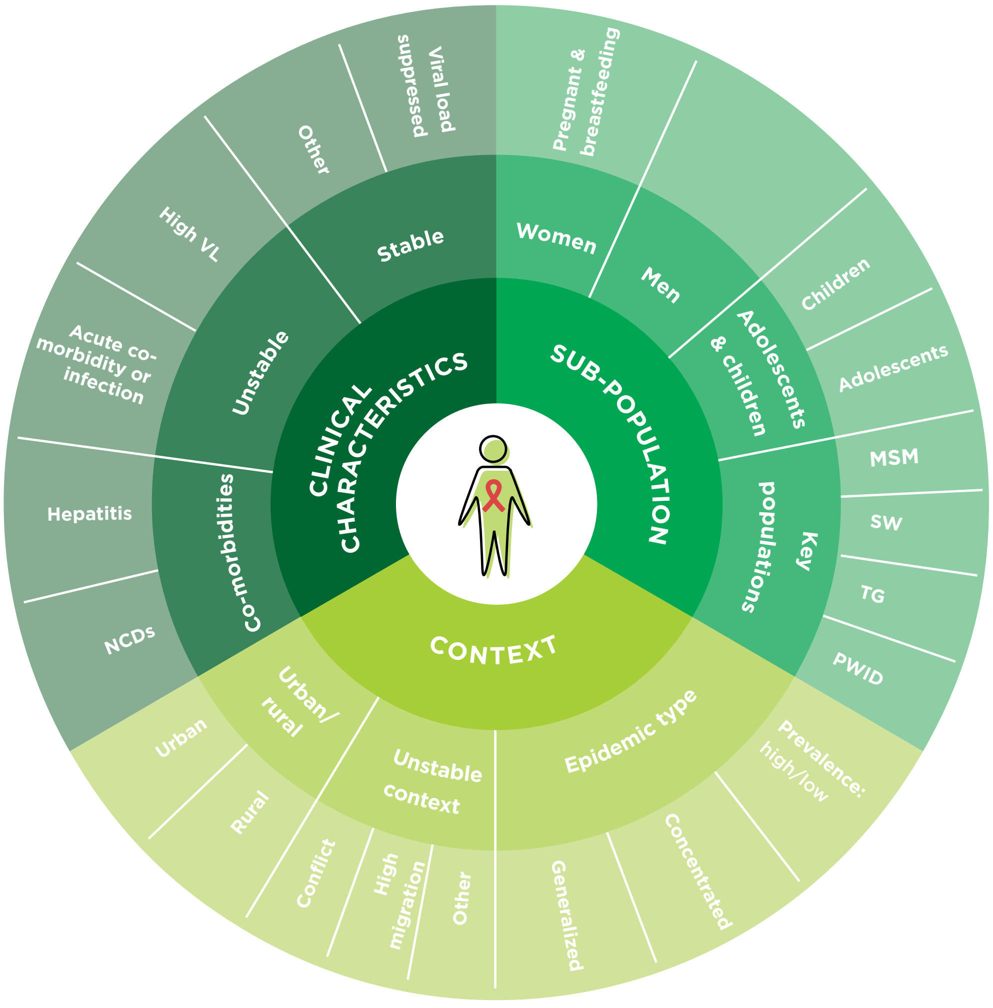

Once the elements have been selected, a model of testing, prevention, initiation or ART delivery can be built using the building blocks of when, where, who and what is provided as part of a specific service.

At each step of the HIV prevention, care and treatment cascade, differentiated models of service delivery should be designed and implemented as a direct response to specific challenges or barriers identified for clients and health care workers. Each of the following sections of Chapter 2 are aimed at using these elements and building blocks to look at how testing, prevention, initiation and ART delivery may be differentiated to achieve the 90-90-90 goals in the era of "Treat All".

#### Figure 3: The building blocks of differentiated service delivery: ART delivery example

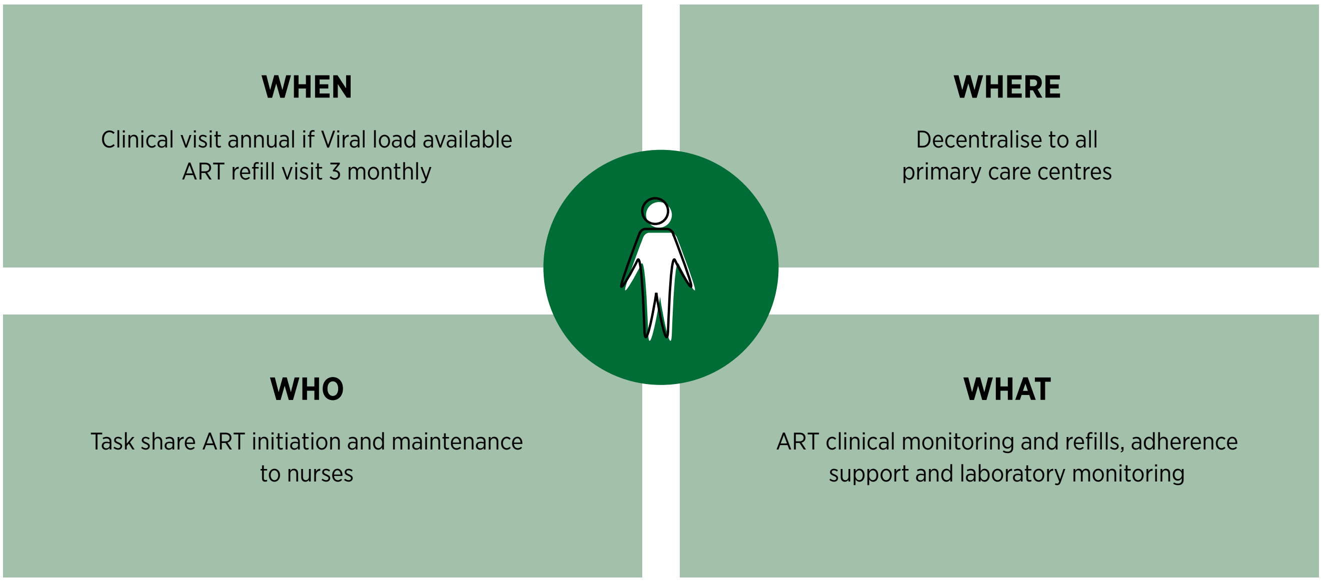

### 2.2 Differentiated HIV testing services (HTS)

#### 2.2.1 Background

Accessible and acceptable access to HIV testing services is the first critical step of the HIV treatment cascade. Globally, testing remains the biggest challenge to meeting the 90-90-90 targets, with 13.4 million people living with HIV worldwide being unaware of their status. In Zimbabwe, 1581 (96%) of all facilities offer HTS, with 2.2 million people having been tested in 2015. However, to reach those people living with HIV who do not yet know their status, who are clinically well and have high CD4 cell counts, innovative approaches to HIV testing services are needed. Using the principles of differentiated service delivery (adapting services according to the elements and building blocks), testing services may be adapted to target both high-risk populations and those that are currently hard to reach.

Different testing approaches will have different positivity yields.

**High-yield testing strategies:** facility-based PITC, facility- and community-based index client testing, targeted testing of high-risk populations (female sex workers, MSM, people who inject drugs).

**Low-yield testing strategies:** door-to-door community testing, general community campaigns.

Although high-yield strategies are likely to be more cost effective, the benefit of linking those testing HIV-negative to prevention services, such as VMMC, and condom use should not be forgotten.

#### 2.2.2 When are HTS provided?

HTS should be available during the standard opening hours of the facility. For sites with inpatient or maternity services, it should be ensured that a staff member trained to test is available overnight and at weekends. The timing of community-based testing services should be adapted to the specific sub-population being targeted.

#### 2.2.3 Where are HTS offered?

##### Facility-based HTS: provider-initiated testing and counselling (PITC)

PITC services with an opt-out strategy should be provided to all adults, adolescents and children attending all health facilities as the recommended standard of care. In addition to rapid HIV testing, this must include access to DBS for DNA PCR testing for infants less than 18 months. HTS should be provided: within antenatal care (ANC); at tuberculosis (TB) clinics; at sexually transmitted infection (STI) and outpatient clinics; in medical, surgical and paediatric wards; within maternal, newborn and child health (MNCH) services, including EPI; and within reproductive health, family planning, adolescent SRH, nutrition, mental health and male circumcision services.

Pre-test information on HIV should be given as a group where possible, after which the individual can decide whether to proceed or opt out. In OPD, the health care worker responsible for the health education talk should give the group pre-test information first thing in the morning, and try to repeat it mid-morning for latecomers to the clinic. Those who opt in can proceed immediately to having the HIV test performed by the HTS provider, ideally prior to their clinical consultation. Clients who opt out are the ones who require further individual pre-test counselling to understand their barriers to HIV testing. Once tested, the main emphasis is on quality individual post-test counselling.

##### Facility- and community-based index client testing

**Index client HTS** is a strategy whereby partners and family members of an identified HIV-positive case are offered HTS. Index case testing may be performed either at the facility or in the community. The following steps should be followed when implementing index client testing:

- Every client testing positive during facility-based or community-based testing should be asked to consent to testing of partners and family members. This consent should be documented.
- All partners and family members should be listed on page 5 of the patient care and treatment booklet. This page should be regularly checked to ensure that the status of all family members is known.
- The client should first be asked to bring their partner and family members to the facility for testing. With consent, the client may be linked with a community health worker or expert client who may support and encourage family members to attend. This referral should be made using the AIDS and TB referral form (Appendix 1).
- Previous partners should also be contacted either directly by the client or, with their consent, anonymously by the health care worker.
- If the client indicates that it will not be feasible to bring family members to the facility or if, after one month in care, the status of family members is not known, consent for community-based testing should be sought.

Community-based index client testing may be performed through the following strategies:

- Facility-based health care worker, e.g., primary counsellor or nurse performs targeted index client community-based testing once per month at an agreed time for the clients.
- The facility-based primary counsellor or nurse links with a community-based cadre who is trained to test. This may be through supervised use of self-tests or through a test for triage strategy where the community health worker (CHW) performs one rapid test. In both cases, if the single test is positive, the client must be referred to the facility for the full algorithm to be performed.
- The facility-based health worker teaches the client how to perform a self-test and provides self-tests to the client for unsupervised self-testing to be performed at home.

##### Community-based testing strategies

Community-based testing strategies have been shown to increase access to HTC by reducing costs of transport for the client, reducing stigma and increasing convenience of testing. It has been demonstrated to increase the number of men and children tested, people using couple HTS and the number of first-time testers, as well as identifying more clients with higher CD4s.

Community-based testing may be targeted towards specific high-risk populations (see Section 2.2.9, special considerations for sub-populations) by locating testing in specific locations, e.g., hotspot testing for female sex workers or generalised testing, such as door-to-door or general campaign events. The former strategy will be high yield and the latter has been shown to be low yield.

**Mobile/outreach HTC campaigns** have been utilised as strategies for some time in Zimbabwe. Their frequency and how they are targeted to specific populations, however, should be coordinated by the District Health Executive (DHE) teams. Each facility should plan **at least one outreach HTS activity per quarter**. Analysis of whether there are areas to target specific key populations should be made, along with analysis of facility HIV testing data to guide where best to target these activities. For example, if the proportion of men testing at the facility is low, an evening (moonlighting) outreach activity or one linked to a sporting event or a particular workplace may be more appropriate. Outreach activities aimed at educational facilities are also important to normalise HIV testing within the institution and encourage teachers and parents to test along with their children.

**HTS during EPI and clinic outreach activities.** Many sites have existing outreach activities (EPI and ANC) for hard-to-reach areas. HTS for adults, children and infants could be integrated into these activities. For this to be effective without compromising the existing activities, additional staff may have to join the team. DHE teams should coordinate these activities and consider the addition of a primary counsellor (PC) or nurse to support testing activities. Measures taken to ensure confidentiality (provision of a small tent) should be taken.

#### 2.2.4 Who performs HTS ?

##### Health care workers

All cadres of health care worker, as illustrated in Table 7, should be trained to perform HTS. To expand community-based testing, community health workers and lay cadres should be trained to perform HTS. This may be through performing HIV rapid tests or through the use of self-testing kits. The complete HIV testing algorithm may be performed or a test for triage approach may be taken. Test for triage is a strategy where a single HIV test is performed and, if positive, the client is referred to the facility for formal HIV testing to be performed. At all times, these cadres should be under the supervision of the facility nurse and quality assurance adhered to.

**To successfully scale up opt-out PITC in both outpatient and inpatient settings, PITC should be routinely initiated by the primary counsellor or health worker at the entry point and not be dependent on clinician referral.**

##### Clients: HIV self-testing

HIV self-testing (HIVST) is a process whereby an individual, who wants to know their HIV status, collects their specimen, performs the test and interprets the test result in private or in the presence of someone they trust. The oral self-test is a triaging test. It does not provide a final HIV diagnosis. Anyone who tests HIV positive using a triaging test must undergo another different test to confirm the diagnosis prior to being treated for HIV. HIVST provides confidentiality and empowers users to be responsible for their own HIV status. HIVST is being piloted to gather evidence on which may be the most cost-effective models to distribute self-testing kits; models may change based on evidence gathered. The ministry is in the process of mobilising funds to buy the kits for scale up of HIVST.

In Zimbabwe, test kits are currently distributed by a health care worker in a facility or by community health workers who are supervised by a health care worker. When the HIVST kit is distributed, basic pre-test information is given, permission to leave the test with the client or in their home is gained, and there is explanation of how to use the test kit. The HIVST kit also includes a self-referral form, which the client should present to the facility. Monitoring and evaluation tools are being piloted for self-testing.

In the future, HIV self-testing may be of particular value in increasing access to:

- Community-based index client testing
- Testing and repeat testing in high-risk key populations
- Partner testing, especially within PMTCT settings.

##### Couple testing

Couple HTS has a number of advantages. Because the couple has chosen to be tested together, mutual disclosure is immediate and, in the case of a sero-discordant couple, treatment can be offered immediately to the HIV-positive partner and PrEP offered to the HIV-negative partner. Although couple HTS has been emphasised as part of ANC and PMTCT, couple HTS should be encouraged as a general approach.

Community and male mobilisation strategies to encourage couples to test for HIV, is an essential component for community networks to consider. Outreach HTS may also target locations where couples are more likely to attend together. Self-testing may also be an approach to increase the uptake of couple testing.

#### 2.2.5 What services are provided?

##### Stand-alone versus integrated HTS

Integrating HTS into an offer for a general health screen may encourage more clients to access HTS, particularly in community-based testing strategies. Integrating screening for other diseases may also make such approaches more cost effective. Providing an integrated approach will require further training for any community-based cadre providing these services.

Screening for the following should be considered in an integrated approach:

- TB
- STIs (and possible immediate treatment)
- Diabetes (random blood sugar)
- Blood pressure
- Nutrition.

#### 2.2.6 Re-testing for HIV

Encouraging re-testing is increasingly important. Recommendations for re-testing for the general population, key populations and for HIV-negative pregnant and breastfeeding women can be found on page 88 of the Job Aide. A new recommendation in the 2016 *Clinical guidelines for antiretroviral therapy for the prevention and treatment of HIV in Zimbabwe* is for all clients to be re-tested prior to initiation of ART. Re-testing should ideally be conducted by a different service provider with a different specimen. However, if there is only one health worker at the facility, they can take another blood sample a few hours apart and re-test. Frequently asked questions regarding re-testing prior to ART initiation can be found in Appendix 2.

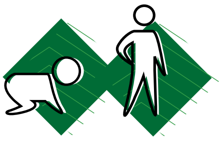

#### 2.2.7 Special considerations for testing children and adolescents

Testing of infants younger than 18 months should follow the early infant diagnostic algorithm outlined in the *Guidelines for Antiretroviral Therapy for the Prevention and Treatment of HIV in Zimbabwe*. Identifying HIV-positive children (0-9 years) and young (10-14 years) and older adolescents (15-19 years) is a challenge, especially in those adolescents below the age of consent (16 years) and where it may be difficult to gain consent from a parent or guardian.

All children under the age of five attending MNCH, outpatient or inpatient units should be routinely offered PITC. To offer routine opt-out HIV testing, staff trained to perform HTS and DBS preparation should be available across all departments. Regarding children older than five years, the HTS screening tool for children and adolescents (See Job Aide, page 18) may be used to prioritise those who should automatically be offered HIV testing. Questions to be asked:

- Has the child ever been admitted to hospital?
- Has the child had recurring skin problems?
- Has one or both of the child's natural parents died?
- Has the child experienced poor health in the past three months?
- Are there any symptoms or signs of an STI? (for adolescents)

IF YES TO ANY OF THE ABOVE, OFFER AN HIV TEST.

Figure 4 shows an example of how HTS could be differentiated for adolescents using the four building blocks of differentiated service delivery.

Wherever HTS is being offered, the health care worker should understand the following two important considerations regarding testing of children.

#### Figure 4: Differentiated HIV testing services for adolescents

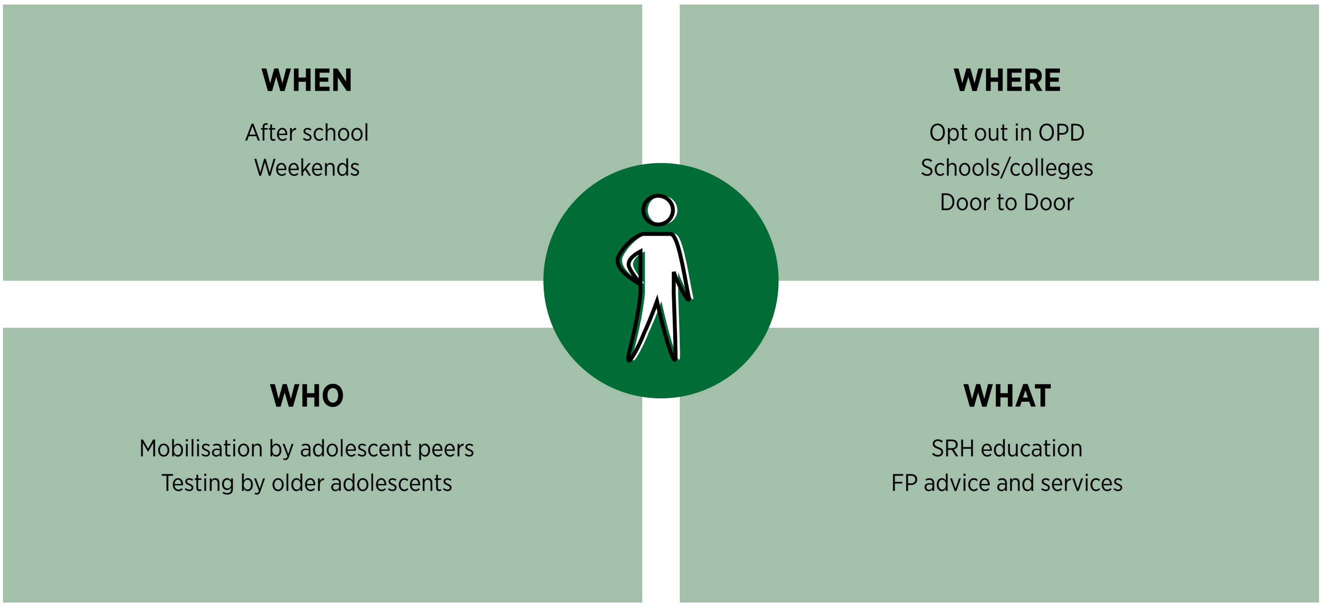

##### Age of informed consent for HTS:

- Any child who is aged 16 years or above, is married, pregnant or a parent or who requests HTS is considered able to give full informed consent.
- The consent of a parent or caregiver is required before performing an HIV test on a child who is below 16 years of age.
- A child below the age of 16 who is a mature minor may provide informed consent for HTS. A mature minor is a child or adolescent who can demonstrate that he or she is mature enough to make a decision on their own. A counsellor should consider the following factors in determining whether a child or adolescent should be treated as a mature minor:
  - The minor's ability to appreciate the seriousness of HTS and the test result and to give informed consent
  - The minor's physical, emotional and mental development
  - The degree of responsibility the minor has assumed for his or her own life, such as heading a household or living independently from a parent/caregiver.

If a parent or caregiver will not or cannot give consent for a child younger than 16 years, the health worker can exercise the "best interest of the child" principle and seek approval from the person in charge of the clinic or hospital to perform the HIV test.

##### "Best interests of the child" principle

A service provider should seek approval from the person in charge of the clinic or hospital in order to provide HTS without consent from a parent or caregiver when it is in the best interests of a child. This includes when:

- A child is ill and diagnosis will facilitate appropriate care and treatment
- A child is a survivor of sexual abuse
- A child is sexually active
- A child is concerned about mother-to-child transmission
- A child has been exposed to HIV through vertical or sexual transmission
- A child expresses concern that, given an HIV-positive result, he or she will be denied access to care and treatment by a parent/caregiver.

Please do not avoid performing HTS for children and adolescents. Further details of how to proceed with counselling and testing for a child can be found in the *Zimbabwe HTS Guidelines for Children and Adolescents (2014)*. If you do not feel comfortable testing a child or adolescent, please seek further support from your supervisor or local mentorship team.

#### 2.2.8 Special considerations for HTS for men

HTS services for men should be provided in a male-friendly environment. Some important considerations include the following:

- HTS services should be offered during times that are flexible for men so they can attend after working hours. Consider having one day a week with extended hours for men to access health care services.
- HTS should be offered as part of a more general male health screening package (e.g., hypertension and diabetes screening, alcohol and smoking advice) or linked to the provision of VMCC.
- Outreach testing to workplaces and community gatherings where men may attend.

#### 2.2.9 Special considerations for HTS for key and priority populations

Due to specific high-risk behaviours, key populations are at increased risk of being infected or affected by HIV. Key populations include sex workers, injecting drug users, transgender people and men who have sex with men (MSM). Priority populations are made up of people who are at an increased risk of HIV transmission because of their circumstances. They include mobile workers, adolescent girls and young women, truck drivers, artisanal miners, migrant populations, and people in prisons or other closed settings.

When designing a testing strategy for key populations, special considerations to the building blocks of differentiated testing should be made. The time that testing is offered should be adapted (e.g., moonlight testing for female sex workers); the location should be chosen to seek out a particular population, e.g., border points or trucking stops; and the role of peers should be explored both for mobilisation and performing of HIV testing services. Each group should also have specific additional services incorporated into an integrated screening approach. Linking HIV-negative key populations to prevention services, including PrEP, should be a priority. Figure 5 gives an example of differentiated testing services for sex workers.

#### 2.2.10 Documentation and M&E requirements for HTS

All clients undergoing HTS should be entered into the HTS register. Where testing is performed in the community, the testing activity must be linked back to the clinic responsible for that catchment area. Clients tested in the community should have an AIDS and TB referral form given to them to facilitate linkage to prevention or care and treatment services. A rapid test request form should be filled out for each client and the result slip completed. This result slip should be kept by the client to facilitate linkage to care either within the same facility or if being referred elsewhere.

##### Figure 5: Differentiated HIV testing services for female sex workers

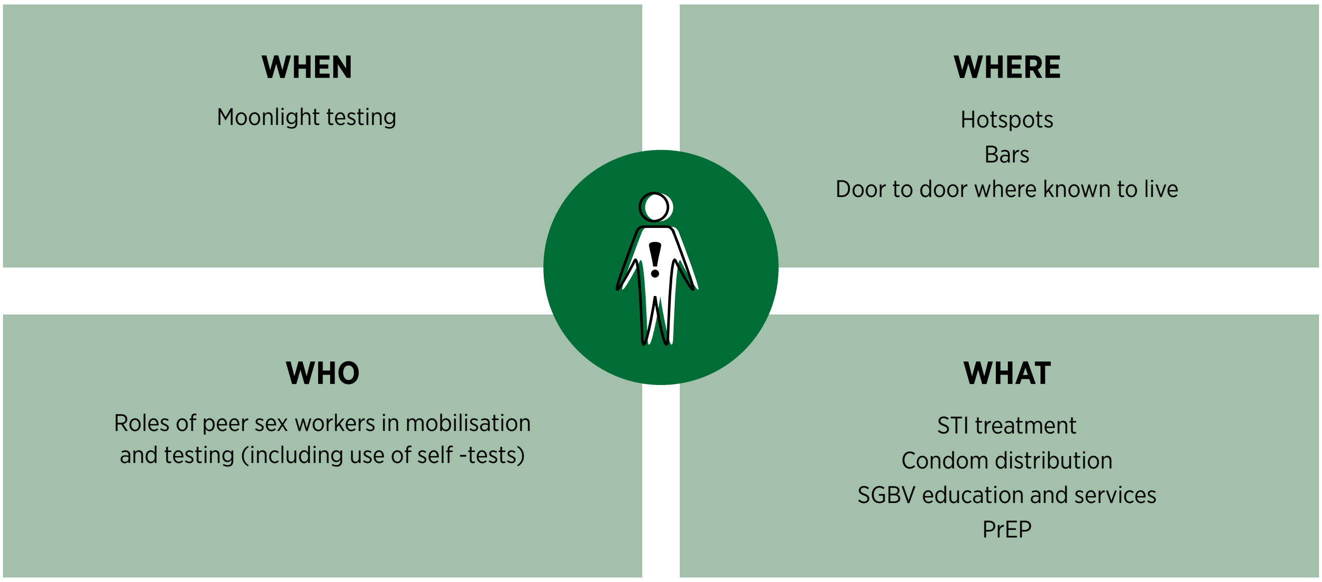

#### 2.2.11 Quality assurance for HTS

Ensuring quality assurance both internal and external for all HTS sites is the responsibility of the district and provincial laboratory scientists.

A clear schedule of when quality control (QC) samples are to be sent to the facilities (this must include all decentralised sites) should be made. In addition, regular ongoing supervision of HIV testing sites and competency assessments of personnel performing HTS is critical to ensuring that high-quality services are being offered in the programme. As part of the QA system, the district/supporting laboratory scientist should periodically (quarterly) carry out support and supervision visits to testing facilities. Full details regarding QC can be found in the Zimbabwe 2014 HTS Guidelines.

#### 2.2.12 Linkage of clients performing HTS to HIV prevention, care and treatment

It is no longer good enough that clients just test for HIV. It is important that those who test HIV positive are linked to care and treatment services and that those who test HIV negative are linked to preventive services to ensure that they remain negative. Systematic reviews have revealed some interventions that can improve linkage. These include involving community outreach workers to identify those lost to follow up, and linking with expert clients along with mobile-health strategies.

##### Clients testing through facility HTS

All clients who have been tested for HIV should be referred for the appropriate post-test services at the same facility or another facility if the required service is not available. If tested in the same facility, clients should either be referred to the OI unit or be immediately assigned an OI number, a patient care and treatment booklet opened, basic HIV and ART education started, and have clinical and psychosocial readiness assessed (Section 2.4.3-2.4.5). Where clients are tested in different areas of the hospital and referred to HIV prevention, care and treatment services, the client should be referred on the same day and accompanied if possible to ensure that they are linked to care. A checklist of all those who tested positive from each department should be kept and reviewed with the OI clinic at the end of each week to see if they have linked to care.

##### Clients testing through community HTS

Where community testing is offered, a number of steps should be put in place to strengthen linkage. Clients should be offered different local options for sites offering HIV prevention, care and treatment and should be free to make an informed choice. The client must be given clear written instructions and an **AIDS and TB referral slip** (Appendix 1) in order to access care at the facility of their choice. They should also be asked for their consent to give their telephone number and be linked to a community health worker or expert client to facilitate tracing.

The community HTS provider should keep a clear record of HIV-positive clients to be followed up. The HTS provider should contact or visit the referral sites to confirm enrolment within an agreed time (ideally within one month) or, with the client's consent, they may be followed up by phone or by a CHW or other community-based cadre.

##### Reference materials

Zimbabwe National Guidelines on HIV Testing and Counselling 2014.

HTS Guidelines for Children and Adolescents 2014.

##### Table 9: Summary of differentiated HIV testing services

| WHEN | WHERE | WHO | WHAT |
|---|---|---|---|
| HTS should be available in all facilities during government opening hours. HTS should be available 24 hours (overnight and weekends) for facilities providing maternity and inpatient care. | Facility-based testing:<br><br>PITC should be offered at the point of entry in all facilities. Entry points should include OPD, IPD (including malnutrition and paediatric wards), TB, STI, MNCH.<br><br>Facility- and community-based index client testing should be offered from all facilities.<br><br>Targeted (sub-population based) community-based outreach testing should be offered at least once per quarter from all facilities. | All cadres of existing health care workers should be eligible to be trained to perform HTS.<br><br>Every facility must ensure that there is always a HCW on duty who has been trained to perform HTS.<br><br>HIV self-testing may be used both at the facility and in community-based approaches. | Integrated screening approaches should be implemented in community testing strategies. This may include HIV testing, TB and STI screening, blood pressure, blood glucose checks and nutrition assessments. |

#### Special considerations for children and adolescents

| WHEN | WHERE | WHO | WHAT |
|---|---|---|---|
| Provision for testing outside school hours (late evening or Saturday morning) and outreach to schools. | Routine opt-out facility-based testing should be offered. The HTS screening tool may be used to identify those most in need of testing for children older than five years.<br><br>Targeted outreach testing to schools, orphanages and during EPI should be included in outreach planning. | Expert adolescent and young adult peers should be trained to mobilise adolescents for testing.<br><br>All cadres of existing health care workers should be trained to prepare DBS samples for EID testing.<br><br>All facilities should ensure that there is always a HCW on duty who has been trained to prepare DBS samples for EID testing. | Integrate EID DBS into outreach EPI services.<br><br>Integrate SRH education and services into testing services for adolescents. |

#### Special considerations for key populations

| WHEN | WHERE | WHO | WHAT |
|---|---|---|---|
| Key populations should be consulted to determine the most appropriate time to offer community- or facility-based HTS, e.g., moonlight testing for those in sex work. | Districts should map the locations where specific key populations will access HTS and offer targeted outreach testing from the facility serving the defined location. | Key population peers should be trained to mobilise their communities to access HTS and other services.<br><br>HIV self-testing should be considered for testing and re-testing in key populations. | Key populations should be offered an integrated package of services with HTS. For example, for female sex workers:<br><br>• HTS<br>• Condom distribution<br>• Family planning<br>• STI screening and treatment<br>• SGBV services<br>• Prevention services (PEP or PrEP) |

### 2.3 Differentiated HIV prevention strategies

The provision of prevention strategies for HIV should take a combination approach, including HIV testing services (HTS), use of male and female condoms, lubricants, ART for HIV-positive partners in sero-discordant couples, voluntary medical male circumcision (VMMC), PreP and STI prevention and management. This section will focus on the programmatic strategies that may be employed in the implementation of VMMC, PrEP and condom distribution using the building blocks approach.

#### 2.3.1 VMMC

VMMC has been shown to reduce the risk of female-to-male sexual transmission by up to 60%. As part of a comprehensive HIV prevention package, WHO recommended the inclusion of VMMC in 2007 and Zimbabwe has been identified as one of 14 priority countries for the scale up of VMMC. To achieve the highest impact, demand generation should be focused in the age group of 13-29 years, although circumcision is offered from the age of 10.

##### Where is VMMC offered?

VMMC may be offered via a:

- Static model, utilising existing permanent facilities
- Outreach model , temporarily utilising existing structures that are not routinely offering VMMC, such as rural health centres
- Mobile model, utilising temporary structures, such as tents. All resources are obtained from the static site, including health personnel.

##### When is VMMC offered?

VMMC should be available every working day from the static sites. The outreach and mobile models may be used during periods of high demand, visiting a static site 2-3 times per month or establishing a mobile site for a one-week period and then moving to another site.

##### Who provides VMMC?

Health care workers who may be trained to perform circumcision include doctors and nurses. Training nurses to perform VMMC will significantly support the scale up of services.

The team structure for each of the three models is as follows:

- Static site: two trained circumcisers, three nurses, one theatre assistant and one receptionist
- Outreach and mobile model: one trained circumciser, three nurses, one theatre assistant, one receptionist and one driver. Staff should be dedicated to VMMC activities for the duration of the outreach activity.

##### What services are offered during VMMC?

Circumcision may be performed using either a surgical or non-surgical technique (PrePex device). All clients are offered HIV testing prior to circumcision, along with counselling to ensure that the client understands the procedure and the symptoms and signs of complications.

##### Reference materials

Zimbabwe Policy Guidelines on Voluntary Medical Male Circumcision.

Accelerated Strategic and Operational Plan 2014-18 VMMC.

#### 2.3.2 Pre-exposure prophylaxis (PrEP)

WHO defines PrEP as the use of antiretroviral drugs before HIV exposure by people who are not infected with HIV in order to prevent the acquisition of HIV. PrEP has been included in the Zimbabwe combination prevention strategy and will be implemented in a phased approach. PrEP is taken as dual therapy (TDF+FTC or 3TC) daily during periods of risk and does not have to be for life. If taken strictly PrEP may reduce the risk of HIV infection by 90%, but works best as part of other HIV prevention measures; it does not protect against STIs or pregnancy.

Population groups that may be at higher risk of infection may include:

- Female and male sex workers
- Sero-discordant couples (the HIV-sero-negative partners)
- Adolescent girls and young women
- Pregnant women in relationships with men of unknown status
- High-risk men (MSM, prisoners, long-distance truck drivers)
- Transgender people.

A set of screening questions to identify those clients who may benefit from PrEP can be found on page 31 of the Job Aide.

##### Where will PrEP be offered?

PrEP will be phased in across the country, and it should be offered from all facilities. Where there is provision of ART refills through mobile outreach, PrEP may also be provided through this service delivery model.

##### When is PrEP offered?

PrEP should be offered during routine clinic opening hours. However, where the time for ART service delivery has been adapted for specific sub-populations, similar strategies should be applied for PrEP. Examples are providing PrEP after school hours for adolescents or during specific afternoon or evening hours for sex workers, depending on an assessment of the key population's preferences.

##### Who can provide PrEP?

Any health care worker who is trained to provide ART can also provide PrEP.

##### What is offered?

After initiating PrEP, the client should be reviewed after one month to monitor adherence and side effects, as well as for resupply of medicines; thereafter, the client should be reviewed every three months. HIV testing should be performed every three months. The client should also be linked with other combination prevention strategies, such as VMMC and condom use.

##### Reference materials

Guidelines for Antiretroviral Therapy for the Prevention and Treatment of HIV in Zimbabwe 2016.

#### 2.3.3 Condoms

The national comprehensive condom programme is aimed at increasing availability, access and informed demand for male and female condoms. Both male and female condoms should be available and offered at any interaction a client has with the health system. This should be both at the facility and during community-based activities, such as HTS. Condoms may be distributed by health care workers, through community distributors or from strategically placed distribution points. Condom distribution strategies should be adapted according to the sub-population, such as finding youth-friendly distribution strategies, offering a range of condom brands, and, for key populations, ensuring adequate distribution of lubricants alongside condoms.

### 2.4 Differentiated ART Initiation

#### 2.4.1 The four steps of ART initiation

Once a client has tested HIV positive at a facility or when a patient who has tested HIV positive elsewhere links with a facility, they should be immediately registered and an OI number issued. All clients who are HIV positive are now eligible to start ART under the "Treat All" guideline.

> Although all clients are eligible for ART, before initiation is performed, a readiness assessment must be made. This includes clinical readiness and psychosocial readiness. The key factor differentiating how ART initiation is performed is the clinical characteristic of the client: whether the client is well (asymptomatic and/or with high CD4) or presenting with advanced disease (symptomatic and/or with low CD4 <100 cells/mm$^3$).

Once registered, the client passes through a number of steps with both the clinician and counsellor to determine their readiness for ART initiation. These steps should be done as soon as possible, preferably on the same day as testing or when the client links to the facility of their choice. The steps that must be carried out include:

**Step 1:** Provision of basic HIV and ART education (clinician, primary counsellor or expert client)

**Step 2:** Clinical readiness: history, physical examination, baseline laboratory investigations, including investigation of those with advanced disease (clinician)

**Step 3:** Psychosocial readiness assessment (clinician or primary counsellor)

**Step 4:** Treatment plan (clinician)

##### Figure 6: The four steps of differentiated ART initiation

**STEP 1 — HIV and ART education**

Give basic HIV and ART education

(See Page 106-124 Job Aide)

**STEP 2 — Clinical readiness,** clinical history, examination and investigation

**Clinically ready, asymptomatic** and POC baseline CD4 >100 cells/mm$^3$ (or CD4 screening at baseline is not available)

**Clinically NOT ready**

Asymptomatic and POC baseline CD4 <100 cells/mm$^3$

**OR**

Symptomatic and requires further investigation and clinical management

↓

Screen for Cryptococcal disease

Screen for TB according to the TB screening algorithms

Manage clinical condition

**STEP 3 — Psychosocial readiness**

Complete counsellors' ART initiation checklist

Page 55 Job Aide

**STEP 4 — Treatment plan**

**Clinically and psychosocially ready**

Offer rapid initiation

**Psychosocially ready but clinical reason for delay** (e.g TB , cryptococcal disease) delay ART initiation according to clinical guidelines

**Clinically ready but psychosocial reason for delay**

Give further counselling

Use counselling tools and link to expert client where possible

Aim to start within one week of diagnosis

#### 2.4.2 Step 1: Basic HIV and ART education

All clients should be referred for the session of basic HIV and ART education on the day of testing or first day of linkage to a facility. In addition to using the counselling tools on page 110-118 of the Job Aide, the soldier game may also to be used to strengthen understanding of basic HIV and ART concepts (see soldier game, SOP, page 106 of Job Aide). The counsellor must assess how much information the client is able to absorb in the initial interaction. For some, it may be appropriate to complete basic HIV education and ART education on the same day. For some, these sessions should be scheduled two to three days apart. It is important to remember that clients may travel long distances to attend the clinic and may have to take time off work. Where expert clients are present in the community, follow-up counselling covering the information in these basic sessions may also be performed in the community with the consent of the client.

Counsellors should write their findings in the notes section of the patient care and treatment booklet. Clinicians should refer to these notes and discuss face to face with the counsellor if problems have been identified.

##### Special consideration for basic HIV education for children and adolescents

The way that basic HIV and ART education is carried out will depend on the age of the child and the decision made with the guardian around disclosure. The guardian should be given all the education, as outlined in the disclosure session guides (Job Aide, pages 125-137). Younger children should be included in the sessions and can participate without having to name HIV. Older children and adolescents should be disclosed to at least by the age of 12. If this can be supported early in the process of their diagnosis, this has been shown to support adherence.

#### 2.4.3 Step 2: Clinical readiness

At the first assessment a full history, clinical examination and request for baseline investigations should be carried out.

##### History

A full medical history should be taken at the first visit.

###### *History of present illness*

Check for the main problem of today. For each problem, further enquire: Since when? Where? How? Any aggravating conditions? Note a timeline of the complaint.

Are there any associated constitutional symptoms, e.g., loss of appetite, loss of weight, night sweats?

###### *Past medical history*

**TB:** Past history of TB: Was treatment completed? Has there been any recent/current contact with a TB case (drug sensitive or drug resistant)?

**OIs:** Has the client been treated for or admitted for any staging OIs?

**Other conditions**, e.g., epilepsy, diabetes, hypertension. This is important as symptoms may overlap and they may predispose the client to a higher risk of side effects, e.g., diabetes and hypertension may mean a higher risk of renal side effects with TDF; there may also be significant drug interactions that must be taken into consideration.

**Previous psychiatric disorder** – anxiety and depression

###### *Medicine history*

Is there any prior exposure to ARVs (including PMTCT, PrEP and PEP)? The client may have previously been initiated on ARVs in another setting.

In particular, look out for other nephrotoxic drugs, e.g., long-term NSAIDS or aminoglycosides. Avoid TDF use with these drugs, especially if creatinine cannot be monitored, and any drugs that may interact with ART (anti-epileptics, oral contraceptives).

###### *Family history*

Does the client have a partner or children? Have they all been tested?

###### *Social history*

What is the client's support network at home? Are they employed? Have they been able to maintain their job? Is there any history of alcohol or other substance abuse?

###### *Review of systems*

Any weight loss noticed by the client

Any rashes; any history of a painful blistering rash (herpes zoster) = Stage 2

Any diarrhoea (more than a month = Stage 3)

Any recurrent fever (more than a month = Stage 3)

Screen for STIs

Screen for TB symptoms using the Zimbabwean TB screening tool

Screen for symptoms of depression

A full antenatal obstetric history should be taken at the first visit.

Record birth history and history of immunisations.

#### Examination

**All clients should be examined from top to toe at the first clinical assessment.** The purpose of the examination is to stage the client and to detect any signs of possible OIs or HIV-related cancers prior to initiation of ART in order to avoid IRIS. A swollen cervical lymph node may be the only sign of TB in a client with HIV and may go unnoticed if the client is not examined.

**To examine the client properly, you must undress them and examine them on the couch.** You will need a torch/flashlight to properly examine the mouth and a stethoscope to examine the chest.

##### *What is the client's general condition?*

Check weight and height. If there is any previous weight documented in the client notebook, have they lost more than or less than 10% (<10% Stage 2: >10% Stage 3)?

Check the vital signs (pulse, respiratory rate, oxygen saturations if possible, blood pressure and temperature). Is this client stable or unstable? (If unstable, consider the need to refer.)

Assess for any pallor (anaemia due to TB or HIV itself) or jaundice.

Assess from top to toe the skin – herpes zoster acute or scars; PPE; fungal rashes.

Examine the mouth and palate for signs of oral thrush, oral hairy leukoplakia or Kaposi's sarcoma.

Examine the lymph nodes in the neck, above the collar bones under the armpits and in the inguinal region to see if they are enlarged.

Examine the chest for any signs of respiratory distress or focal respiratory signs.

Examine the abdomen. Are there any masses or tenderness?

Examine the genital area for STIs.

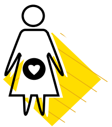

In pregnant women a full obstetric examination will need to be performed.

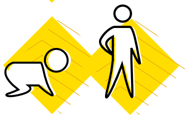

In children assess growth and developmental milestones. Remember to perform an ENT examination in children. Chronic otitis media is common in children with HIV and, if untreated, can lead to long-term hearing problems.

#### Baseline investigation

##### *Essential:*

Repeat HIV testing on the day of ART initiation on a second sample and, ideally, by a different HCW.

##### *It is preferable in most instances to perform the following baseline tests/measurements:*

- Full blood count (especially if zidovudine will be used)
- Serum creatinine test (if tenofovir will be used)
- Baseline CD4. NB: A baseline CD4 test is recommended to determine the degree of immune suppression of a patient to inform differentiated ART initiation for the patient
- Pregnancy test
- Alanine transaminase test (ALT)
- Mantoux test (useful in children)
- GeneXpert test or chest X-ray (to exclude TB)
- Blood pressure measurement.

##### *If possible, perform the following tests also prior to commencing ART:*

- Syphilis serology test
- Hepatitis B and C virus screening.

#### 2.4.4 Step 3: Psychosocial readiness assessment

##### Assess readiness to start

- Ask patient what would be 3 important reasons for them stay healthy and alive.
- Assess willingness to start ART.

Recap knowledge of ART education session (Job Aide, page 113). Can the client describe:

- Routes of transmission of the HIV virus and ways to prevent transmission, including how to use condoms
- The evolution of HIV infection with and without any ARV treatment
- What happens if ARVs are not taken as prescribed (development of resistance and treatment failure)
- What we want to see happening to viral load (going down)
- How to recognise the red-flag symptoms and signs (OIs and side effects) that they must come immediately for consultation
- Why they are eligible to start ART today and that ART treatment is lifelong
- For each of the drugs, the name, frequency and side effects that might occur
- Use of herbs: Why it's important to stick to ARVs as a treatment
- Why it is important to come on the review date given and what to bring (all remaining medications)
- What to do in case of travel.
- Plan with patient how they will take their medicines:
    - What would be the best timing for you to take your drugs, taking into account your daily habits?
    - What tools will you use to remind yourself to take your drugs (alarm)?
    - Where will you store your drugs?
    - Where will you keep extra doses in case you are out of the house?
    - How will you manage missed doses?
    - What will you do in case of side effects?

##### *Explain follow-up plans*

Follow up is quite intense at the start of treatment ( D14, M1, M3) but will become less frequent once the patient is stable. Options for long-term follow up will be discussed at later counselling sessions.

Ask their consent to be called or traced if they miss an appointment.

Findings from this assessment should be documented in the notes section of the patient care and treatment book.

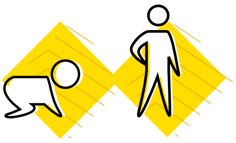

##### Special consideration for readiness assessment in children and adolescents

Younger children are dependent on their caregiver for administration of ART. It is therefore essential that these caregivers understand the importance of ART administration for the child. A very common reason for poor adherence in children is when there are multiple caregivers, some of whom have not been educated on the importance of taking the medication. Depending on the age of the child, involve the child in the planning of how and when they will take their drugs.

***Answer the following questions:***

- Is there an identified dedicated caregiver? If possible, two caregivers should be identified. If the child is regularly left in another home, try to include that caretaker in the preparation.
- What is the current disclosure status of the child? If not disclosed, a clear plan on disclosure should be made with the caregiver. Partial disclosure should be achieved at least by age nine and full disclosure should be achieved at the latest by age twelve (Job Aide, page 125-137).
- Is the child able to swallow the dispersible tablets or use the pellets correctly?
- If using lopinavir/ritonavir syrup, is the child able to tolerate the medication?

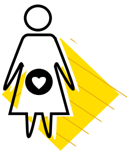

##### Special considerations for pregnant and breastfeeding mothers

The content of the initiation counselling session should be prioritised around: 1) the motivation for taking medication (to keep the baby negative and, in the longer term, to keep the woman healthy to care for her baby); and 2) how to take the medication. At subsequent sessions, ongoing counselling and assessment of HIV and ART knowledge must be further developed. In addition, at a later date, the woman must be counselled on planning a safe delivery, use of the NVP, AZT and cotrimoxazole syrups that her baby will need, testing her baby, and infant feeding options.

**At first visit, emphasise that this treatment is to keep her baby HIV negative and to keep her healthy in order to look after the baby.** If the woman has concerns about life-long treatment, these should be further discussed during follow-up sessions, but now encourage her that the immediate motivation is to keep her baby negative.

**Lack of disclosure** is a very common reason for pregnant and breastfeeding women not to take their medication. Start to discuss options for how she might disclose to her partner.

**For pregnant or breastfeeding women, the following additional aspects related to prevention of mother-to-child transmission should also be explained.**

*Explain ways of transmission of HIV*

- Explain different ways that a mother can infect her child: during pregnancy at delivery or during breastfeeding.
- Explain chances of transmission from mother to child. With the correct follow up on ART, there are high chances that your baby will be HIV negative.

*Give brief PMTCT ART education*

Finding out you are HIV positive is a lot to deal with today, but it is important that we already speak for a moment about the health of your baby. You could have a HIV-negative baby if you take the right precautions:

1. **Start ART as soon as possible:** HIV has no cure, but there is a treatment to control HIV in your body. All pregnant women are to start this treatment as soon as possible as this gives a high chance of preventing the transmission of the virus from you to your baby. We invite you to start taking the treatment today, but it is up to you to decide if you feel ready for this.
2. **Delivery in a health facility:** It is safest to go to a health facility for delivery and inform the staff that you are HIV positive; then the staff will be able to take all precautions to protect the baby during delivery.
3. **Correct feeding of the baby:** After delivery, it is important to only give breast milk for the first 6 months. After 6 months, other foods can be introduced, but continue breastfeeding until at least 12 months.
4. **Correct treatment of the baby:** The baby will be given different protective syrups (NVP, AZT CTX) right after birth.

Through these four actions, you will protect your baby and the chances of him or her becoming infected are very small. Today we will focus on how to take the treatment correctly and we will cover other topics at later sessions. We will make a plan together to enable you to take the medication correctly.

**Make a plan with the patient on how to take ARVs** as for the general population, but specifically ask about: What are your travelling plans in the coming months (mobility issues, "Kusungirwa", etc.)?

**Make a plan for disclosure and testing of partner.** Discuss strategies to get their partner to come for testing (invitation letter from the clinic, communication with partner, re-test both partners together) and how she may be able to disclose her status.

**Ask them if they have any questions** and explain that they are going to be booked for a second session at week 2 on ART.

Aim to link the woman with a **community health worker or PMTCT "champion"** who can support them in the community with the consent of the woman.

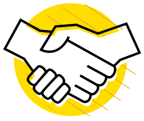

Ask their consent to call or trace them if they miss an appointment.

#### 2.4.5 Step 4: Treatment plan

**Option 1:** If the client is both clinically and psychosocially ready, initiate ART on the same day if they agree. Cotrimoxazole should be initiated according to the criteria in the clinical guidelines, two weeks after the initiation of ART.

**Option 2:** If the client is clinically ready but not psychosocially ready, book the client for a further session in the next one to seven days according to what is convenient to the client. **At each subsequent appointment, assess readiness and utilise tools, such as the soldier game and flip charts.** Linking the client with an expert client or other peer on ART may also address issues that may be a barrier to accepting ART. If possible, aim to initiate ART within one week after the patient has tested positive in the facility or has linked to care.

**Option 3:** If client is not clinically ready:

- Which conditions do I need to treat today before I start ART?
- What conditions need further investigation today?
- If CD4 is available at first visit, **clients with a CD4 <100 cells/mm$^3$ need special attention.** They are classified as having advanced disease. These clients need particular attention to screening for TB, examination for Kaposi's sarcoma and enquiry about any visual problems (potential CMV). These clients need to have blood sent to the laboratory to be screened for cryptococcal antigen with CRAG testing. For further details on the clinical management for pre-emptive treatment of cryptococcal disease, see the *Guidelines for Antiretroviral Therapy for the Prevention and Treatment of HIV in Zimbabwe* and page 102 of the Job Aide.
- Clients with TB should be initiated on ART after at least two weeks of TB treatment. Those with a CD4 less than 50 cells/mm$^3$ should receive ART within the first two weeks of TB treatment.
- Clients diagnosed with cryptococcal meningitis should take two weeks of amphotericin B before initiation of ART. If amphotericin B is not used, at least four weeks of fluconazole treatment should be taken before ART is initiated.

#### ART initiation checklist for the clinician

**Step 1:** Has HIV testing been confirmed with a second test, on a different sample ideally by a different health care worker? This should be documented on page 2 of the patient care and treatment booklet.

**Step 2:** Does the client have sufficient understanding about HIV and ART, and is the client psychologically ready to start ART? The clinician needs to review the counsellor's notes from the preparatory sessions to ensure that there are no outstanding issues that may affect initial adherence (severe depression, denial, plans to travel). For children, does the caretaker fully understand their responsibility for providing the child's ART?

**Step 3:** Screen again for TB. This is essential to avoid episodes of TB IRIS.

> Clients with TB co-infection should start TB treatment first. ART should be started after two to eight weeks of TB treatment. Those with a CD4 less than 50 cells/mm$^3$ should receive ART within the first two weeks of TB treatment.

**Step 4:** Ensure that all OIs and other infections have been adequately screened for (cryptococcal disease if CD4 <100; TB; STI) and treated.

> Clients diagnosed with cryptococcal meningitis should take two weeks of amphotericin B before initiation of ART. If amphotericin is not used, at least four weeks of fluconazole treatment should be taken before ART is initiated.

**Step 5:** Examine the client.

**Step 6:** Review the baseline laboratory tests – if performed – to decide on the choice of regimen.

**Step 7:** Choose a regimen according to the clinical guidelines for antiretroviral therapy in Zimbabwe.

**Step 8:** Review potential side effects of the medication with the client and ensure that they know what symptoms to report to the clinic early (Table 10). Be especially alert to side effects of TDF in the elderly, diabetic or hypertensive client.

**Step 9:** If all of the above steps have been checked and the client is ready, initiate ART.

**Step 10:** Enter the client in the chronic ART register or EPMS.

##### Table 10: Symptoms and signs that clients must report

| IMPORTANT SYMPTOMS OR SIGNS A CLIENT SHOULD REPORT TO THE CLINIC | |
|---|---|
| Cough, night sweats, weight loss | Possible TB or other chest infection |
| Severe headache | Possible new OI such as cryptococcal or TB meningitis |
| Breathlessness, dizziness | Possible chest infection or anaemia if on AZT |
| Diarrhoea or vomiting | Possible new OI or side effect of medication (Lop/rit) |
| New rashes | Possible side effect of NVP and EFV (less common with EFV)<br><br>Flare up of existing PPE or fungal rashes |
| Facial swelling or ankle swelling | Renal dysfunction, possible side effect of TDF |
| Change in how they are urinating (especially if urination reduces or stops) | Renal dysfunction, possible side effect of TDF |
| Severe sleep disturbance; change in behaviour | Possible side effect of EFV (NB: Before attributing behaviour change or confusion to EFV, ensure that the patient has been investigated for OIs, i.e., has had a lumbar puncture performed) |

#### 2.4.6 ART follow-up schedule

##### ART follow-up schedule

| | D0 | WK 2 | MTH1 | MTH 3 | MTH 6 | MTH 7 | MTH 9 | MTH 12 | LONG-TERM FOLLOW UP |
|---|---|---|---|---|---|---|---|---|---|
| **CLINICAL** | Complete ART initiation checklist | If on a NVP-based regimen or<br><br>Had same-day initiation | | | | Review first VL result CHOOSE REFILL OPTION. | REFILL OPTION | | If patient remains virologically suppressed continue with patients refill option.<br><br>3-monthly supplies of ARVS and cotrimoxazole should be given. If monitored with viral load, see for clinical review yearly.<br><br>If no viral load, see for clinical review every 6 months.<br><br>If not virologically suppressed follow the viral load algorithm. |
| **COUNSELLING** | Complete readiness assessment | | | | Give viral load key messages.<br><br>Discuss refill options. | | | | Adherence should be assessed by the nurse at each clinical visit. At refill visits, peer support for adherence is given by the group members if the refill system is in a club or CARG.<br><br>After month 6, clients should see the counsellor only if a red-flag sign is picked up by the nurse, or if client attends late, or has a high viral load. |
| **LABORATORY** | CD4, Cr if on TDF, Hb if on AZT | | Hb if on AZT | | VL | | | VL | Viral load yearly.<br><br>If no viral load available, CD4 6 monthly.<br><br>Creatinine (TDF), HB (AZT), ALT (NVP) should be checked if there is any suspicion of side effects. |

#### Special considerations for ART follow up for children, adolescents and pregnant women

| | FIRST YEAR | LONG TERM FOLLOW UP |
|---|---|---|
| **PREGNANT WOMEN** | Pregnant or breast feeding women initiating ART as part of PMTCT undergo rapid initiation on the same day as testing.<br><br>They should then be seen at week 2, month 1 and then monthly while they are attending for ANC and PNC/bringing the exposed baby monthly. As described in Section 1.4. this should be offered as an integrated one-stop service.<br><br>A counselling session is given at week 2 to ensure more detailed HIV and ART education is given. Counselling follow up is adapted to the changing motivation for taking ART over time; discussion on delivery; infant testing and infant feeding should be included at the appropriate time. | Once the baby is diagnosed HIV negative (6 weeks post cessation of breastfeed-ing), the woman can decide which refill option she would like to consider for future long-term follow up. |
| **CHILDREN** | Infants up to 2 years old should be reviewed monthly. Thereafter children should be seen every 3 months. **THIS IS BECAUSE THE DOSE MUST BE ADJUSTED FOR THE WEIGHT.**<br><br>For children, follow the adult counselling schedule until month 6 and then see them 3 monthly until full disclosure is achieved.<br><br>Plan to group your children on the same day each week/month. This automatically allows for peer support to enhance adherence. | Until on adult doses, children should be seen every 3 months.<br><br>Once on adult doses, follow up as for adolescents.<br><br>Children should continue to see the counsellor every 3 months until full disclosure is achieved. |
| **ADOLESCENTS** | If starting on adult doses, adolescents can follow the routine follow-up schedule as above for the first year.<br><br>The counselling content should be adapted to their particular needs (SRH, coping with school, starting new relationships etc.). | Until fully disclosed (goal by age 12) con-tinue to see clinician and counsellor every 3 months. Once disclosed and on adult doses, adolescents should be seen clini-cally once every 6 months, unless serious psychosocial issues are identified.<br><br>Offer a facilitated group refill (Page 72). |

#### 2.4.7 Follow up of a client on ART by the counsellor

After initiation, the client should see the counsellor at week 2, month 1, 3, 6 and 7. These sessions **could be done as a group if a few clients with the same duration on ART are attending the same day or as individuals.** The client should see the counsellor before the clinician, and the counsellor should document their findings in the notes section of the patient care and treatment booklet. The counsellor should be guided by the client about the specific challenges they would like to discuss, but it is also important that some key issues are covered for all by the time they are six months on ART. These topics include:

- Planning for travel
- Family planning; and planning your family
- Understanding treatment failure and interpretation of viral load results. This must be reinforced prior to the viral load being taken at month 6
- Understanding the need for lifelong ART related to cultural and religious beliefs
- Understanding and choosing a refill option for long-term follow up. This should be introduced during the general ART education, but a decision should be made when the first viral load result is reviewed.

##### Special considerations for the counselling follow up of pregnant and breastfeeding women

A counselling session **at week 2** is given for pregnant and breastfeeding women. Additional topics related to their stage of PMTCT should be incorporated: **planning a facility-based delivery; NVP and /or AZT use; CTX use; DBS testing; and infant feeding advice.** There are also some key **transition points in the journey of PMTCT** where key messages should be emphasised:

- Planning where the woman will deliver or, if she will travel away from the facility. Consideration of cultural practices, such as Kusungirwa, must be discussed and, if needed, extended drug supplies given or referral made to another ART site.
- Exclusive breastfeeding for six months is the recommended infant feeding option. When the woman is seen post delivery, it is very important to explain that the medication she is taking is making her **breast milk safe.** The chances of transmitting HIV to her baby if she takes the medication daily are very low. So her motivation for taking the medicine is still to keep her baby negative.
- **Family planning options** should be discussed.
- She should be reassured that the **medication she is taking is not harmful** to the baby.
- During the subsequent sessions, further discussion about lifelong treatment can be developed. **When she is about to stop breastfeeding** is an important stage as, prior to this, she has the additional motivation for treatment for keeping the baby negative. Now the treatment is for her own health. She should also understand that continuing on the ART **will protect a baby in any future pregnancy**.

##### Special considerations for ART follow-up counselling for children and adolescents

In addition to adherence counselling, children and their caregivers need to be supported through the process of disclosure. Evidence suggests that older children who do not understand about their HIV status have worse adherence and retention. Disclosure is a process. Partial disclosure (where the child understands what is going on in their body but does not name the disease) should start as soon as the child is able to understand simple story lines and should be achieved at least by age nine, with many children achieving this earlier.

Full disclosure, where the child also names the disease, should be achieved at the latest by age 12. For some children, full disclosure can be achieved earlier. Although it is encouraged that the caregiver discloses, the health care worker must proactively initiate and guide the process. If full disclosure has not happened by the age of 10, this case must be taken seriously and a plan made with the guardian. Pages 125-137 of the Job Aide gives an outline for these disclosure sessions and some picture tools that may be used.

##### Reference materials

Guidelines for Antiretroviral Therapy for the Prevention and Treatment of HIV in Zimbabwe 2016.

Zimbabwe EDLIZ 2015 (7th edition).

#### Table 11: Summary of differentiated ART initiation

| WHEN | WHERE | WHO | WHAT |
|---|---|---|---|
| All clients should be assessed for **rapid initiation**.<br><br>If client is not ready on the same day for clinical or psychosocial reasons, then ongoing medical investigation and/or counselling support should be provided with the aim of **initiating ART as soon as possible, ideally within 1 week unless there is a clinical contraindication.** | ART initiation is primarily performed **at the facility.**<br><br>Where **mobile outreach** is being performed regularly to a site, ART initiation may also be considered. | Initiation may be performed by any **trained health care worker** (doctor, AMO, clinical officer, nurse). | **Step 1:** Provision of basic HIV and ART education<br><br>**Step 2:** Clinical readiness: including assessment of clients with advanced disease (including screening for cryptococcal disease if CD4 is $< 100$ cells/$mm^3$)<br><br>**Step 3:** Psychosocial readiness assessment<br><br>**Step 4:** Treatment plan |

### 2.5 Differentiated ART delivery

#### 2.5.1 Deciding how to differentiate ART delivery

Differentiated ART delivery is a component of differentiated service delivery. Using the elements (clinical characteristics, sub-population and environmental factors) and building blocks (when, where, who and what), as described in Section 2.1, a model of ART delivery can be built that responds to a particular challenge being faced by the health system or client.

The decision on what strategies to prioritise in a particular site should be led by the district health management team. The team should define what the challenge is for the health care workers and for the clients. These challenges may be different in different facilities in a district. The tool in Appendix 4 may guide your assessment. Certain refill options address specific challenges and should be selected accordingly. Some models give the benefit of peer support; however, not all clients wish to disclose and be part of a group. Hence, alternative and efficient individual models should also be considered. The specificities of the refill options are highlighted in Section 2.5.4.

Hence, any intervention should be a response to a specific local challenge faced by the health system or by clients. The same option may not apply to all sites, and options to address clients' differing preferences should be considered.

The following steps provide a guide to district health management teams for planning how to differentiate ART delivery in their context.

##### Figure 7: Decision framework for differentiating ART delivery

**STEP 1**
**Situation Analysis**

- Use the tool in Appendix 4 to guide your assessment
- Assess facility level retention and workload data.
- If possible disaggregate data by age and subpopulation
- Assess challenges being faced by your health care workers
- Assess challenges being faced by clients in your facilities

**STEP 2**
**Define challenges for each facility**

- What are the common challenges
- What are challenges specific to certain facilities or client subpopulations

**STEP 3**
**Define the priority subpopulation**

- Define the priority subpopulation/s for whom ART should be differentiated
- What is the districts priority in the next six months?

**STEP 4**
**Design a model of ART delivery**

**Ask the following questions**

- Is the maximum refill (3 months) being offered?
- Could ART be offered on additional days of the week?
- Could opening hours be extended for provision of ART refills?

**Design a model of ART clinical and counselling follow up:**

Clinical consultation.
When | Where | Who | What

Counselling consultation.
When | Where | Who | What

**Define steps for ART refills:**

- For **stable clients** choose a refill option (Section 2.5.4) that addresses the local challenges.
- For **other subpopulations** design a refill model to address their particular needs (Section 2.6).

When | Where | Who | What

**STEP 5**

- Implement the differentiated ART delivery model
- Evaluate it's impact
- Consider further adaptations to differentiated ART delivery to address other identified challenges

#### 2.5.2 Clinic systems to be implemented for all differentiated ART delivery models

##### Client triage and clinic flow

Identifying what services the client is attending for will help ensure that clients receive the correct package of services. The Electronic Patient Management System (EPMS) appointment list should indicate:

- the type of refill,
- who is due a viral load,
- and clients whose last viral load was >1000 copies/ml.

In the clinic diary and patient-held book, clinicians should write the return date and what the client will be attending for, e.g., refill or clinical review and viral load. This will facilitate the health care worker allocated to triage clients at the next visit.

Clients needing any **blood tests should be identified first** and directed to the allocated member of staff performing tests on that day. Where a client (according to the follow-up schedule, Section 2.4.6) needs to see both counsellor and clinician, they should **see the counsellor first**.

The exact client flow may vary across sites. Each facility should develop a triage system and map out their client flow. This flow should be drawn on a poster and all clinic staff should be made aware of the details.

##### Appointment systems and defaulter tracing

At all steps of the cascade, the ability to identify that a client **has not attended for their appointment** will be dependent on the **use of an appointment system**. In facilities utilising the EPMS, daily lists of clients due that day can be generated. The list should indicate who is due to attend, the refill model they are in, whether VL is due, and if the last VL was >1000 copies/ml. In facilities without the EPMS, **a simple paper diary can be used with assigned codes indicating the reason for the visit.** Clients should be traced three days after their appointment if they have not attended (i.e., this is about tracing clients to prevent them becoming lost to follow up).

##### Standard operating procedure for defaulter tracing

- At enrolment, clients should be asked **if they agree to consent to tracing**. Their decision should be clearly indicated on page 2 of the patient care and treatment booklet.
- **All sites should have an appointment system for HIV-positive clients.** In primary care clinics, all clients should be booked in the same clinic diary. In larger facilities, each clinic (OI, MCH, TB) will have their own appointment diaries.
- **The nurse in charge of the clinic must be clear which staff member** (nurse, nurse aid, PC, receptionist) **is responsible** for updating the diary on a daily basis and for initiating the defaulter tracing process.
- **All clients registered for ART preparation, ART and PMTCT (including the HIV exposed infant)** services should be given an appointment date, which is recorded in the EPMS or clinic appointment diary. In some sites, it may be appropriate to give a booked time (morning or afternoon), as well as a day in order to stagger appointments. If group club refills are implemented, the group number should be recorded in the appointment diary and a booked time for the group allocated.
- The OI number, client's name, telephone number and the reason for the next appointment (clinical consultation +/- counselling, refill for drugs, blood draw for VL) should be listed in the diary.
- The diary or EPMS list can be used to pull the patient care and treatment booklets the day before and also to pre-pack refills in larger sites.
- When the client arrives, it should be marked off in the diary that they have attended.
- At each visit, whoever is registering the client should check that an up-to-date phone number is available and is documented in the EPMS or appointment diary.
- If the client does not attend for their appointment, their patient care and treatment booklet should be kept aside in a tray or shelf allocated for late-attenders. Tracing is not triggered immediately and patients' files coming 1-3 days late should be found in this tray.
- If the client has not attended for three days, the client should be traced. All clients, when first registered, should give consent to be traced.
- For each patient to be traced, an **AIDS and TB programme referral form (Appendix 1)** should be completed and given to the appropriate staff member to carry out the tracing. If just phone tracing is required, this may be done by the nurse; if phone or home visit is required, this may be done by the primary counsellor, village health worker or other CBO/expert client representative. Who is performing the tracing should be clear within the health centre human resource management structure.
- Files for patients being traced should be placed in a "tracing" tray.
- Tracing should be carried out as follows:
  - If a phone number is recorded, phone the client with the clinic phone. If not reachable on first attempt, try again on two subsequent days.
  - If there is no phone number or no response on phoning, proceed to visit at home.
  - If client or relative is not found at home, attempt again after seven days, and then monthly until three months from the referral for tracing.
  - If clients have missed doses they should be reviewed by the nurse or counsellor to ensure strategies are put in place to avoid this again. Once problems are addressed they may continue in their refill option.

- The outcome of the defaulter tracing should be indicated in the diary, the green patient care booklet and ART registers or EPMS. Outcomes of tracing include:
  - In care
  - Lost to follow up
  - Died
  - Moved away not on treatment
  - Official transfer out
  - Self-transfer to another facility; still on ART.

Mobile phone SMS appointment reminders and sending automatic SMS reminders to those who have missed appointments have been shown to improve retention. In sites with the EPMS, this may be considered in the future.

#### 2.5.3 Differentiated ART for stable clients

##### Eligibility criteria for differentiated ART delivery for stable clients

For all the differentiated models of ART delivery for stable clients, the following eligibility criteria should be met.

A stable client on ART (first- or second-line) is defined as someone who:

- Where viral load is available:
  - has no current OIs,
  - has a VL <1000 copies/ml,
  - is at least six months on their current regimen
- Where viral load is not available:
  - has no current OIs,
  - a CD4 >200cells/mm$^3$,
  - been at least six months on their current regimen.

When assessing eligibility for a particular model, a psychosocial assessment should be made. The pros and cons of entering a model for stable clients should be assessed as, in some cases, alleviating the burden of visits may support improved adherence. For some clients with psychosocial issues, the benefits of becoming a member of a group model may also provide additional support for adherence.

##### When do visits happen?

Stable adult clients on ART who are being **monitored with viral load** should be seen for a clinical assessment and repeat VL once every 12 months. Four 3-month refills of ART and cotrimoxazole can be written so that the client attends for three refills before the next clinical visit. Appointment dates for each visit should be recorded in the diary and the EPMS at each refill visit.

Stable adult clients on ART who are being **monitored clinically and with CD4** should be seen for a clinical assessment and repeat CD4 once every six months. Two 3-month refills of ART and cotrimoxazole can be written so that the client attends for one refill before the next clinical visit. Appointment dates should be updated in the diary on the day of each refill.

##### Where do visits happen?

Clinical visits will usually be performed at the facility. Where a mobile outreach model is in place (Page 60), clinical visits may also be performed within such a model.

Where refills of medication are received, will vary according to the individual refill model (Page 56-66).

##### What happens at the clinical visit?

A **clinical visit** is a scheduled appointment where the clinician makes a thorough assessment and requests/reviews monitoring blood tests. According to the follow up protocol (Section 2.4.6), they may also be reviewed by the counsellor.

**In group models, the clinical visit should be scheduled together**. Each client is seen individually for their clinical review, but by coming together as a group, the clinician may assess group dynamics. A routine clinical visit should take 10-15 minutes with the clinician. For those clients with problems or treatment failure, more time should be allocated. A client should aim to spend no more that 2-3 hours within the facility from time of entry to time of departure.

The facility may ask clients attending for a clinical visit to pay a user fee. Refill visits should be free of charge.

At each ART clinical consultation, the following points should be addressed. Be guided by and complete the columns of the patient care and treatment booklet.

- Is the weight increasing or stable? Assess nutritional status.
  - If the weight is decreasing, WHY? Screen for possible TB; assess the nutritional status. Are there other signs of possible treatment failure?
- What family planning method is being used or is the client now pregnant and ANC/PMTCT interventions are needed? Are condoms also being used?
- Screen for TB. Is TB preventive therapy due?
- Screen for STIs.
- Take the patient's blood pressure.
- Are there any other complaints today?
- Are there any side effects of the medication being prescribed (swollen ankles or face, oliguria, polyuria,

haematuria for TDF; pallor, dizziness or breathlessness with AZT; sleep disturbance or altered mood with EFV; rash, yellow eye or right upper quadrant pain with NVP)?

- Check adherence to medications (not just the ART!).
- Are there any blood results viral load, creatinine, etc., that should be documented and reviewed today? If yes, have I acted on them?
  - Is the viral load >1000 copies/ml? If YES, the client needs to start enhanced adherence or should be being considered to switch regimens (Section 2.7).
  - If the client is on TDF and creatinine monitoring is available, is the last creatinine clearance more than 50ml/min? If no, discuss with nurse mentor or doctor regarding action.
- Are there any blood tests that should be ordered today? When routine viral load is available, it will be checked at month 6 and 12 and then yearly on ART. If on TDF, check creatinine yearly if available. If on AZT, check Hb after 6-8 weeks; thereafter, check HB if the patient presents with symptoms of anaemia.
- Prescribe medications (cotrimoxazole and ART) needed for today and complete documentation for subsequent ART refills. Whatever the refill option chosen, complete the patient care and treatment booklet and patient notebook, as shown in Appendix 5.
  - Column 2 now indicates the code for the type of ART refill model the client has selected.
  - For today's clinical visit, complete all columns.
  - To prescribe medication for subsequent one (if no VL monitoring) or three (where VL monitoring) refills, complete columns 11, 20a and 25.
- Complete the chronic ART register and appointment diary (this may be more efficiently done at the end of a session referring back to the patient care and treatment book) and/or send the patient care and treatment booklet for data entry into the EPMS.

##### What happens at the refill visit?

A **refill visit** is a scheduled appointment where the client has a pre-filled prescription and can collect the medication through a range of refill options (see Section 2.5.4). Whatever the choice of refill, the documentation will be carried out in the patient care and treatment book and patient notebook, as described below. This documentation will be carried out by the consulting health care worker or by whoever is dispensing ART from the pharmacy or distributing ART in the group- or community-based models. The specific ART refill models are described in detail in the standard operating procedures (SOPs) on pages 56-66.

- Fill or tick off columns 1, 2, 11, 20b, 26, 27, 28 to show that the patient has attended and the refill has been dispensed.

For group or community ART refill options, ART can be pre-packed (medication clearly named and labelled and ideally placed in individual patient-named bags) in order to allow for distribution by a primary counsellor, community health worker, expert client or client group representative.

**It is important to emphasise that if at any point the client has additional clinical needs, they can be seen by the clinician at any time and appropriate follow up organised.**

Clients must be educated on what symptoms and signs they should report in between appointments:

- Any symptoms/signs of TB: cough, fever, weight loss
- Diarrhoea or vomiting
- Ongoing or severe headache
- Persistent fever
- New rashes
- Symptoms or signs related to possible side effects of their medications.

#### 2.5.4 ART refill options for stable clients

According to the action plan for differentiating ART delivery carried out during the assessment phase, a range of options for ART refill may be considered to address the challenges faced by the facility and the patient. Although there should be flexibility, it is advised to focus on no more than two options in the first phase of implementation.

Note that the eligibility criteria and visit schedule is the same for all models (see Section 2.5.3).

**Before considering other refill options, ensure that the following three questions have been addressed:**

Is the maximum duration of ART for refills being prescribed? Refills should be given every three months. Before considering other models, ensure that the maximum refill duration is being prescribed in your site?

Are ART services being offered every day? If one day is heavily booked with ART clients, consider distributing over additional days.

Would extending opening hours of the pharmacy or ART clinic (early morning, late evening, Saturday morning) once a week or once a month improve retention for your clients, particularly working men and women and children at school?

Five refill options for stable clients on ART may be considered when planning how to differentiate ART delivery. Eligibility criteria are the same for all models (see Section 2.5.3). The choice of model should be guided by assessment of local data and consultation with health care workers and clients locally. Evidence published to date suggests that 20-50% of a facility's ART cohort may opt for a group model and, therefore, ensuring that an efficient individual facility-based option is offered should be a minimum requirement for all sites.

##### Fast-track

**Facility-based individual refill from pharmacy (see SOP, page 56) – target duration 30 minutes. This option should be available at all sites where drugs are dispensed from a separate room to where the clinical consultation is performed.**

The client collects their refill directly from the dispensing point. They do not queue to see the clinician. The client can collect the medication any time during clinic opening hours on his/her refill day. This model has most value in sites where dispensing is performed in a separate room by a different health care worker to the clinical consultation.

##### Club refill

**Facility-based health care worker-led group refill (see SOP page 58) – target duration 30-60 minutes. Experience to date suggests that this model is more popular in sites with large cohorts and in urban areas. As a group model, it provides the additional benefit of peer support.**

For clinics with larger cohorts, clients booked for refills on a given day can be organised into groups of 10-20. The group is then booked at the same time for each refill. On arrival, a health care worker (nurse, primary counsellor or expert client) facilitates discussion, identifies any group member who has a new clinical problem requiring review, and then distributes the medication. Medication can be pre-packed and labelled prior to the group meeting.

##### Outreach

**Community-based individual ART delivery through mobile outreach (see SOP on page 60). This option should be considered for clients from hard-to-reach areas or where existing outreach activities are already occurring to a fixed location. Logistics to support regular visits to the location must be ensured.**

Refills may be collected by the individual client and may be distributed by the nurse or, if pre-packed, by a primary counsellor, community health worker or expert client.

(If feasible differentiated ART delivery for different subpopulations and for clients with high viral loads may be delivered through outreach).

##### Community ART refill groups (CARGs)

**Community-based client-led group refill (see SOP on page 62). This refill option has been shown to be more popular in rural areas and where distance is a major challenge to the client. As a group model, there is the additional benefit of peer support. Some clients may benefit from the peer support gained by this model even where distance is not a challenge.**

Community ART refill groups are self-formed groups of clients on ART. They are usually from the same geographical area and are willing to disclose their HIV status to each other. The system ensures that all members attend the clinic for their clinical visits and monitoring blood tests together as a group. For refill appointments, the group members nominate one member to collect the drugs from the facility and distribute the refill to all group members.

##### Family member refill

See SOP on page 66. When a number of family members are on ART, it may be possible for one member to collect for the others. If a child is involved, it is essential that the child follows the paediatric follow-up schedule (Section 2.4.6) in order to ensure that drug doses are adjusted correctly according to weight.

#### 2.5.5 Additional considerations for differentiated ART refill options

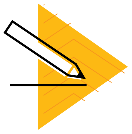

##### Promotion of refill options to clients

Once refill options have been chosen for a site, the details of the refill should be explained to the client during ART preparation and also included during the counselling at six months, prior to the first viral load, at which point the client may become eligible (see Job Aide, page 123). For clients already on ART for several years, clinicians should assess their stability according to the criteria outlined in Section 2.5.3 and explain the options available during the consultation.

Placing information material (posters to promote the variety of refill options) at the facility and utilising expert clients and community health workers to sensitise clients on the available models in the community may also support the uptake of the different options for differentiated ART delivery.

##### Incorporating family planning into follow up for stable clients

Women of childbearing age receiving ART through such refill models must continue to access family planning services. Utilisation of long-acting methods is advantageous as the

#### Table 12: Benefits of differentiated ART refill options

| | HEALTH SYSTEM | | | PATIENT | | | COMMENT |
|---|---|---|---|---|---|---|---|
| | Clinic decon-gestion | Additional documen-tation required | Additional resources needed | Decrease waiting time | Number of visits per year for each individual | Peer support | |
| Fast-track | X | | | X | 4 | | Should be available in all sites where dispensing is performed separately to the clinical consultation |
| Club refill | X | | | X | 4 | X | Often more appropriate in large volume urban sites |
| Outreach | X | | X | X | 4 | | |
| CARG | X | X CARG community refill form | | X | 1 (or 2 if nominated to collect) | X | Often more appropriate in rural sites or where distance to clinic a major barrier |
| Family member | X | | | X | Varied | | |

client does not need regular clinic visits. Where clients use depot injections, they should ideally be able to access this in the community via the community health workers or outreach activities. If this is not feasible, the client should attend the clinic independently for the regular depot injections.

##### Action when a client becomes "unstable" or enters a new sub-population

To enter into any of the refill options, the client must initially meet the definition of a "stable client" as defined in Section 2.5.3. However, at any point, the client may become "unstable" or move to another sub-population. Situations to consider include:

- Patient develops a high viral load >1000 copies/ml.
- Patient develops a concurrent OI, TB or other medical condition requiring additional medical follow up.
- Patient becomes pregnant.

If these situations occur, the client should be referred to the appropriate differentiated intervention, enhanced adherence counselling pathway, treatment of TB or integrated PMTCT/antenatal care. The client will then continue in this intervention until they again meet the eligibility criteria for stable clients. The client may continue to attend the group meeting if they wish to benefit from the ongoing peer support.

#### 2.5.6 Monitoring and evaluation for differentiated ART delivery for stable clients

In order to report the number of clients in each model of differentiated ART delivery, the coding of column two of the patient care and treatment book has been adapted to code the different refill options. These codes will then also appear in the automated appointment list:

- A Present self (conventional care)
- B Sent caregiver/treatment support/family refill
- C Visit made at another clinic
- D Community ART refill group (CARG)
- E Group facility pick up (club)
- F Individual pick up from facility pharmacy
- G Individual pick up via mobile outreach.

#### 2.5.7 Standard operating procedures for ART refills

Pages 56-66 outline the detailed standard operating procedures for the refill options.

Eligibility criteria, frequency of clinical review and reasons for requiring more intensive follow up are the same for all models.

Clients must be educated on what symptoms and signs they should report in between appointments:

- Any symptoms/signs of TB: cough, fever, weight loss
- Diarrhoea or vomiting
- Ongoing or severe headache
- Persistent fever
- New rashes
- Symptoms or signs related to possible side effects of their medications.

### STANDARD OPERATING PROCEDURE FOR "FAST TRACK"

#### Facility-based individual refill from pharmacy

This model should be available in all facilities where clients currently queue to see the clinician and again at the dispensing point.

**WHEN**

Every 3 months

Any time during opening hours

**WHERE**

Direct from dispensing point

**WHO**

The client does not see the nurse for a consultation, only the ART dispenser

**WHAT**

ART and CTX refill only

##### What preparation is needed before implementing this refill model?

- Training of health care workers on completion of documentation for the patient care and treatment book as per Appendix 5 and Section 2.5.3.
- Agreement with staff dispensing ART to complete the ART refill documentation.

##### Where is the refill given?

Direct from the pharmacy/dispensing point.

##### When is the refill given?

The client should be able to attend any time during clinic opening hours on their refill appointment date or at agreed times for fast-track refills for an individual facility. Extended opening hours for the pharmacy should be considered.

##### Who does the client see during the refill?

The client goes directly to the dispensing point and sees whichever staff member has been allocated to dispense medication. They do not queue to see the clinician.

##### What happens during the refill?

The client is asked at the dispensing point if they have any problems today. If they do, they are directed to the clinic for review. If there are no problems, the client receives their ART and cotrimoxazole refill. The refill documentation is carried out as documented in Appendix 5 by the dispenser of the medication.

The client should ideally receive their medication within 15 minutes, but should not wait longer than 30 minutes.

##### How are the patient care and treatment books filed?

As this is an individual model, the patient care and treatment books should be filed according to the standard cohort system.

#### What happens on a fast-track refill?

| | |
|---|---|
| **STEP 1** | • Ideally the day before use the EPMS appointment list or patient appointment diary to pull out the patient care and treatment books for the next day.<br>• Identify which clients are receiving ART in the fast-track model. |
| **STEP 2** | • For clients in the fast-track model, send the patient care and treatment books to the dispensing point. |
| **STEP 3** | • Client attends on day of refill appointment any time during clinic opening hours.<br>• Client attends the dispensing point directly.<br>• Client does not have individual nurse assessment unless patient requests. |
| **STEP 4** | • Dispenser provides refill as prescribed, and completes patient care and treatment book and patient notebook according to the refill documentation (Appendix 5). |
| **STEP 5** | • The patient care and treatment book is sent to the data clerks for entry into EPMS.<br>• At paper-based sites, the next refill date is written into the appointment diary.<br>• If any client does not collect medication as per their appointment, the standard defaulter tracing system should be triggered (page 51). |

### STANDARD OPERATING PROCEDURE FOR "CLUB REFILL"

#### Facility-based health care worker-led group refill

This model is often more appropriate in high-volume or urban sites and provides peer support.

**WHEN**

Every 3 months

At fixed meeting time for the group

**WHERE**

Refill takes place in room allocated for group refills

**WHO**

The group can be facilitated by a nurse, primary counsellor or expert client

**WHAT**

Group discussion and peer support

ART and CTX refill

If lay worker distributing drugs must be pre-packed

##### What preparation is needed before implementing this refill model?

- Training of health care workers on the model SOPs and completion of documentation for the patient care and treatment book as per Section 2.5.3
- If required, agreement with staff dispensing ART to pre-pack and label ART for the group refill session. Pre-packing of medication will facilitate groups being led by non- clinicians, such as the primary counsellors or expert clients
- A room or other location (e.g., waiting area) should be assigned, and defined booking times should be agreed on for the groups. These may also be in the afternoon, after working hours or at the weekends

##### How are the groups formed?

Groups can be between 10 and 20 clients. In order to facilitate group formation, a designated health care worker in the clinic (nurse or primary counsellor) should be allocated to coordinate group formation. Groups are formed primarily by the health care worker, and may be formed as the health care worker screens clients as eligible and refers them to the designated focal point for the groups. If there are pre-existing support group members or a sub-group of clients that would like to receive refills within the same group, then this should be facilitated. The list of group members with the contact details should be kept in the facility-held **facility group ART register (Appendix 6).** Each group should be given a specific group number, which is indicated on the front of the patient care and treatment book and in the patient-held notebook.

##### Where is the refill given?

The medication refill is given in the allocated facility room or location nearby the clinic (waiting area) where the group meets.

##### When is the refill given?

Each group is booked at a specific time to collect their refill. Ideally, the group should select the timing of their refill. Groups may be booked during or after clinic hours or at weekends.

##### Who does the client see during this refill?

The group should be facilitated, if possible, by the same HCW at each refill visit to establish rapport with the group. This may be a nurse or a lay cadre according to availability.

##### What happens during the refill?

Once group members arrive (a maximum of 15 minutes past the booked time for the group meeting should be given before the activities start), the HCW leading the group facilitates discussion. Clients are asked as a group if they have any specific clinical problems or any cough, sweats or weight loss. Any client with a clinical issue is then directed to see the clinician. Clients are then asked to share any other challenges or positive experiences they have faced with the group members. The length of the discussion is dependent on the participants, but the entire refill session should not take longer than 60 minutes. The HCW then distributes pre-packed and labelled medication to each group member individually.

##### How are the patient care and treatment books filed?

In EPMS sites, the patient care and treatment books should be filed in one folder labelled with the group number to facilitate pulling of files. The group number should be written on the front of the patient care and treatment book. At paper-based sites, files may be kept according to groups and replaced for cohort analysis or pulled individually at each group meeting.

##### What happens at the clinical review?

All the group members should be aligned to receive their clinical review at the same time, either once or twice a year according to Section 2.5.3. They are seen individually by the clinician, assessed clinically and have their viral load drawn. Aligning the clinical visit for the group facilitates uptake of viral load testing and allows the group to discuss VL results and other issues that are raised at the annual review together.

#### What happens at a club refill?

**STEP 1**

- The day before use the EPMS appointment list or patient appointment diary to identify which groups are attending the next day.
- Pull the patient care and treatment books for groups identified.

↓

**STEP 2**

- In settings where a lay cadre will distribute ART to the group (or where the team feels that pre-packing of medication will facilitate dispensing in the group room by the nurse), send the patient care and treatment books to the dispensing point for ART to be dispensed and pre-packed in patient-named bags.

↓

**STEP 3**

- At the time of the refill, the patient care and treatment books and dispensed pre-packed medication should be sent to the group meeting room.
- The clients in the group attend at the specified time for their group.
- If any clinical problem is identified, they are referred to see the nurse. This may be in the OI clinic or where the nurse facilitates the group, she may also consult directly if there is privacy.

↓

**STEP 4**

- Faciliated discussion is held for 30-45 minutes.
- The HCW distributes ART to the clients.
- The HCW distributing the medication should complete the patient care and treatment book as indicated in the refill SOPs in Appendix 5.

↓

**STEP 5**

- The patient care and treatment books are sent to the data clerks for entry into EPMS.
- The next refill date for the group, indicating the group number, is written into the appointment diary.
- If any client does not collect medication as per their appointment, the standard defaulter tracing system should be triggered (page 51). Group members themselves may be used to facilitate tracing.

### STANDARD OPERATING PROCEDURE FOR "OUTREACH"

#### Community-based individual ART delivery through mobile outreach

This model should be used if significant numbers of clients will benefit from provision of ART at a designated mobile outreach point in a hard-to-reach area. Commitment to continue visiting the site must be assured.

| WHEN | WHERE |
|---|---|
| Every 3 months<br>At fixed date and time | At fixed mobile outreach site |
| **WHO** | **WHAT** |
| The nurse<br>Primary counsellor or Expert client | ART and CTX refill only |

##### What preparation is needed before implementing this refill model?

- This refill option should be chosen only where it is guaranteed that the logistics for regular outreach to the point would be made available every three months.
- Health care workers should be trained on completion of documentation for the patient care and treatment book as per Section 2.5.3.
- A decision should be made on whether ART refill through mobile outreach will be integrated into existing outreach activities, or whether a stand-alone ART refill outreach is required due to numbers of people who may benefit in a specific hard-to-reach area.
- Prior to the day of outreach, prepare the HIV care and treatment booklets and medication required for refill. Formal pre-packing of medication may facilitate distribution at the outreach site.

##### Where is the refill given?

At a pre-defined community-based outreach location.

##### When is the refill given?

A fixed date and time is booked for the mobile outreach activity.

##### Who does the client see during this refill?

**Option 1:** The client sees the nurse who is performing the outreach activity.

**Option 2:** If the facility staff feels that pre-packing of medication would facilitate distribution by a primary counsellor or expert client supporting the mobile outreach, allowing the nurse to carry out other clinical duties clients, then the client will only see the HCW who distributes the pre-packed medication.

##### What happens during the refill visit?

The client is asked to attend the mobile outreach site. They are seen individually by the nurse or lay cadre, and ART is dispensed or distributed accordingly. If a clinical problem is identified, the client is reviewed by the clinician who is performing the outreach.

##### Where is the clinical review carried out?

If feasible and privacy can be ensured, it may be possible to perform the annual clinical review at the same outreach site. The nurse would review each client individually and take blood to perform viral load testing. If this is not possible, then the client should attend the facility for review once or twice a year according to the criteria in Section 2.5.3.

##### How are the patient care and treatment books filed?

As this is an individual model, the patient care and treatment books should be filed according to the standard cohort system.

#### What happens at a mobile outreach refill?

**STEP 1**

- The day before use the EPMS or patient appointment diary to pull the patient care and treatment books for clients booked for mobile outreach.

**STEP 2**

- Care and treatment books are used to prepare ART medication for outreach.
- If pre-packing, label medication with name of client and place all medication in a bag with client's name.

**STEP 3**

- Patient care and treatment books, ART and cotrimoxazole medication are transported to the mobile outreach site.
- Patients attend at the designated outreach site and are seen individually either by the nurse or lay cadre.

**STEP 4**

- ART may be dispensed by the nurse who completes the patient care and treatment book according to the refill SOP (Appendix 5).
- If pre-packed, ART may be distributed by the primary counsellor or other lay worker supporting the outreach activity. The distributor must complete the patient care and treatment book according to the refill SOP (Appendix 5).

**STEP 5**

- On return, the patient care and treatment book is sent to data clerks for entry into the EPMS.
- At paper-based sites, the next mobile refill date for each client is documented in the appointment diary.
- If any client does not collect medication as per their appointment, the standard defaulter tracing system should be triggered (page 51).

### STANDARD OPERATING PROCEDURE FOR CARGs

#### Community-based, client led group refill

This refill option is often more appropriate in rural or hard-to-reach settings. The model provides peer support.

| WHEN | WHERE |
| --- | --- |
| Every 3 months at agreed time in community and appointed date at facility | Community meeting is held at group members house or chosen community venue. Medication dispensed at facility or at a mobile outreach site and distributed in community |
| **WHO** | **WHAT** |
| Group Leader completes community form<br>Chosen group representative collects medication and distributes<br>Nurse sees group representative | ART and CTX refill only<br>Peer support |

##### What preparation is needed before implementing this refill model?

- Health care workers (nurses) should be trained on the model, completion of documentation for the patient care and treatment book as per Section 2.5.3, the community ART group register and community ART refill form.
- Health care workers should receive training to be able to provide the additional required education to the client group leaders (ensure basic treatment literacy and being able to complete the refill form correctly).
- To provide additional accountability within the community, sensitisation meetings on the model should be carried out with community leaders during the regular facility/ community meeting.
- Liaison with existing CBO groups and PLHIV support groups to facilitate group formation.
- Additional forms and tools required:
- Community ART group register (held at facility)
- Community ART group refill forms (given out from facility; held by the group)
- Folder to file completed community ART group refill forms for M&E purposes.

##### How are the groups formed?

Groups should be made up of 4-12 clients. In order to facilitate group formation, a designated health care worker in the clinic (nurse or primary counsellor) should be allocated to support group formation. Groups are ideally self-formed from clients known to each and who are from the same geographic location. In the initial phases of scale up, groups may often be formed from existing support group members or through groups known for existing CBO activities. When this model is being established, the clinic focal person may also play a role in identifying clients on ART from the same geographical area who, with their consent, are introduced to each other.

After the initial phase of group formation, new clients who have been classified as stable and who are from an area where there is an existing community ART group known to the clinic focal point may also, with their permission, be introduced to the group.

Such self-formed groups may then present themselves at the facility and be assessed by the clinician to ensure that they meet the eligibility criteria. In addition to being clinically stable, all group members must meet a minimal treatment literacy requirement: having a basic knowledge of HIV and being able to name and identify their medication. If there are gaps, the basic ART education sessions illustrated on pages 110-118 of the Job Aide can be reiterated.

Once formed, each group should be assigned a group number. The list of group members with their contact details should be kept in the facility-held **community ART group register (Appendix 6).** The group number should be indicated on the front of the patient care and treatment book and in the patient-held notebook.

#### Selection and training of the group leader

Each group must select a group leader who receives additional education on how to conduct the community ART refill meeting and complete the community group refill form. The group leader must have basic reading/writing and treatment literacy skills, but does not have to be seen as an expert client.

Training of the group leaders should be the responsibility of the facility staff, nurse or community ART group focal point. If there are a number of new groups, the group leaders may be brought together at the facility. It is often more effective to complete the education with the group leader in collaboration with their group members. This ensures that all group members understand the system, but it must be ensured that the group leader holds responsibility for ensuring the refill form is completed correctly.

Topics to be covered:

- Recap on basic treatment literacy topics, HIV/AIDS, use of ART and viral load monitoring. Pages 110-118 of the Job Aide may be used to recap these topics.
- The schedule of the refill visit, as outlined in this SOP. The Job Aide (Page 71) should be used during this education session.
- How to complete the clinical questions on the community group refill form outlined in the section, "to be completed by group leader".

#### Where is the refill given?

The group meets at an agreed meeting point in the community usually the day before their refill appointment (most often one of their houses). The group nominates one member to collect the refill medication for the group from the facility. If there is a member with a clinical problem at that time, that client will be nominated to attend. If very unwell, that member would be supported and accompanied by another member in case investigation or admission is required.

#### When is the refill given?

The group is given an appointment date and the group representative attends during working hours in the same way as a client in conventional care. The group member should be given a place in the clinic queue according to when they arrived in the same way as other clients and does not have to be prioritised.

#### Who does the client see during this refill?

The group representative is seen by the nurse.

#### What happens during this refill?

The group refill form is reviewed by the nurse who addresses any questions and concerns with the group representative. The nurse then completes their section of the group refill form, indicating prescription of the ART, and completes the patients care and treatment booklets according to the refill SOP (Appendix 5).

ART is then dispensed for each group member in a named pre-packed bag.

The group representative then returns to the community to distribute the ART to their group members. Each member must sign on the group refill form that they have received their ART. This form is then also returned to the clinic nurse at the subsequent refill.

#### How are the patient care and treatment books filed?

In EPMS sites, the patient care and treatment books should be filed in one folder labelled with the group number. The group number should be written on the front of each book and in the patient-held notebook. This facilitates pulling of files on days when group refills are booked. At paper-based sites, the patient care and treatment books may be filed according to their CARG group and replaced only when cohort analysis is required.

#### What happens at a CARG refill?

**STEP 1**

- The day before their refill date or early morning on the refill date, the community group members meet at chosen house/community venue.
- The group leader completes the CARG refill form together with the group members.
- The group chooses a representative to attend the clinic to collect the ART; if a member has a clinical problem, this member is selected. The group representative takes the completed community ART refill form from the previous visit and the one completed for this refill to the clinic.

↓

**STEP 2**

- At the facility, use the EPMS or patient appointment diary to pull the patient care and treatment books for clients booked for CARG refill; care and treament booklets should be filed according to their group membership.
- The CARG representative is seen by the clinic nurse.

↓

**STEP 3**

- The nurse reviews signatures from the previous refill form to ensure all clients have received their medication (this form is filed in a "CARG refill folder").
- Prescription of ART is given and documentation of any results is made on today's refill form.
- Today's community group refill form is given back to the CARG representative.
- Patient care and treatment book is filled according to the refill SOP (Appendix 5).

↓

**STEP 4**

- Patient care and treatment book is sent to data clerks for entry into EPMS.
- Next refill date for the group is documented in the appointment book (can document group number rather than individual names).
- If any group representative does not collect medication as per their appointment, the standard defaulter tracing system should be triggered (page 51).

↓

**STEP 5**

- Group representative returns to the community and distributes the ART to their group members.
- Each member signs that they have received their refill.

##### How is the clinical review carried out?

All group members should have their individual clinical visit aligned so they are seen together for their clinical review and laboratory monitoring once or twice a year according to whether CD4 or VL is available. Bringing all the group members together enhances the uptake of viral load, allows group discussion of results, and allows the facility staff to see the group together and address any problems.

##### The role of the community health worker

CARGs should function independently. However, with the clients' permission, the community health worker (CHW) in their area should be made aware of any group in their catchment area. The community health worker may then act as a point of contact should problems arise within the group and, if necessary, be a means of communicating from the facility to the group. The CHW does not have to attend the group meeting and should not be seen as having a supervisory role.

### CARG Community Refill Form

| Community ART group refill monitoring form | | | | | | | | | | | | | | |
|---|---|---|---|---|---|---|---|---|---|---|---|---|---|---|
| Facility name: | | | CARG number: | | | | | | | | | | | |
| Date group meeting before refill ../ .. /.... | | | Signature of group leader: | | | | | | | | | | | |
| Date ART prescribed by nurse ../ .. /.... | | | Signature of nurse: | | | | | | | | | | | |
| Date ARVs distributed ../ .. /.... | | | | | | | | | | | | | | |
| | To be completed By Group Leader | | | | | | To be completed by nurse | | | | To be completed By CAG member | | | |
| CARG member number | Full name | Pregnant (P) or on family planning (FP) | TB symp-toms* Y/N | Other "alert" prob-lems** | ARV tablets remain-ing | CTX tablets remain-ing | ARV regimen prescribed / quantity | CTX quantity prescribed | VL result (CD4 if not available) | Date VL | Full name | Signa-ture of recipi-ent | Date drugs received | Comments (include any reason for tempo-rary clinic follow up) |
| 1 | | | | | | | | | | ../ .. /.... | | | ../ .. /.... | |
| 2 | | | | | | | | | | ../ .. /.... | | | ../ .. /.... | |
| 3 | | | | | | | | | | ../ .. /.... | | | ../ .. /.... | |
| 4 | | | | | | | | | | ../ .. /.... | | | ../ .. /.... | |
| 5 | | | | | | | | | | ../ .. /.... | | | ../ .. /.... | |
| 6 | | | | | | | | | | ../ .. /.... | | | ../ .. /.... | |
| 7 | | | | | | | | | | ../ .. /.... | | | ../ .. /.... | |
| 8 | | | | | | | | | | ../ .. /.... | | | ../ .. /.... | |
| 9 | | | | | | | | | | ../ .. /.... | | | ../ .. /.... | |
| 10 | | | | | | | | | | ../ .. /.... | | | ../ .. /.... | |
| 11 | | | | | | | | | | ../ .. /.... | | | ../ .. /.... | |
| 12 | | | | | | | | | | ../ .. /.... | | | ../ .. /.... | |
| *TB symptoms: Ask if the member has a current cough of any duration, is losing weight, has night sweats or has had contact with TB patient in last month | | | | | | | | | | | | | | |
| **Alert problems: Ask if the member has any ankle swelling, puffiness of the face, breathlessness, diarrhea for more than 2 weeks, severe headache | | | | | | | | | | | | | | |

**Step 1:**

In the community meeting the group leader completes this section.

They ask all group members about TB symptoms and other clinical or adherence problems.

**Step 2:**

The group representative attends the clinic bringing the refill form from previous visit and from this refill.

The nurse checks the previous form to ensure all clients have received their medication.

The nurse completes this section and completes the patient care and treatment booklet according to the refill SOP (Appendix 5).

**Step 3:**

The group representative distributes the medication and shares any results. Each group member must sign that they have received their medication.

### STANDARD OPERATING PROCEDURE FOR "FAMILY MEMBER" REFILL

This model follows the same steps as the community client-led group refills, but the group is defined by being made up of family members.

##### How are the patient care and treatment books filed?

The files should be kept together as a group.

##### How is the clinical review carried out?

The clinical review of the family should be done together.

**WHEN**

Every 3 months

**WHERE**

Family group completes group refill form at home

Medication is collected from the facility or mobile outreach point

Medication is then distributed at home

**WHO**

Nurse sees family representative at facility or mobile outreach

**WHAT**

ART and CTX refill only

#### What happens at the time of refill?

**STEP 1**

- The day before use the EPMS or patient appointment diary to pull the patient care and treatment books for clients booked for refill via this model.

**STEP 2**

- Family representative completes the group refill form at home.
- Family representative attends facility or mobile outreach site, bringing today's refill form and the completed refill form from the previous visit.

**STEP 3**

- Family representative is reviewed by nurse.
- ART for all family members is dispensed.
- Dispenser completes patient care and treatment book according to the refill SOPs (Appendix 5).

**STEP 4**

- Patient care and treatment book is sent to data clerks for entry into EPMS.
- The next appointment for the family group is documented in the appointment diary.
- If any client does not collect medication as per their appointment, the standard defaulter tracing system should be triggered (page 51).

**STEP 5**

- Family member distributes ART to other family members at home.
- Each member signs that they have received the medication.

### 2.6 Differentiated ART delivery for specific sub-populations

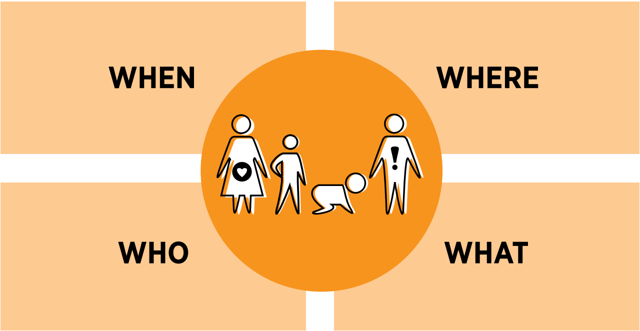

Using the building blocks of differentiated service delivery, the following section outlines ways that ART delivery may be differentiated according to the sub-population.

#### 2.6.1 Differentiated ART delivery for pregnant and breastfeeding women

##### When are ART services delivered?

PMTCT services should be available during standard clinic hours, and must also be available 24 hours a day in the maternity unit to ensure that services are provided to women during labour and delivery.

##### Where are ART services delivered?

ART delivery should be integrated into existing MNCH services antenatally, at delivery and postnatally. Women should receive their SRH and ART services on the same day, in the same room, from the same health care provider as outlined in Section 1.4.3. Postnatally, mothers, HIV-exposed and HIV-positive infants and, where appropriate, their fathers should receive their care together on the same day in the MNCH department until the exposed child is discharged or an HIV-positive child reaches five years of age.

Where antenatal services are provided in regular mobile outreach, provision of ART refills for HIV-positive clients should also be considered.

##### Who provides ART delivery?

In the integrated model, nurse midwives should be trained to provide SRH and ART services to clients. In addition, evidence supports the role of expert clients who have experience of PMTCT to support adherence counselling and defaulter tracing. Such counsellors may be either facility or community based.

##### Refill options for pregnant and breastfeeding women

HIV-positive pregnant women require additional antenatal and postnatal services, including specific follow up of the exposed infant. The client may therefore be required to be seen more frequently for these additional medical services. Both antenatal and postnatal clients should be offered the option of "group ART refills" using the same standard operating procedures as for facility-based health care worker-led groups. Such groups should be facilitated by the health care worker or expert client.

For women already on ART who become pregnant and who are already in a group refill option they should be offered the choice of continuing to access their ART through the group or through MNCH. However it must be ensured that the client attends for the additional antenatal and postnatal clinical and counseling services.

#### 2.6.2 Differentiated ART delivery for men

Ensuring that services are provided in an environment that is conducive to men's needs and at times that are male friendly should be reviewed. Consultation with community leaders could facilitate ideas about how services can be made more attractive to men. Possible interventions that may improve access to care for men are:

- A weekly or fortnightly extended-hours service that men can access (NB: These extended hours services should also be available to other clients as well).
- Adopt refill policies to reduce time spent at the clinic.
- Offer other "male" services (BP, diabetes, prostate screening and health education advice on smoking and alcohol use) alongside provision of HIV prevention, care and treatment services.
- Offer flexible refills for men working away from home.

#### 2.6.3 Differentiated ART delivery for children and adolescents

##### When is ART given?

**Children aged 0-2 years** should have a clinical visit monthly. Children may be booked on the same day for clinical review for additional peer support for the parents but are seen individually for clinical review.

**Children 2-5 years** should have a clinical visit every three months. Children may be booked on the same day for clinical review for additional peer support for the parents but are seen individually for clinical review.

**Children >5 years** until on adult doses should have a clinical visit every three months.

**Adolescents** who are on adult dose and who are fully disclosed should have a clinical visit every six months but may be offered the refill options described in Section 2.5.4 once they meet the eligibility criteria for a stable client.

For school-age children and adolescents, appointments should be booked outside school hours whenever possible. Children attending boarding schools should be given longer refills to match their term times and appointments during the school holidays. Strategies for how they will manage their medication at school must be discussed with the child, guardian and, ideally, teaching staff.

##### Where is ART given?

**Children aged 0-2 years** should be seen in MNCH through a **family approach** – mother and child being booked on the same day.

**Children 2-5 years** should be seen in MNCH through a **family approach** – mother and child being booked on the same day.

Older children (>5 years) and their guardians should be seen in the OI clinic and be offered a **group refill approach**.

##### Who provides ART?

Health care workers should receive training in order to provide adequate follow up for children and adolescents. Every clinic should have at least one health care worker trained to perform paediatric disclosure counselling. Providing care to HIV-positive adolescents requires specific skills. A training on the care of HIV-positive adolescents is available from the MoHCC. One of the most important factors is for health care workers to approach adolescent and young adult clients in a non-judgmental manner.

Adolescent or young adult peer expert clients have been shown to provide additional support for adherence and retention. These peers should receive some basic training and may support treatment literacy activities and tracing of those clients who default. They may be based at facility level to help facilitate clinic activities on child and adolescent clinic days, and also engage with clients in the community to provide additional adherence support.

##### What additional services should be provided for children and adolescents

Special considerations to be taken into account for the clinical follow up of a child on ART are listed below.

**Monitoring of weight and height is VERY important for assessing a child's response to ART and to ensure the correct medication dose is given.** Weight and height should be plotted on the growth chart so trends can be followed. If the growth curve is flattening or dropping down the centiles, we need to investigate why?

1. In the history and examination:
   - Screen the child for TB? Poor growth is a possible sign of TB in children.
   - What is the nutritional status of the child?
   - Are there other signs of treatment failure (for definitions, see Section 2.7.1): new rashes, other OIs, recurrent cough, diarrhoea?
   - Are there other intercurrent illnesses?
2. Assess developmental milestones (refer to clinical guidelines).
3. Are the child's immunisations up to date?
4. The clinician should be aware of the child's disclosure status and work together with the counsellor to ensure that full disclosure is reached by the age of 12.
5. The doses of **ARVS must be prescribed according to the weight of the child. DO NOT JUST COPY** what your colleague wrote the last time. **If the dose is not increased, this may lead to sub-optimal dosing and possible development of resistance and treatment failure.**

In addition to clinical review and ART refills, an essential component of follow up for children is to ensure HIV disclosure counselling has been carried out. Tools to assist HIV disclosure can be found on pages 125-137 of the Job Aide.

Adolescents should also be offered access to SRH education and family planning services in a non-judgmental environment.

##### Refill choices for stable children and adolescents

*Eligibility*

Until on adult doses children must have regular clinical review ( monthly age 0–2 years ; 3 monthly if 2 years and above). Bringing younger children together on the same day will allow parents to share experiences and challenges and allow the children to play together. Having the children on a particular day will also help the health care worker focus on the needs of the children and be a reminder for the nurse and counsellor to have their child-friendly set of tools with them.

Once on adult doses, VL < 1000 copies/ml and no concurrent OIs, children and adolescents who are fully disclosed may be seen every six months with one refill in between.

The child and their carer should be given the choice of a fast track, family member or club refill. For adolescent clubs, the role of the peer community adolescent treatment supporter – both to facilitate the group and to give additional support in the community – has been shown to improve retention and adherence. Identified adolescent peers should receive additional training and support to perform this role.

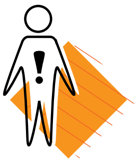

#### 2.6.4 Differentiated ART delivery for key and vulnerable populations

##### When are services delivered?

If stand-alone services are to be offered, the timing of clinics should be made after consultation with the specific key population. For example, in many settings, female sex workers prefer to access services in the mid-afternoon to early evening.

##### Where is ART delivered?

ART delivery may be integrated into routine clinic services or where demand and resources permit a stand-alone service may be provided.

##### Who delivers ART?

Health care workers should offer services in a non-judgmental way to all key populations. Where feasible, they should receive specific training in order to offer the adapted service package for the specific sub-population.

Peer educators have been shown to impact on adherence and retention and should be considered when designing services specifically for these groups. They may play specific roles in treatment literacy and defaulter tracing.

##### What services are delivered for key populations?

In addition to the minimum package of services, specific key populations will require additional services. Examples of these include:

All:

- STI screening
- Sexual and gender-based violence services
- Condom and lubricant provision
- Hepatitis B vaccination.

People who inject drugs:

- Clean needle provision
- Methadone replacement therapy.

##### Refill options for key populations

Clients from key populations who are stable on ART should be offered refill options as outlined in Section 2.5.4. Group refill options with the group formed from the same key population (facility-based HCW-led groups or client-led community group refills) may also provide additional peer support, but should be entered into voluntarily. Facility-based group refill approaches may be facilitated by peer expert clients where appropriate.

#### 2.6.5 ART delivery in prisons

Clients living with HIV in prisons should have access to the minimum package of ART delivery services. Due to issues around storage of medication within the cells, a one-month supply of ART should be issued, with clients being seen for a clinical review every three months.

Additional services that should be offered include:

- Hepatitis B vaccination
- Condoms
- Clean needles
- TB screening.

#### 2.6.6 Differentiated ART delivery for people living with disabilities

ART services for people with disabilities (impaired hearing, vision, mobility) should be adapted wherever feasible. People living with HIV, especially children who were vertically infected, have very high rates (up to 52%) of disability. Those at risk should be clinically assessed by their health care workers and be referred where possible to rehabilitative services and access to assisted devices.

Ensuring adequate treatment literacy is achieved in the initial stages of ART use is especially important, and efforts should be made to access information in accessible formats. Each district should have access to the list of services that provide support (braille communication, sign language services, etc.) in their locality, and link clients with such services. Trained signers in a given locality should also be supported to be able to give HIV and ART basic education.

##### 2.6.7 Differentiated ART delivery for mobile populations

All clients should be asked about their travel plans during ART consultations in order to plan their ART refill duration.

However, some clients will be identified as having particular challenges related to mobility due to their work or family circumstances. For clients who are working abroad or travelling frequently, the following steps should be discussed in the consultation:

- Duration of ART refill may be extended to up to 6 months.
- A designated family member should be permitted to collect medication for the client.
- For those clients living in neighbouring countries, linking the client to a cross-border clinic should be investigated.
- It is advisable that **a transfer letter is completed and kept updated with the client's latest regimen and monitoring results.**
- If the client is travelling to a fixed destination, identify an ART site where they can access care and ART delivery in an emergency.

### STANDARD OPERATING PROCEDURE FOR FACILITY-BASED ADOLESCENT GROUP REFILL

**WHEN**
Every 3 months

**WHERE**
At the facility

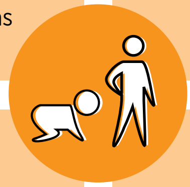

**WHO**
Nurse assisted by primary counsellor or community adolescent treatment supporter

**WHAT**
ART and cotrimoxazole refills

Peer support

SRH education and services for adolescents

##### What preparation is needed before implementing this refill model?

- Health care workers should be trained on specific facilitation methodologies for adolescents.
- If feasible, 1-2 trained peers should be identified. These peers will co-facilitate the group sessions and provide additional support as required in the community, including tracing of defaulters.
- A room should be assigned and defined booking times for when the groups will meet agreed on. These can also be in the afternoon, after school hours or at the weekends, but should be agreed on with the group members.
- Pre-packing of medication will facilitate groups being led by non-clinicians, such as the primary counsellors or adolescent and young adult peer facilitators.
- These groups will function as a support group while also providing ART refills.

##### How are the groups formed?

Groups can be made up of between 10 and 20 clients (and the caregivers of younger children). In order to facilitate group formation, a designated health care worker in the clinic (nurse or primary counsellor) should be allocated to coordinate group formation. Groups are formed primarily by the health care worker, and may be formed as clients are screened as eligible by the health care worker, consent has been provided by the caregiver, and referral made to the designated focal point for the groups. Groups may be formed from existing support groups.

It is suggested that children and adolescents are grouped according to their disclosure status and with similar age groups (10-14, 15-19, 20-24). The list of group members with the contact details should be kept in the facility-held **facility group ART register (Appendix 6)**. Each group should be given a specific group number, which is indicated on the front of the patient care and treatment book and in the patient-held notebook.

##### Where is the refill given?

The medication refill is given in the allocated facility room where the group meets. Ideally medication should be pre-packed and labelled to facilitate fast distribution and allow a lay cadre to run the group.

##### When is the refill given?

Each group is booked at a specific time to collect their refill. Ideally, the group should select the timing of their refill. Groups may be booked during or after clinic hours or at weekends.

##### Who does the client see during this refill?

The group should be facilitated, ideally, by the same HCW at each refill visit. This HCW should have received some training on facilitation of groups and adherence counselling for adolescents. This may be a nurse or a lay cadre supported by a peer facilitator.

##### What happens during the refill?

Once group members have arrived (a maximum of 15 minutes past the booked time for the group meeting should be given before the activities start), the HCW leading the group facilitates discussion. Clients are asked as a group if they have any specific clinical problem or any cough, sweats or weight loss. Any client with a clinical issue is then directed to see the clinician. There should be a specific activity for the day chosen from a selection of topics that can be rotated at each refill session. The choice of topics should recognise the age, developmental and disclosure status of each group. Topics may include:

- Growing up: changing bodies, changing emotions, feeling good about ourselves.
- Coping with difficult situations, problem solving.

- Sex and relationships: love and sex, safer sex, social pressures to have sex.
- Unwanted pregnancy and use of contraception; PMTCT.
- Living with HIV.
- ART and adherence.
- Disclosure.
- Relationships with family and friends.

There should also be time allocated for some fun, singing and games.

Further information and tools that may facilitate these sessions may be found at:

- http://www.tarsc.org/auntiestella/index.php/
- www.africaid-zvandiri.org

The length of the discussion is dependent on the participants, but the entire refill session should not take longer than 90 minutes.

The HCW then distributes pre-packed and labelled medication to each group member individually.

##### How are the patient care and treatment books filed?

The books should be filed in one folder labelled with the group number. The group number should be written on the front of the patient care and treatment book. This facilitates pulling of files on days when group refill options are booked.

##### What happens at the clinical review?

All the group members should be aligned to receive their clinical review at the same time. For adolescents, this should be twice a year. At this visit, they are seen individually by the clinician, assessed clinically and once a year have VL drawn. Aligning the clinical visit for the group facilitates uptake of viral load testing and allows the group to discuss VL results and other issues that are raised at the annual review together.

#### What happens at an adolescent group refill

**STEP 1**

- The day before use the EPMS appointment list or patient appointment diary to identify which groups are attending the next day.
- Pull the patient care and treatment books for groups identified.

**STEP 2**

- In settings where a lay cadre will distribute ART to the group (or where the team feels that pre-packing of medication will facilitate dispensing in the group room by a nurse facilitating the groups), send the patient care and treatment books to the dispensing point for ART to be dispensed and pre-packed in patient-named bags.

**STEP 3**

- At the time of the refill, the patient care and treatment books and pre-packed medication should be sent to the group meeting room.
- The clients in the group attend at the specified time for their group.
- If any clinical problem is identified, they are referred to see the nurse. This may be in the OI clinic, or the nurse may also consult when she facilitates the group.

**STEP 4**

- Faciliated discussion is held for 30-40 minutes and fun games and activities for another 30-40 minutes.
- The HCW distributes ART to the clients.
- The HCW distributing the medication should complete the patient care and treatment books according to the refill SOP (Appendix 5).

**STEP 5**

- The patient care and treatment books are sent to the data clerks for entry into EPMS.
- The next refill date is written into the appointment diary indicating the group number
- If any client does not collect medication as per their appointment, the standard defaulter tracing system should be triggered (Page 51).

### 2.7 Differentiated ART delivery for clients with high viral load

#### 2.7.1 Detecting treatment failure

Clients on ART should be monitored to ensure that their treatment is successful. Monitoring for treatment failure can be defined in three ways: clinical, immunological, and virological.

Routine viral load is the monitoring strategy of choice and is being phased in across Zimbabwe. Viral load has been chosen as the monitoring strategy of choice because virological failure is the first type of failure to occur.

Performing a viral load test is not enough. **The result must be acted on.** In addition to clients with a high viral load, clients who have attended late for their appointments or appear to be having significant psychosocial problems should receive additional enhanced adherence counselling.

##### Clinical failure

The definition of clinical failure is a new Stage 4 OI in adults or Stage 3 or 4 in children. However, clinicians should be alert to new Stage 2 or 3 OIs, recurrent episodes of diarrhoea, respiratory tract infections, new rashes and decreasing weight (it is important to document sequential weights, especially in children, in the patient care and treatment book).

**A targeted viral load should be requested where there is no access to routine viral load; new Stage 2, 3 or 4 OIs; a 10% drop in weight; or the client is recurrently unwell.** If viral load is not available and the client is very sick, refer immediately to a doctor who will assess the client.

A client on ART presentin**g with new OIs needs URGENT assessment for possible treatment failure management. A targeted viral load should be taken urgently.**

##### Immunological failure

If viral load monitoring is not available, immunological criteria should continue to be used. The immunological criteria for failure are: in adults, a CD4 count at or below 250 cells/mm$^3$ following clinical failure or persistent CD4 levels below 100 cells/mm$^3$; in children younger than five years, a persistent CD4 count below 200 cells/mm$^3$; and in children older than five years, persistent CD4 levels below 100 cells/mm$^3$ after six months of effective treatment. If these criteria are met, a targeted viral load should be requested.

##### Virological failure

Routine viral load (month 6 and 12 and then yearly) is being phased in across Zimbabwe. During this period, all clinics should still have access to targeted viral load at their district or provincial referral hospital for clients who have clinical or immunological failure. Blood should be drawn where the dried blood spots will be prepared and then transported to the viral load testing laboratory. Where plasma samples are being used, samples must reach the laboratory where centrifugation is performed within six hours of the blood being drawn.

**A viral load >1000 copies/ml NEEDS ACTION. If there are signs of clinical failure, the client must be reviewed urgently by the doctor to decide whether a switch is required immediately.**

Figure 8 outlines the algorithm for when to take a viral load and the action to be taken depending on the result.

#### Figure 8: Routine viral load monitoring algorithm

**TAKE VIRAL LOAD TEST :**

- **Any patient with clinical or immunological failure must be urgently assessed individually**
- Routinely at 6 months after starting ART, 12 months after starting ART and then every 12 months (24, 36 months etc.)
- Give viral load key messages before client is bled for VL

**VL <1000 COPIES/ML**

- Maintain first-line therapy
- Schedule next VL testing at month 12 after ART initiation then yearly thereafter
- Offer client options for differentiated ART delivery for stable clients

**VL >1000 COPIES/ML**

Assess for and address any possible causes of non-adherence and treatment failure

Refer for enhanced adherence counselling (EAC)

↓

1st EAC session on day of result

↓

- 2nd EAC session after 4 weeks
- 3rd EAC and additional sessions as required over the next 8 weeks

↓

Repeat VL 12 weeks after result has been given

**VL <1000 COPIES/ML**

- Maintain current regimen
- Offer client options for differentiated ART delivery for stable clients
- Schedule next VL testing at month 12 and yearly

**VL >1000 COPIES/ML**

- Refer to clinician experienced in switching to second line
- Gather information on patient from both clinicians and counsellors
- Switch to second line within 2 weeks from receipt of second high viral load unless clear clinical or psychosocial contraindication
- Urgency of switch will be dependent on clinical condition of patient, CD4 or if woman is pregnant or breastfeeding

**This algorithm should be completed within 6 months**

**Use the EPMS appointment system to identify client:**

**Who is due VL testing**

**Whose last viral load was >1000 copies/ml. These clients need:**

**Enhanced adherence counselling**

**OR**

**Repeat VL testing**

**OR**

**Change of regimen**

**If there is evidence of clinical or immunological failure the patient should be referred to a clinician and a treatment plan made on an individual basis. In these cases a switch to second line may be considered earlier and without repeat of VL in some cases.**

If point of care viral load is available, use this for patients who are clinically unwell and for the repeat VL test after enhanced adherence.

#### 2.7.2 Scaling up routine viral load

For effective scale up of routine viral load monitoring, the following programmatic elements must be considered:

- Client education to raise awareness of viral load, create demand for testing and ensure understanding of results
- Health care worker education on how to prepare the DBS sample, complete the VL request form and complete the actions in the viral load algorithm
- Primary counsellor or nurse education on the delivery of enhanced adherence counselling
- Programmatic clinic systems to ensure uptake of viral load and action on results.

##### Client education on treatment failure and viral load monitoring

As part of routine adherence counselling from ART preparation onwards, clients should be educated about signs and symptoms of possible treatment failure (weight loss, recurrent infections, new rashes, any new staging condition). This is important so that they can report promptly to their health facility. Having community health workers and other CBO members educated about treatment failure will also provide additional support to ensure that clients with possible treatment failure are encouraged to attend the facility before a booked appointment.

As viral load is rolled out, client education on viral load is an essential step in the implementation plan. Clients should know when their viral load should be taken, why it is being taken, and how to interpret the result. All clients should know that if the viral load is >1000 copies/ml, there is possibly an adherence problem or resistance, and action should be taken by the clinician and the counsellor.

Key messages on viral load (Job Aide, page 121) should be given to clients during ART preparation and prior to bleeding for VL. This session can be as a group if patients are triaged for viral load blood draw together. An undetectable viral load does not mean that the virus is no longer there. Make sure that the patient understands this, and that they know that they must continue to take their ARVs if their viral load is suppressed.

##### Identification of clients due viral load testing

Clients who are due viral load testing must be identified on arrival. Strategies that may enhance identification include:

- Use the EPMS appointment list that can identify how many months ago the last VL was taken and if VL is now due.
- Documentation in the client's booklet of the services required at the next appointment, e.g., clinical review and VL. This will assist the health care worker performing triage to quickly identify those in need of blood draw.
- If clients are in differentiated ART delivery models for stable clients, they will only have a clinical review once a year. This review is therefore always accompanied by a viral load test.
- Clients who are in group models should have their clinical review and viral load testing aligned. This will improve uptake and facilitate peer support when the results are received.

Standard operating procedures for making a DBS sample can be found on page 75 of the Job Aide.

#### 2.7.3 Action on receipt of the first viral load result

When the VL results are received:

- The viral load result should be **entered into the EPMS.**
- The file should be pulled, the **the VL result documented in the VL column of the patient care and treatment book and the result filed**.
- For clients with VL >1000 copies/ml, the file should be flagged or placed in a "high viral load tray". Sticker systems may be used if available (red sticker placed on file if >1000 copies/ml; and green sticker placed on file if <1000 copies/ml). Note: Red stickers should be removed when the patient suppresses to <1000 copies/ml.
- The high viral load form should be opened. As the steps of the VL algorithm are worked through:
- The nurse should complete the patient information, previous VL results and ARV information.
- The primary counsellor or nurse performing EAC should complete the enhanced adherence counselling section at each session. Additional notes can be made in the notes section of the patient care and treatment book.
- The nurse should fill the outcome section with the clinical screening information.
- The clinician trained to switch completes the final outcome section if the second VL is persistently high.

#### Figure 9: Action on receipt of viral load results

**RESULTS ARRIVE**

- All VL results are entered into the EPMS.
- All VL results are documented in viral load result column of the patient care and treatment booklets and filed.
- For all VL results >1000 copies/ml, files should be kept out and flagged either by placing in high VL tray or with a red sticker (these must be removed once the patient has stabilised).
- Open the high viral load summary form.

↓

**TRACE PATIENTS WITH HIGH VIRAL LOAD**

- Staff member is delegated to trace patients with high viral load via phone or through the community health worker.
- SMS of high viral load results, sent to to both clinic and patients, has been introduced as an additional means of contacting patients.

↓

**PATIENT ATTENDS FOR FIRST EAC SESSION**

- Patient is identified through the flagging system (red sticker on file or EPMS appointment flag) and is triaged to EAC on arrival.
- Patient is given 1-month refill and booked for a second EAC session in one month. If significant psychosocial issues are identified during the first session, the client can be seen sooner if required.

- Unless the client is attending within the next two weeks, the client **must be traced** and asked to come to the clinic as soon as possible. The client may be phoned or traced via a community health worker using the AIDS and TB referral form.
- Clients who have consented to receiving an SMS with their results will also receive an SMS advising them to attend the clinic as soon as possible.
- If the client is not traced and only attends at their next appointment, the EPMS appointment list or red sticker should flag them as a client with high VL at their next booked appointment.

##### Action plan for a client with a first viral load less than 1000 copies/ml

When the patient is suppressed, the client should be congratulated and encouraged to continue with their current regimen. If they meet the additional eligibility criteria, they should be offered the options for differentiated ART delivery for stable clients that are available at their site (see Section 2.5.4).

##### Action plan for a client with a first viral load more than 1000 copies/ml

No client should be switched on the basis of one viral load except where there are clinical reasons for an urgent switch of regimen. Any patient with signs of clinical failure should be urgently assessed by an experienced clinician. For clients who are clinically stable the repeat viral load should be repeated 12 weeks after the first result has been given.

Table 13 outlines the common reasons for a high viral load. Poor adherence is the most common reason and is the focus to explore during enhanced adherence sessions.

#### Table 13: Common reasons for a high viral load.

| PSYCHO-SOCIAL REASONS FOR HIGH VIRAL LOAD | CLINICAL REASONS FOR HIGH VIRAL LOAD |
|---|---|
| Poor adherence may be:<br>• Behavioural<br>• Emotional<br>• Socio-economical<br><br>Some common reasons for poor adherence:<br>• Non-disclosure<br>• Economic barriers for accessing care<br>• Alcohol or drug abuse<br>• Feeling better so clients think they don't need the ARVs<br>• Changing guardian for paediatric ART care<br>• Side effects causing patients to stop their medication | 1. Drug interactions causing sub-therapeutic levels of ARVs: i.e., rifampicin, traditional medicines, anti-epileptics (any enzyme-inducing drug)<br>2. Inadequate absorption leading to sub-therapeutic levels of ARVs (i.e., chronic diarrhoea or vomiting)<br>3. Side effects – patient therefore not taking their drugs as prescribed<br>4. Previous exposure to ART (including PMTCT) resulting in primary resistance<br>5. Any defaulter or anyone who had to stop AZT- or D4T-based ART without tail protection resulting in resistance<br>6. Not changing the ARV dose for weight in paediatric patients |

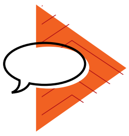

##### Action by the counsellor for a client with VL >1000 copies/ml

The client should be seen by the counsellor to start enhanced adherence counselling on the day the result is given. The high viral load form should be filled at each counselling session as this will facilitate the multi-disciplinary discussion of the client when being considered for second-line treatment.

The first session of enhanced adherence (page 79) should be given on the day the viral load result is given. The goals of enhanced adherence are to discuss possible behavioural, cognitive, emotional and socio-economical barriers to adherence. After presenting the result, the best way to start the session is to ask the patient: What do you think is the reason for your high viral load? In addition, exploring the client's motivation for taking medication often highlights reasons for non-adherence. Clients with high viral load should also have a formal mental health assessment, including the use of alcohol and other substances of misuse.

Two simple screening questions may be asked:

- During the past month, have you felt like you were losing interest or pleasure in doing things?
- Have you felt down, depressed or helpless?

If there is a positive answer to either of these questions or if the client presents with red-flag issues (virological failure, missed appointments, challenging psychosocial issues), carry out a formal mental health assessment using a standardised tool, such as the Shona Symptom Questionnaire (Appendix 3). Clients should then be managed with ongoing counselling interventions or, where appropriate, referred for formal mental health assessment.

The second session of enhanced adherence (page 82) is given four weeks after the first, and is aimed at following up the strategies put in place during the first session. If the client requires a third or more intensive adherence support, this should be provided on a case-by-case basis.

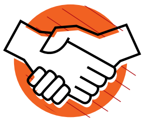

With the client's permission, they should be offered to link with a CHW or expert client in their home area who may be able to give additional adherence support in the community. Clients who are in group models may benefit from having other group members attend their EAC sessions with them.

#### Enhanced adherence session 1

Counsellors should document their findings in the patient care and treatment booklet notes section

| | |
|---|---|
| **Timing** | Session 1: Date high viral load result given to client |
| **Duration** | Approximately 15 minutes |
| **Mode** | Individual |

EAC session prompt Job Aide page 139.

Introduce yourself to the patient.

##### *Step 1: Viral load education review*

Assess patient's understanding of viral load, high viral load and suppressed viral load. Ask the patient to explain to you what each means. If patient requires more explanation, you can say things like:

- The main job/work of your ARVs is to reduce the HIV in your body to a very small amount.
- We can measure this amount of HIV by taking a blood test that we call a viral load test. If ARV treatment is successful, the amount of HIV in the blood will be very low/small/suppressed and you will be healthy.
- The reason it is important to take your medication every day is to make sure that treatment is successful and the amount of virus in the blood is low.
- We have noticed that your viral load is going up. This is not something that can be ignored. We have to find the cause, overcome it, and make sure that your viral load becomes suppressed. We are here to help you achieve this.
- Most of the time, the cause for a high viral load is when you sometimes forget to take your medication.
- Learning to take these medicines is complex, but very possible. Just like learning anything new, it can be overwhelming at first and may take a lot of effort, but with practice, can become part of your daily routine.

##### *Step 2: Discuss the patient's reason/explanation for his or her high viral load*

Sometimes the patient already knows why his/her viral load is going up. Here you can give them a chance to give their own explanation. Often they will already tell you at this point that they are struggling with their adherence.

If they really don't know why their viral load is high, you can say:

- *We notice that when people sometimes forget to take their ARVs every day, it gives the virus a chance to multiply. Do you think that you sometimes forget?*

Make a short note of the patient's explanation. Then move on to the next step. Don't linger too long on this step.

##### *Step 3: Review time the medication is taken (dosing times) & create a medication schedule*

This step is to review the time that the patient has chosen to take their ARV doses. Establish what the patient is doing and where they are at the time they have chosen. For example, if the patient has chosen 9pm, but is already asleep in bed by 9pm, then that is not a good dosing time.

Establish with the patient whether the time they are meant to take their medication is appropriate or whether the time is a problem.

If the time is a problem, then determine a new, more appropriate time with the patient based on their schedule. Remind them that the time chosen is just a suggestion; medicines should always be taken even if not on time.

Then write down the new medication schedule in the counsellor's notes and in their patient-held record.

Other reminders that may be used include a cell phone alarm, a specific TV programme, radio programme, or taking the medication with meals.

***Step 4: Plan for storing medications***

Help the patient identify where at home they are going to keep their medications. If they are afraid of people seeing or finding the medication, then brainstorm a good place to hide them.

- Storage place: ____________________

**Deciding on where to keep extra or emergency doses**

Keeping an extra supply of tablets in specific places is always helpful in emergencies.

Help the patient identify where they can keep an extra supply of medication in case they don't get home in time to take their medication This could be: handbag, locker at work, backpack, wallet, jacket pocket, briefcase, car, etc.

With women, you might identify their handbag as an item they always carry with them.

These tablets are then only to be used when not home in time to take the next dose.

- Extra/emergency supply will be carried in: ____________________

***Step 5: Motivation cards***

This step can help patients learn strategies for remembering to take medications and for thinking helpful thoughts each time they look at their tablets. It is especially helpful for patients who have treatment fatigue, are depressed or are stigmatising themselves.

Introduce the patient to the notecard. Ask the patient to think of their own personal goals/dreams for their future. What are the 3 most important things they still want to achieve in their future?

Have them write it in their own language on a notecard:

e.g. "I want to see my children grow up", "I want to be healthy for my job"

Ask the patient if they think that their ARVs can help them achieve these goals for the future? The answer will be yes, because ARVs will prolong life.

**Encourage the patient to place the notecard where they will read it every day, preferably right before they take their medication.** This will associate taking ARVs with the positive things they want for their future.

- Top 3 goals for the future: ____________________

____________________

____________________

- Do you think that your ARVs can help you achieve your goals for the future?

***Step 6: Discuss patient's support system***

Has the patient disclosed his or her status to any family, friends, or co-workers?

You can ask the patient:

- *Do you have any people in your life who you can talk to about your HIV and ARVs?*
  Suggest to the patient that they enlist the support of their family, friends and co-workers in reminding them to take their medication if they have not already done so.
- The members of the patient's support system are: ____________________
  If they have not disclosed to anyone, write "none".

*Step 7: Planning for substance use*

In the past, the message given to patients was that they shouldn't mix ARVs with alcohol or drugs; the result is that patients decide not to take their ARVs on the day that they use alcohol or drugs. In time, we can support the patient to stop abusing alcohol or drugs, but for the meantime we want to help them to adhere to ARVs while using alcohol or drugs.

You can ask the patient in a casual way (not in an accusing way) if they sometimes like to have a few drinks.

Explain to the patient:

- "We know now that taking ARVs together with alcohol or drugs in not a problem."
- "Taking alcohol or drugs sometimes makes it difficult for us to remember to take treatment. If possible, it is best to limit your use, but if you are planning to take any alcohol or drugs, it is important to plan ahead so that you don't forget to take your treatment."
- "Can you think of ways to still remember to take ARVs while drunk or high?"
- "It is a good idea to take ARVs before you start drinking, even if it is before your schedule dosing time."
- "If you are already out, ask a friend who is not drinking to make sure that you take your ARVs."
- "Ask your wife or a family member to bring your medication to you and remind you to take them on time."
- "If you feel that your alcohol or drug use is affecting your adherence, would you feel ready to be referred to some professionals that may help you to work on that problem?" (Refer this patient to alcohol support service if available.)

Write the patient's plan down in the counsellor's notes.

*Step 8: Getting to your clinic appointment*

- This step helps the patient solve problems associated with getting to his or her appointments.
- Make a plan for getting to appointments:
- How do you get to your medical appointments?
- What would you do if your usual way of getting to your appointments was not an option (e.g., if there was a taxi strike or it was raining when you usually walk)?
    - How do you usually get to clinic: ______________________________
    - Back-up plan: ______________________________
- If they are not able to come on appointment date: remind the patient that if they are unable to make their appointment, they must make sure to go to the clinic the next day, BEFORE they run out of medication.

*Step 9: Review plans and plan way forward*

- Briefly summarise plans made above.
- Identify the steps that the patient needs to complete at home before your next visit, i.e., placing emergency doses in their handbags, their new dosing time, etc.
- Give a short motivational summary on how you believe in the patient! You know they can do this! Together you will make sure that they suppress their viral loads!!

**Plan a way forward**

- Inform the patient that they will be seen after 4 weeks.
- VL will be repeated after 12 weeks.
- VL results will be reviewed together and a way forward will be discussed.

| Enhanced adherence session 2 | |
|---|---|
| **Timing** | Session 2: One month after 1st EAC |
| **Duration** | Approximately 10 minutes |
| **Mode** | Individual |
| **Tools** | EAC session guide of Job Aide Page 139 |

***Step 1: Identify any difficulties with plans & problem solve any new issues***

- Review action plan from previous session: e.g., motivation card and emergency doses.
- Ask the patient if he/she thinks that adherence has improved in the last month. Enquire in a friendly way if any doses have been missed.
- If the patient experienced any difficulties implementing the plans, brainstorm solutions to the identified problem.
- Also problem solve any new issues that may have come up in the past month.

***Step 2: How to learn from mistakes***

- This step may help clients prepare to recover from missing doses, which, in the long run, is likely to occur.
- If a mistake occurs, the best choice is to return to one's adherence programme as soon as possible instead of acting on hopeless thoughts and giving up.
- Identifying what led to the mistake can provide important information that can help avoid future mistakes.
- It should be stressed that mistakes are normal and not a big problem. They only become a big problem when they lead to giving up.
- It is important to tell the patient that they must not beat themselves up if they miss a dose. They must tell themselves that they are only human and that mistakes happen, but that they must return to their medication schedule as soon as possible. If they continue to have many mistakes, then the patient must speak to their medical team as soon as possible.
- Make a plan with the patient:
  - Positive thoughts you can think after you made a mistake ______________________________

    ______________________________________________________________
  - What can you learn from a mistake that will help you avoid another in the future? ______________________

    ______________________________________________________________

    ______________________________________________________________

    ______________________________________________________________

***Step 3: Check your notes to see whether the patient has been referred to other services – if not, skip this step***

- This includes referrals to psychology services, substance abuse groups, social services, etc.
- Ask patient if they attended the appointment? Assure them that if they answer "no", the topic will not be brought up again during these sessions (e.g., we won't force them to go to substance abuse groups).
- If they answer "yes", then check in on their experience with the referral services.

***Step 4: Preparing for travel***

- Holidays are always a risk for poor adherence or default of treatment. Encourage patient to plan for holidays, to make sure that they have enough medication on hand before they leave town, and to remember to pack it !
- Make sure that all relevant information is on the patient's notebook – clinic's phone number, patient's current regimen and doses, latest CD4 and VL, etc.
- Explain to them that if they are ever away from home and they run out of medication, they must go to the closest ARV clinic and show their patient notebook. Hopefully that clinic can help them access medication.
- As back up, have the patient programme their local clinic phone number and file number into their phone. This way, they have it on their phone in case they lose their patient notebook.
- Save on phone: Clinic number, My folder number
- Identify where the patient usually travels to and ask if they know where the closest ARV clinic is.

***Step 5: Review plans***

- Give another short motivational discussion on how you believe in the patient! You know they can do this! Together you will make sure that they suppress their viral loads!!
- Book for repeat VL in 2 months. Give 2-month supply of ART.
- If additional EAC sessions are needed, schedule appointment earlier.
- Repeat viral load should still be taken 12 weeks after result was given.

#### Action by the clinician for a client with a first viral load >1000 copies/ml

If the client already has signs of clinical failure, discuss urgently with a doctor to decide on future management. Adherence will have to be assessed, but factors such as duration on treatment and previous ART exposure will have to be taken into account when assessing likelihood of resistance and the urgency to switch to second line regimen. If clinically indicated switch may be made on the basis of the first viral load. Each case should be assessed individually.

The clinician should read the findings of the counsellor, and also make their own assessment of adherence. The clinician should also make a thorough clinical assessment. All the standard steps of an ART follow-up consultation should be completed, but in addition the clinician should:

- Perform a thorough screening for TB.
- Assess if there are any symptoms or signs of other OIs.
- Ask: Has there been any history of chronic vomiting or diarrhoea now or in the past (possible malabsorption)?
- Ask: Has there been use of any traditional medicines or other medication that may interact with ARVS (e.g., rifampicin, carbamazepine, phenytoin)?
- Ask: Has a child received inadequate dosing of ARVs at any point?

Findings should be noted in the patient care and treatment booklet and the high viral load form.

The client should then be given a one-month supply and booked for the second session of enhanced adherence.

At the next appointment, the client should be assessed again clinically and can be given a two-month supply. They should be booked for further EAC sessions as required, but should have a repeat viral load taken 12 weeks from when the first VL result was given.

#### 2.7.4 Programmatic strategies to ensure completion of EAC and repeat VL testing

Ensuring that enhanced adherence sessions are completed and the repeat VL taken is a challenge. While enhanced adherence is ongoing, each appointment date should be entered into the EPMS. **Every appointment for a client with their last VL >1000 copies/ml should be flagged on the EPMS appointment list.** Hence, anyone appearing with this flag will either need EAC or be due their repeat VL test. If a client defaults on an appointment for EAC or repeat VL testing, they should be traced as per the standard defaulter tracing system.

District mentorship teams should be supplied with a list from the laboratory indicating for each site the clients with a VL >1000 copies/ml. **Mentors may then use these lists of high viral load clients to assess whether clients have completed EAC and repeat VL testing.** It is suggested that mentors check the patient care and treatment booklets of these clients four months after the initial high viral load (i.e., results received in January, check in April) to see the outcome of EAC and repeat VL testing.

#### 2.7.5 Clinical action plan for the repeat viral load result

##### Repeat VL is <1000 copies/ml

If the repeat viral load 12 weeks after the result has been given has suppressed to less than 1000 copies/ml, the client should be congratulated and can continue on their first-line medication. Discuss the ongoing importance of good adherence and the goal of maintaining a viral load less than 1000 copies/ml. They should return to or be offered their differentiated ART delivery refill option of choice.

##### Repeat VL is >1000 copies/ml

If the second viral load remains more than 1000 copies/ml, the client's case should be discussed in a multi-disciplinary approach. This does not have to be at a formal meeting, but it must be ensured that the clinician and counsellor in the clinic have discussed the outcomes of the EAC sessions and clinical review. The completed high viral load form may act as a summary for this discussion.

Once the clinic team has discussed the case, the client should then be discussed with the trained nurse switcher, nurse mentor or doctor, either at the next clinic mentorship visit if it will be **within the next 1-2 weeks** or by telephone. To guide decision making, the high viral load summary form may also be sent to the nurse mentor or doctor by WhatsApp or by the sample transport system.

The approved nurse switcher, nurse mentor or doctor will then make the decision whether to switch based on whether there are significant adherence challenges (child with no caregiver, severe substance misuse or mental health issues) and the clinical condition of the client, including where possible, the current CD4 level.

Once the decision is made, second-line should be initiated in the patient's primary clinic, avoiding unnecessary transportation costs for the client.

**A client should not wait more than two weeks** from the repeat viral load result being available to having a decision and appropriate initiation of second-line therapy. Completion of the viral load algorithm, from the date the first viral load is taken to a final decision being made **should not take more than six months.**

#### 2.7.6 Switching to second-line therapy

The preferred second-line treatments for adults and children can be found on page 77 and 80 of the Job Aide.

##### Counselling preparation for second-line

In the same way that a client must be prepared for starting their first-line regimen, they must be prepared to start second-line. These clients are often those who have had challenges with adherence and it is particularly important that adherence to second-line is optimal from the start (See details of the counselling session on the next page).

##### Clinical preparation and follow up for second-line

Prior to switching to second-line, clients should have a full history and examination performed, including a thorough screening for TB.

If TDF is in the first-line, check hepatitis B serology. If hepatitis B surface antigen is positive, TDF must be continued in the second-line.

If available, check glucose and lipids.

Follow up then follows the same schedule as when first-line was initiated, month 1, 2, 3, 6 and 12 on second-line. If clinically well and virologically suppressed after 6 months on second-line, the client can return to a differentiated ART refill system for stable clients.

#### Second-line preparation session

***Provide repeat (second) viral load result***

- If VL is >1000 copies/ml and there are no major adherence barriers, switch to second-line. The decision to start second-line ART is taken as a team (nurse & counsellors), supported by the mentors. It does not require a specific meeting.
- How does the client feel concerning the result?

***Give general info on second-line treatment***

- Explain how second-line treatment consists of other drugs, which will be able to fight the virus, if taken correctly.
- Explain the benefits of second-line treatment: CD4 will increase, OI will decrease, and viral load should be undetectable.
- Explain that second-line treatment can have some side effects (yellow eye with ATV/RIT; dizziness, breathlessness with AZT).
- Explain the need for good adherence on second-line treatment. If not, the client will get resistant to these drugs also, and there are limited options for other treatment.
- Revise strategies identified during EAC on how to ensure good adherence.

***Assess readiness to start second-line treatment***

Counselling and clinical follow up after second-line initiation is the same as for first-line initiation. Undertake to follow the client and give adherence support at:

- Months 1 (M1)
- M3
- M6 – emphasise that client will be bled for viral load
- Then to follow the refill option of choice if fits eligibility criteria

#### 2.7.7 Access to third-line antiretrovirals

Clients failing second-line antiretroviral regimens (i.e., have had two consecutive viral loads more than 1000 copies/ml three months apart and after an adherence intervention) should be referred to the tertiary level for assessment for genotyping and assessment for possible third-line regimen.

#### 2.7.8 Special considerations for children and adolescents with high viral load

Children and adolescents have higher rates of treatment failure than adults. Children are usually dependant on a caretaker for administration of their ARVs and, for many who are orphaned, that caretaker often changes or is elderly. When a child is identified with a high viral load, the multi-disciplinary team at the clinic should discuss this case and **consider a home visit**. An assessment by a social worker may also be needed. Investigating further **community support** for this child should be considered. There may be another adult or peer on ART living in the same community who may be willing to support the child and their family as a daily **treatment buddy** to improve adherence to medication. Another common barrier to adherence for children and young adolescents is non-disclosure. Working with the caretaker to fully disclose is therefore an essential step in the enhanced adherence process for a failing child (see pages 125-137 of the Job Aide).

Ensuring this child's follow up through enhanced adherence and repeat viral load is imperative. If the second viral load remains more than a 1000 copies/ml, the child's case should be discussed with the nurse mentor or doctor as soon as possible. If on a PI based regimen non-adherence is more likely than resistance, however discuss with an experienced clinician to decide whether genotyping may be feasible.

#### 2.7.9 Special considerations for viral load monitoring in pregnant and breastfeeding women

Management of high viral load in pregnant and breast feeding women impacts not only on the woman, but also the baby. Hence viral load monitoring and the management of high viral load has been differentiated for this population.

For ART naive pregnant or breast feeding women, the first VL should be taken after 3 months on ART.

For women already on ART check the VL at the first ANC visit.

If any pregnant or breastfeeding woman has a VL > 1000 copies/ml start enhanced adherence and repeat the VL after one month.

If the repeat VL remains greater than 1000 copies/ml prepare to switch regimen.

### CHAPTER 3

#### Pharmacy, Laboratory and Strategic Information

### 3.1 Pharmacy

Access to quality and affordable ARVs and essential medicines is a fundamental component of any HIV prevention, care and treatment programme. With the increasing number of clients requiring ART with the introduction of "Treat All", the supply systems needed to ensure a sustained supply of medicines will have to be strengthened. The following section outlines the key points that must be considered for effective pharmacy management related to the HIV prevention, care and treatment programme.

**If stock levels are equal to one month or less, facilities MUST place an EMERGENCY ORDER**. In order to do this, the pharmacy technician, pharmacist or nurse in charge must ensure that stock cards are kept correctly, that transactions are recorded as they occur, and that minimum and maximum stocks levels are updated regularly. They should contact the district hospital pharmacy manager by the quickest possible means (phone, email, fax, visit) if stocks fall below the one-month supply level.

#### 3.1.1 Duration of supply to clients

Stable clients should receive **a three-month** supply of cotrimoxazole.

Stable clients should receive **a three-month** supply of ARVs.

It is good practice to review the ART needs for the coming month (especially for paediatric ARVs or clients on second-line or less common regimens) in order to ensure that the facility will be able to supply the clients. If NatPharm has not delivered an emergency order, the district pharmacy manager should be contacted or arrangements be made with local clinics to borrow drugs. If this is done, it should be clearly documented in the pharmacy records.

Clients should be given a shorter duration of cotrimoxazole or ARVs for their refill only if efforts to receive an emergency order from NatPharm or receive supplies from another facility have failed. If this happens, it should be reported to the district pharmacy manager. The reasons for this short supply should be explained to clients so they understand that this will be followed up and that all efforts will be made to ensure a three-month drug supply for stable clients. The role of the community in monitoring the supply chain of ARVs and essential drugs will also be a mechanism to report supply problems back to programme managers.

#### 3.1.2 Pharmacy requirements for decentralisation

In order to become an accredited ART site, certain requirements must be met regarding pharmacy management. These requirements can be found in the *Manual for Primary Health Care Facility Comprehensive HIV/AIDS Capacity Assessment.* The District Health Executive (DHE) has the responsibility of ensuring that the facility meets these standards.

##### Ordering and supply

Reporting and ordering for both ARVs and essential drugs should be performed every three months, using the appropriate standard operating procedure for the level of the facility. To ensure adequate supplies, accurate documentation in the ART pharmacy registers is essential, along with accurate reporting of ART data and consumption documentation. All reports should be submitted by the due date to avoid delays in receiving supplies from the National Pharmaceutical Company of Zimbabwe (NatPharm).

NatPharm will supply **up to six months of ARVs** and essential medicines to the clinics.

NatPharm will **directly deliver** both ARVs and essential drugs to the clinics **every three months**.

#### 3.1.3 Pre-packing of medication for distribution

In certain refill models as described in Section 2.5.4, pre-packing of ART prior to the refill facilitates faster distribution by a nurse, e.g., during a facility-based group refill, or allows a primary counsellor, community health worker, expert client or community ART group member to distribute medication to other clients.

Medication should be dispensed according to the refill prescription. Ideally, each box of ART should be labelled with the client's name and all medication placed in a bag, which also is labelled with the client's name.

#### 3.1.4 Specific drugs for opportunistic infections

##### Fluconazole

Fluconazole is a B level drug (i.e., it can only be initiated at the hospital).If a client is initiated on fluconazole for either oesophageal thrush or for cryptococcal meningitis and is then referred back to their local primary care clinic, the referring doctor must ensure that clear documentation regarding dosage and duration of treatment is made in the client notebook. In addition, the district pharmacy manager must ensure that an adequate supply of fluconazole is sent to the receiving clinic. This requires improved communication between clinicians and pharmacy management.

##### Aciclovir

Aciclovir is a C level drug. Aciclovir should be available for treatment of both genital herpes and herpes zoster at all health facility levels.

#### 3.1.5 Key messages and reference materials

- Three-monthly supplies of ART and cotrimoxazole can be given for stable clients.
- Decentralised primary care clinics must meet the pharmacy requirements as outlined in the *Manual for Primary Health Facility Comprehensive HIV and AIDS Capacity Assessment.*
- Clinics should follow the ZAPS SOPs for ordering and hospitals the ZADS SOPs.
- Clear documentation of prescriptions dispensed should be made in the ART pharmacy register and patient care and treatment booklet.
- Reporting, ordering and supply of drugs will be performed every three months.
- **If stock levels are equal to one month or less, facilities MUST place an EMERGENCY ORDER.**

##### Reference materials

Zimbabwe Assisted Pull Systems (ZAPS) Standard Operating Procedure.

Standard Operating Procedures Manual for the Zimbabwe ART Distribution System (ZADS).

### 3.2 Laboratory

#### 3.2.1 Background

To implement the new clinical guidelines and strengthen service delivery across the cascade, increased support to laboratory and diagnostic services will be needed. Quality assurance systems for all testing services must be in place and acted on. The challenge of an increasing number of point-of-care tests deployed at multiple settings also poses challenges for supervision and quality assurance.

#### 3.2.2 Supporting a dedicated sample transport system

Sample transportation is an essential part of the provision of the HIV prevention, care and treatment minimum package. Not only do samples have to be delivered to the central laboratory, but it also serves as a mechanism for result delivery. Specimen collection should be integrated across programmes in order to maximise efficiencies. The MoHCC is currently developing a strategy for such an integrated sample transport system.

Different settings may have different challenges regarding sample transport, but each district should have a **written plan** as to how **reliable weekly** sample transport from each decentralised site is provided. The district receiving laboratory should be responsible for the sample transportation schedule to ensure specimens are received throughout the week and the laboratory is not overloaded on any particular day.

Some possible solutions include:

- Employment of one or two dedicated personnel or assignment of one or two environmental health technicians (EHTs) for the task of sample transportation for the district. Scheduling is organised by the laboratory receiving the samples.
- Several EHTs are responsible for assigned clinics (a solution for when they are away on other duties must be found). The district laboratory should still be responsible for the scheduling of these EHTs.
- Clinics delegate a nurse aid, general hand or volunteer to take specimens to the laboratory via public transport. The overall cost of this for the district should be analysed.

> Whatever the sample transport system, there must be a plan for weekly specimen collection from all sites that is overseen by the district laboratory in charge. The system should be regularly monitored for efficiency.

If motorbikes are used, a district plan for maintenance and fuel should be clearly documented in the sample transport strategy.

#### 3.2.3 Quality management systems

Ensuring a comprehensive quality management system, including internal and external quality control, is essential.

The quality management system should:

- Be implemented within the laboratory network and all remote testing sites
- Be incorporated into the routine testing procedures, and monitored
- Ensure that all testing sites undertake quality control
- Ensure that all testing sites are enrolled in an external quality assessment scheme (proficiency testing programme)
- Ensure the use of standard operating procedures for all processes, including specimen collection and processing, test methods, interpreting results and reporting. Where testing has been decentralised, these SOPs must be ensured through regular site visits
- Ensure the use of standardised logbooks or electronic data management and reporting, including identification of errors and non-conformances
- Ensure that all equipment at all facilities is maintained with both preventive and curative actions
- Ensure regular competence assessment for all testers.

#### 3.2.4 Role of the laboratory in mentorship and supportive supervision

When planning for district mentorship and supportive supervision, the district (or provincial) laboratory in charge **must be included**. Ongoing implementation of training and quality assurance **will require scheduled visits to all sites.** When new interventions, such as viral load, are introduced, close liaison between the clinical mentoring staff and the laboratory in charge will be needed.

#### 3.2.5 Key messages and reference materials

The minimum package of laboratory investigations should include:

- **At primary facility:** HIV testing kits; DNA PCR kits; pregnancy tests; syphilis rapid tests; Hb; urine dipstick; and any available point-of-care technology where appropriate (i.e., CD4, EID or viral load)
- **At district level, all of above plus:** TB diagnosis (smear or Xpert MTB/Rif); CRAG testing for blood and CSF (at minimum access to Indian ink); creatinine (TDF use); ALT (NVP use); CD4 (at least for baseline); and hepatitis B and C screening
- **At provincial or central level, all of above plus:** Viral load (this will be gradually phased in)
- **At national level:** Genotyping; and TB culture and drug sensitivity testing
- A dedicated weekly, reliable sample transport system should be in place for all facilities providing the minimum package of HIV prevention, care and treatment services (Section 3.2.2).
- Health workers who perform a test (any rapid or point-of-care test) must be adequately trained for that task
- Quality assurance (internal and external) must be in place for all tests and all sites performing those tests
- The laboratory scientist/technician must participate in the district mentorship and supportive supervision teams and have scheduled visits to all sites.

### 3.3 Monitoring and evaluation

#### 3.3.1 Background

Monitoring and evaluation is an essential part of the programme cycle (Figure 10). We assess a client's health by taking a history and examination, design a treatment plan, implement the treatment plan, monitor the client's progress, and evaluate the success of treatment. We should take the same steps when we are managing facility activities.

Monitoring and evaluation (M&E) has three main purposes:

- M&E helps us make informed **decisions** for programme and policy planning
- M&E allows us **assess our performance.** Often our performance is assessed against set targets for specific indicators
- M&E provides **accountability**. This may be required for reporting back to stakeholders.

#### 3.3.2 Data management

There are four steps in data management:

- Data collection
- Data quality assessment (accuracy, completeness, timeliness, validity, reliability)
- Data analysis and interpretation
- Data for decision making and dissemination.

It is essential that every facility designates someone who is responsible for the data management process.

Details of how to enter data both in the paper registers and in the electronic patient medical systems (EPMS) can be found in the monitoring and evaluation training manual provided in the reference materials.

Data must be submitted **in a timely manner** according to the protocols. At every level (facility, district, provincial to national), the person responsible must ensure that the necessary quality checks have been performed and that the data has been verified. Figure 11 outlines the flow of reports and timelines.

##### Figure 10: The programme cycle

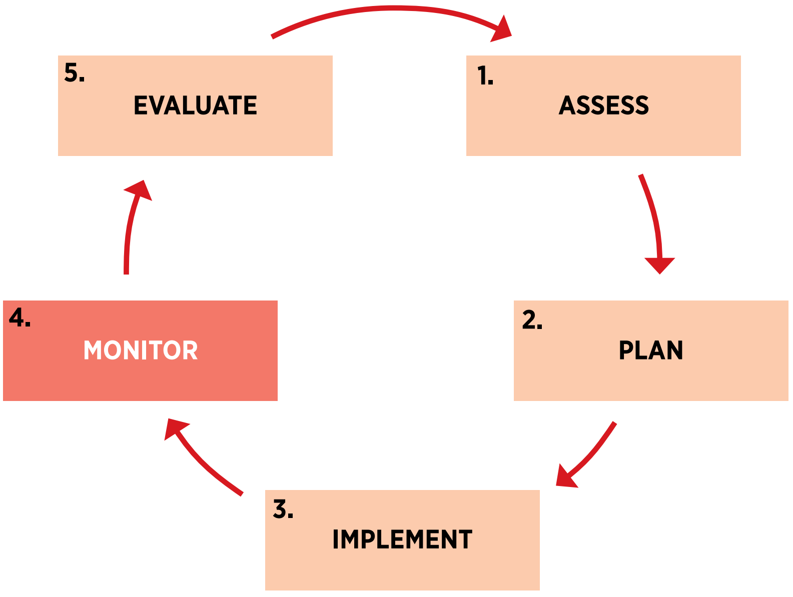

#### Figure 11: Expected flow of reports and timelines

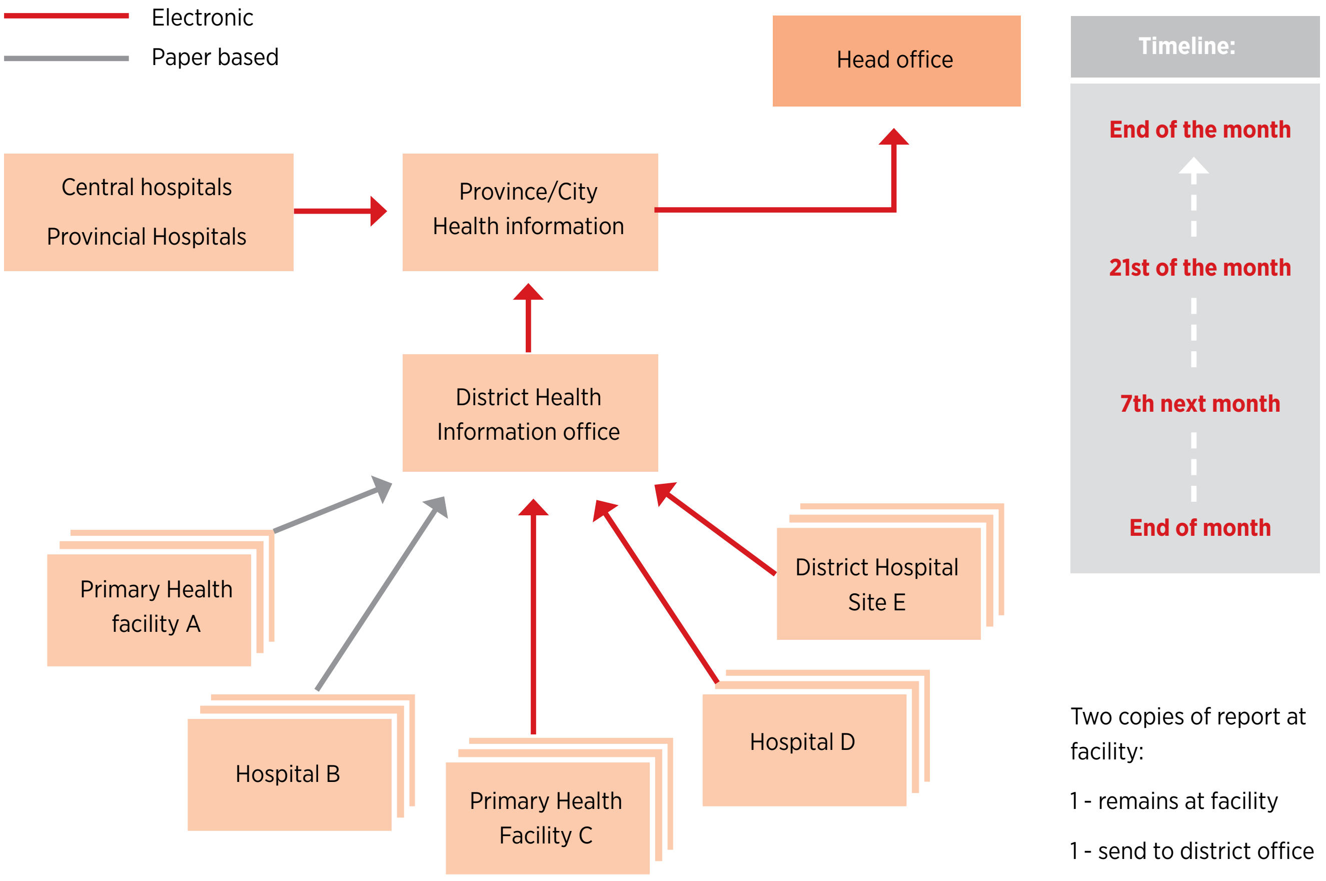

**At facility level, there are two copies of the report: one stays at the facility; one is sent to the district office.**

In addition to ensuring that the data from the facility is submitted correctly, there must be strengthened **coordination between the facility and any community-based activities** in order for those activities to be reported under the facility catchment area.

##### 3.3.3 Data dissemination

It is very important that we share and use the data we collect. Disseminating data is positive for:

- Transparency
- Accountability
- Sharing experiences
- Demonstrating our achievements against set targets.

All sites must provide **a plan for dissemination of their data at their facility and to their DHE**. Possible options for doing this include:

- Use regular staff meetings to discuss what the data could be showing you; brainstorm on ideas for why a certain trend is happening.
- Plan a regular quarterly meeting at district level to share data among facilities. Use these sessions to see how other facilities are performing (e.g., how many paediatric initiations were performed or how many DBS samples performed were positive). Use the data to brainstorm and share experiences.
- Use the data to inform the community about performance at the health facility. For example, are HIV testing and counselling rates decreasing, are very few men coming for testing, or are women coming very late to ANC? Use this data to encourage community mobilisation on these issues.
- Use the data to advocate and lobby for change. For example, workload data may allow lobbying for additional human resources.

#### 3.3.4 Supporting national surveys

Facilities will be expected to participate in national surveys, such as the HIV drug resistance, the transmitted drug resistance, HIV sero-surveillance, ANC and adherence and retention surveys.

#### 3.3.5 Key messages and reference materials

Monitoring and evaluation is performed in order for us to assess how effectively we are delivering services in our clinics, districts, provinces and at national level.

- Data must be collected, verified, analysed and then disseminated. If data is not fed back to the staff doing the job, it will not benefit client care. Each facility should have a plan for data dissemination.
- Track some simple indicators on a monthly basis using graphs in a cascade format on the wall (similar to how EPI activity is monitored), e.g., estimated number of people living with HIV, number of newly diagnosed HIV patients, number enrolled in care, number initiated on ART.
- Each facility must have a focal person for M&E who must have received adequate training.
- Data must be submitted at the correct time across all levels.

Monitoring and Evaluation Training Manual for HIV Testing and Counselling, Sexual and Reproductive Health prevention of mother to child transmission of HIV, opportunistic infections/ ART, TB and male circumcision.

### 3.4 Quality improvement and implementation research

#### 3.4.1 What is quality improvement

**Quality** in health care is defined as proper performance (according to standards) of interventions that are known to be safe, that are affordable to the society in question, and that have the ability to produce an impact on mortality, morbidity, disability and malnutrition.

**Quality improvement** is an interdisciplinary process designed to raise the standards of the delivery of preventive, diagnostic, therapeutic and rehabilitative measures in order to restore and improve health outcomes of individuals and populations (*American College of Medical Quality*).

This section gives a brief overview of the core principles of quality management.

##### Principles of quality improvement

- Understanding work in terms of processes and systems
- Developing solutions by teams of health care providers and clients
- Focusing on client needs
- Testing and measuring effects of changes
- Shared learning.

##### Benefits of quality improvement (QI)

- Reduces morbidity and mortality of clients
- Reduces health care costs and waste of resources
- Enhances client satisfaction – provides care that is responsive to clients' and communities' needs and expectations
- Improves safety of staff, clients and communities
- Cultivates teamwork and effective communication
- Provides good reputation of health institutions and health workers
- Improves staff motivation.

##### Rationale for quality improvement

- QI tools provide a simple, systematic way to monitor, assess and improve care
- Improves quality for the majority of clients, not just the tough cases
- Uses real-time clinic performance data to guide changes
- Takes the time to assess how care is being provided – not just counting
- Improves systems, not just individual provider performance
- Fosters learning from peers and spreading of effective practices.

#### 3.4.2 Plan-do-study-act (PDSA) cycle

The process by which quality improvement interventions are guided is known as the plan-do-study-act (PDSA) cycle. The PDSA cycle guides the implementation of a change to see if the change is an improvement.

- **Plan (plan a change).** The team identifies a change and plans how they will implement this change.
- **Do (try it out on a small scale).** The team members test the proposed change to see whether it results in an improvement.
- **Study (observe the results).** Once the results are analysed and reviewed, the project team answers the following questions: Did we meet our goal? What worked and what didn't? Do we need additional test cycles?
- **Act (refine the change as necessary).** The team maximises the impact of successful changes by increasing the sample size involving providers and expanding the test cycles.

#### 3.4.3 Steps for implementing a quality improvement plan

1. Agree on what needs to be done by answering the question, "What are we trying to accomplish?"
2. The goals and targets should be set according to the needs of consumers, as well as the national standards/guidelines.
3. Sensitise all the stakeholders, including consumers, on the goals and objectives to be achieved.
4. Set up a project team.
5. Measure current performance and determine the performance gap.
6. Develop indicators and agree on measurement cycles.
7. Abstract, validate, analyse, report and disseminate the results.

8. Assess the capacity of the organisation to improve care.
9. Identify problems and causes of the performance gap.
10. Carry out process mapping.
11. Do root cause analysis (brainstorming, 5 WHYs, fishbone diagram).
12. Generate decision matrix for prioritising the problems and the possible interventions.
13. Develop an improvement plan.
14. Outline how (activities/ processes), by who (responsible person), when (specific timelines), using what (resources required) and where (location/ department).
15. Implement the improvement interventions according to the plan.
16. Support through coaching and mentoring, as well as peer learning.
17. Review progress.
18. Measure performance at the end of the cycle.
19. Get feedback from consumers and other stakeholders.
20. Disseminate the results to all stakeholders.
21. Sustain the improvements.

#### 3.4.4 Implementation research

Implementation research is aimed at answering questions raised in real-world conditions with populations being affected by an intervention. In HIV/TB health service provision, this applies to questions being faced by the service providers on the ground. These questions may be developed through use of health information systems to track cases or the impact of prevention and treatment. Implementation research is especially concerned with the users of the research who may be programme managers and clinicians on the ground. The outcomes of implementation research should lead directly to actions and policies to promote improved health service provision.

Some key questions to assess research designs or reports on implementation research are:

- Does the research clearly aim to answer a question concerning implementation?
- Does the research question clearly identify the primary audience for the research and how they would use the research?
- Is there a clear description of what is being implemented?
- Does the research involve an implementation strategy?
- Is the research conducted in a real-world setting?
- Does the research appropriately consider implementation outcome variables?
- Does the research appropriately consider context and other factors that influence implementation?

HIV/TB-related implementation research can be carried out by any health care worker in any MoHCC health facility under the guidance of the MoHCC. A research framework has been developed for HIV/TB, which may help guide those who are thinking of developing implementation research proposals. For further information or support for carrying out implementation research, please contact the Operations Research Fellow in the AIDS and TB Unit.

#### 3.4.5 Key messages and reference materials

Quality improvement is an essential element of running HIV prevention care and treatment services.

Involving health care workers in the process of quality improvement encourages accountability for assessing the quality of their service and engagement in the processes to assess and implement quality improvement changes.

##### Reference materials

National Quality Improvement Programme Guide: for the improvement of HIV prevention care treatment and support services in Zimbabwe.

### APPENDICES

### Appendix 1:
### AIDS and TB programme referral form

| | |
|---|---|
| Part A: Services referred for to be filled out by the organisation making the referral | |
| **Please receive (Client Name):** | **Referring Officer(Name and Contact Details)** |
| **Sex:** M F **Marital Status:** Married Single | **ID Nº** *(Which could be ANC/PMTCT/ART or national ID number)* Please indicate: |
| **Age:** | **Community/Facility:** |
| **Contact No.**: | **Date Referred** **Expected Visit Date:** |
| **Referred to (site/department):** | |
| Circle below the services you are referring this client for. If other please specify | |

| | | | |
|---|---|---|---|
| **01 Pre-ART registration** | **07 ART official transfer** | **12 TB screening/managem** | **17 FBC** |
| **02 ART Initiation** | **08 ART reinitiating (LTFU)** | **13 Family Planning** | **18 LFT** |
| **03 ART refill (defaulters)** | **09 CD4 Count** | **14 VMMC** | **19 U&Es** |
| **04 ART Decentralization** | **10 Viral Load test** | **15 STI –screening /treatme** | **20 HIV Rapid Test** |
| **05 PMTCT** | **11 HIV Rapid Test** | **16 CARG enrolment** | **21 DBS- PCR HIV Testing** |
| **06 CTX/OI Management** | | | |
| **22 Other (Specify):** | | | |

| | |
|---|---|
| **Tick in the box when feedback is received** *(to be completed of the carbon c which stays)* ☐ | **Date Feedback Received:** |

| | |
|---|---|
| Part B: Services Provided to be filled out by the organisation fulfilling the referral *(if at facility tear off and keep in the box for community organization to collect, if at community tear off and return to referring facility)* | |
| We have seen (client's full name): | Client's ID Nº :*(Which could be ANC/PMTCT/ART or national ID number)* Please indicate: |
| **At (Site/Department):** | |
| Circle below the services you have offered this client. If other please specify | |

| | | | |
|---|---|---|---|
| **01** Pre-ART registration | **07** ART official transfer | **12** TB screening/management | **17** FBC |
| **02** ART Initiation | **08** ART reinitiating (LTFU) | **13** Family Planning | **18** LFT |
| **03** ART refill (defaulters) | **09** CD4 Count | **14** VMMC | **19** U&Es |
| **04** ART Decentralization | **10** Viral Load test | **15** STI –screening /treatment | **20** HIV Rapid Test |
| **05** PMTCT | **11** HIV Rapid Test | **16** CARG enrolment | **21** DBS-PCR HIV Testing |
| **06** CTX/OI Management | | | |
| **22 Other (Specify):** | | | |

| | | |
|---|---|---|
| **Comment on services provided:** | | |
| **Name of Service Provider:** | **Position:** | **Date services provided:** |

**PART C: FEEDBACK TEAR OF SLIP** *(slip to be given to the person referring so that they tick referral as complete)*

| Date | Client Name | Services provided(use code) | Further referral (Y/N) | Place & services referred for |
|---|---|---|---|---|

### Appendix 2: HIV re-testing: Frequently asked questions

This factsheet provides answers to FAQs documented during the learning phase implementation of “Treat All” in Zimbabwe. These FAQs are intended to support Ministry of Health and Child Care (MoHCC) health managers and facility-based health care workers to understand and optimise the recommendation to support HIV re-testing in ART services.

#### Q: Why is HIV re-testing before ART initiation being recommended?

A. Recent reports of HIV status misclassification, with both false positive and false negative results, have raised concerns that some individuals might be started on ART inappropriately. To address this concern, Zimbabwe has adopted the WHO 2015 guideline to re-test all persons newly diagnosed as HIV positive with a second specimen before ART initiation.

#### Q: What is the magnitude of HIV status misclassification in Zimbabwe?

A. Globally, misclassification is between 0.2% and 10.5 %. The Zimbabwe 2012 ANC surveillance found that routine PMTCT testing gave incorrect results as follows:

- 8.8% of clients testing HIV positive were eventually confirmed as HIV negative
- 1.3% of clients testing HIV negative were eventually confirmed as HIV positive.

Common sources of error include: technical errors, mislabelling, clerical errors, cross reactivity, incorrect or sub-optimal testing algorithm, and lack of training.

#### Q: Does the recommendation for HIV re-testing indicate a lack of capacity within our health system?

A. Misdiagnosis arises from technical or clerical errors. It does not necessarily mean that the test kits are of poor quality or that they have not been stored properly. Neither does it mean that health care providers are not competent in performing the tests. However, care should be taken to ensure that the integrity of test kits is maintained during transportation and storage. In addition, service providers should exercise extreme care when conducting HIV testing to minimise human error.

HIV re-testing is not about confidence in the system, but it is about ensuring that the right quality care is offered to our clients. Everything done by a human being is prone to error, and HIV re-testing ensures that HIV testing is error proof.

#### Q. How long after the first test is re-testing done?

A. For newly diagnosed HIV-positive clients, re-testing can be done within a few (1-4) hours of the first test. Newly diagnosed HIV-positive clients would be provided with post-test counselling and linked to the health care provider who will initiate them on treatment. The re-test should be conducted by a different provider. A new sample would be collected and tested using the same testing algorithm. The World Health Organization recommends that clients with HIV-inconclusive status (i.e., first test and re-test results are discordant) be re-tested in 14 days.

#### Q. What happens if there is only one nurse on duty and the other is away on leave? Will it lead to delays in ART initiation?

A. If only one nurse is available at the facility, the nurse can re-test the client using a new sample taken a few hours apart from the first sample. Absence of a second provider should not delay ART initiation.

#### Q. What happens to all the clients currently on ART who were initiated without a re-test? Are we going to re-test all clients initiated on ART without a re-test?

A. Clients started on ART without re-testing are NOT going to be re-tested as the presence of ARVs adversely affects the sensitivity of the test kits. ART suppresses viral replication, which may extend to suppression of the immune response, thus reducing HIV antibody production. This may result in negative results in clients who are HIV positive but on ART. Re-testing is therefore not recommended for individuals on ART.

#### Q. Are there enough resources for re-testing given that sites have experienced stock outs of HIV test kits before?

A. The resources for HIV re-testing are available; the MoHCC embarked on a quantification exercise to ascertain the needs. In the long run, it is more expensive to falsely treat someone than it is to re-test. HIV test kits cost approximately US$0.50 per single test, whereas a full year of ART treatment costs US$90-120, without considering the cost incurred by the patient to access HIV treatment.

##### Q: What do you say to a client who had a positive result and then, upon re-test, records a negative result?

A. The client should be informed that their results are discordant, and that there is need to conduct another test. The re-test is to be conducted after 14 days at the same health facility using the same algorithm. If the result of the re-test is negative, then the final result is issued as negative. If the result of the re-test is positive, then the result is positive.

##### Q. Do our laboratories have the capacity to perform re-tests, such as Western Blot or DNA-PCR?

A. At this stage, there is no need for using Western Blot as the re-test. In cases where a re-test is needed, the client can be asked to return after 14 days for re-testing using a third specimen and the same tests.

##### Q. Why not give the client their final HIV result after the re-test?

A. HIV re-testing before ART is a quality assurance and quality improvement strategy. Only a small proportion of patients are expected to be issued with an initial false positive or negative result. For this reason, the initial result has to be issued and a re-test conducted at the point of ART initiation.

##### Q. Will re-testing make clients lose confidence in the service provider?

A. Service providers should explain clearly to the clients the reason and value for re-testing in order to instil confidence in the health system and the HIV results being given. Clients should be told about HIV re-testing during pre- and post-test counselling. Health care workers and counsellors should emphasise that HIV re-testing is a way to reduce human error and avoid starting ART clients who do not need it. If errors are not avoided, HIV services will be expensive for both the clients and health delivery system. It will be important for the MoHCC and its partners to develop a communication strategy on HIV re-testing.

#### Key messages about HIV re-testing

1. Any incorrect diagnosis, whether a false positive or a false negative, can have severe personal and public health consequences.
2. Re-testing ensures that individuals are not needlessly placed on life-long ART (with potential side effects, waste of resources, and psychological impact of misdiagnosis).
3. HIV Re-testing prior to ART initiation should be performed on another sample and ideally by another health care worker.

### Appendix 3: Shona symptom questionnaire for the detection of depression and anxiety

Client name:________________________ Client ID:________________________ Date: ____________

| | Musvondo rapfuura:<br>*During the course of the past week:* | Ehe<br>*Yes* | Aiwa<br>*No* |
|---|---|---|---|
| 1 | Pane pamaimboona muchinyanya kufungisisa kana kufunga zvakawanda here?<br>*Did you sometimes think deeply or think about many things?* | | |
| 2 | Pane pamaimbotadza kuisa pfungwa dzenyu panwechete here?<br>*Did you find yourself sometimes failing to concentrate?* | | |
| 3 | Maimboshatirwa kanakuita hasha zvenhando here?<br>*Did you lose your temper or get annoyed over trivial matters?* | | |
| 4 | Maimborota hope dzinotyisa kana dzisina kunaka here?<br>*Did you have nightmares or bad dreams?* | | |
| 5 | Maimboona kana kunzwa zvinhu zvangazvisinga onekwe kana kunzwikwa nevamwe?<br>*Did you sometimes see or hear things others could not see or hear?* | | |
| 6 | Mudumbu menyu maimborwa dza here?<br>*Was your stomach aching?* | | |
| 7 | Maimbovhundutswa nezvinhu zvisina mature here?<br>*Were you frightened by trivial things?* | | |
| 8 | Maimbota dza kurara kana kushaya hope here?<br>*Did you sometimes fail to sleep or did you lose sleep?* | | |
| 9 | Pane pamaimbonzwa muchiomerwa neupenyu zvekuti makambochema kana kuti makambonzwa kuda kuchema here?<br>*Were there times when you felt life was so tough you cried or wanted to cry?* | | |
| 10 | Maimbonzwa kuneta here?<br>*Did you feel run down (tired)?* | | |
| 11 | Pane pamaimboita pfungwa dzekuda kuzviuraya here?<br>*Did you sometimes feel like committing suicide?* | | |
| 12 | Mainzwa kusafara here mune zvamaiita zuva nezuva?<br>*Were you generally unhappy with the things you were doing each day?* | | |
| 13 | Basa renyu raive rave kusarira muma shure here?<br>*Was your work lagging behind?* | | |
| 14 | Mainzwa zvichikuomerai here kuti muzive kuti moita zvipi?<br>*Did you feel you had problems deciding what to do?* | | |
| | **Scoring : Add together the number of questions to which the client responded “ yes”** | **Total Score:** | |

**Scoring information**

0-7: Re-screen in one year.

8-14: Provide brief counselling intervention. Refer for further assessment and to CBO for psychosocial services.

If a client scores 7 or less but is still suspected of mental health symptoms, they should be considered to have a positive score and receive a brief counselling intervention and referral as appropriate.

Action taken:________________________________

________________________________

Brief counselling: Yes No

Referral: Yes No

Referred to:________________________________

### Appendix 4: Questionnaire to support decision making for differentiated service delivery

#### Differentiated HTS

| M and E | Site |
|---|---|
| Estimated total people living with HIV | |
| Number of people who know their status | |
| Estimated number of adults living with HIV | |
| Number of adults who know their status | |
| Estimated number of children living with HIV | |
| Number of children who know their status | |
| **Where are HTS services offered?** | |
| Is opt-out PITC offered from all entry points ( OPD, IPD, ANC, FP, TB, STI, nutrition)? | |
| Is facility-based index client testing offered? | |
| Is community-based index client testing offered (see OSDM)? | |
| Is targeted outreach testing performed at least once per quarter? | |
| **When are HTS services offered ?** | |
| Are HTS services available during working hours every day? | |
| Are HTS services available overnight and at weekends in maternity and IPD? | |
| **Who is supporting and performing HTS ?** | |
| How many staff are trained to perform HTS | |
| Is self-testing being offered ? | |
| **Differentiated HTS for children and adolescents** | |
| Are HTS services offered at adapted times for children and adolescents? | |
| Is any outreach testing performed to schools or in EPI? | |
| How many staff members are trained to perform DBS? | |
| How many staff members are trained to perform paediatric disclosure counselling? | |
| Are adolescent peers involved in mobilising other adolescents for testing? | |
| **Differentiated HTS for pregnant and breastfeeding women** | |
| Is re-testing of pregnant women offered according to the re-testing guideline? | |
| Is re-testing of negative breastfeeding women integrated into EPI? | |
| **Differentiated HTS for other sub-populations** | |
| Are HTS services adapted for any key populations (e.g., moonlight testing , involvment of peers)? | |
| Are HTS services adapted for men? | |
| Are HTS services adapted for mobile populations? | |
| Are HTS services adapted for people living with disabilities? | |

### Differentiated ART delivery

| M&E | Site |
|---|---|
| Number on ART | |
| Adult retention at 12 months | |
| Adult retention at 48 months | |
| Paediatric retention at 12 months | |
| Paediatric retention at 48 months | |
| PMTCT retention at 12 months | |
| PMTCT retention at 48 months | |
| Is there an appointment system? | |
| Is the defaulter tracing standard operating procedure carried out? | |
| **Health care workload** | |
| How many patients does each HCW see on an ART day? | |
| How many days of the week is ART given? | |
| From what time is ART provided and until what time? | |
| Health care workers perception of workload | |
| **Patient barriers** | |
| How far are patients travelling to reach the clinic? | |
| What are the costs of transport for clients? | |
| How long do patients wait from when they arrive to when they leave? | |
| What are your biggest challenges to accessing ART? | |
| Describe the 5 refill options to a group of clients. Which options address clients' challenges? Is more than one option warranted? | |
| **Differentiated ART delivery for stable clients** | |
| What maximum refill (3 months) is given routinely for stable patients? | |
| How many days of the week are ART refills offered? | |
| Are extended hours offered for ART refills? | |
| Do patients see the nurse every visit or are clinical and refill visits differentiated? | |
| What is the schedule for clinical follow up in the clinic? | |
| What is the schedule for counselling follow up? | |
| Where is ART delivered? (facility or community) | |
| Is there any fast-track refill option? | |
| Is there any club refill option? | |
| Is there any outreach refill option? | |
| Is there a CARG refill option? | |
| Is there any family refill option? | |
| Describe the 5 refill options to a group of HCWs. Which options address their and clients' challenges? Is more than one option warranted? | |

| |
|---|
| **Differentiated ART delivery for children and adolescents** |
| Is a family approach offered for children age 0-5 in MnCH? |
| Are refill clubs offered for adolescents? |
| Are adolescent peers involved in adherence support and defaulter tracing? |
| **Differentiated ART delivery for pregnant and breastfeeding women** |
| Is PMTCT and ANC integrated? |
| Is a family approach offered postnatally for HIV-positive breastfeeding women and their exposed babies? |
| Are refill clubs offered for pregnant and/or breastfeeding women? |
| **Differentiated ART delivery for other sub-populations** |
| Are services adapted for any key populations? |
| Are services adapted for men? |
| Are services adapted for mobile populations? |
| Are services adapted for people living with disabilities? |
| **Differentiated ART delivery for patients with high viral load** |
| Is there a flagging system to identify who needs a viral load taken? |
| Is there a flagging system to identify who has a VL >1000 copies/ml? |
| Is enhanced adherence implemented? |

### Appendix 5: SOPs for documentation at clinical and refill visits

#### Documentation at the clinical visit

##### Patient care and treatment book documentation at the clinical visit

**OI/ART NUMBER** P P - D D - S S - Y Y Y Y - A - S S S S S

| 1 Visit No. *(Includes visits made at other facility while the patient is away)* | 2 Date of review / Visit Type *(use codes below)* | 3 Weight (kg) | 4 Height for Children below 15 years (cm) | 5 5a. Pregnancy/ Lactating Status *(see codes below)* / 5b. Date of 1st ANC booking and LNMP date | 6 Family Planning Status *(multiple response: use codes below)* | 7 Functional Status *(use codes below)* | 8 WHO *Clinical Stage (1-4) refer to Page 1* | 9 TB Status: 9a. TB Screening *(use codes below)* | 9 TB Status: 9b. TB Investigation Result *(use codes below)* | 10 Opportunistic Infections and Other Problems *(use codes below)* | 11 Cotrimoxazole Prophylaxis: Quantity dispensed (tablets/ ml) | 11 Cotrimoxazole Prophylaxis: % Adherence | 12 IPT Criteria: 12a-IPT Eligibility (Y/N) | 12 IPT Criteria: 12b - IPT Status *(use codes below)* | 13 Reason for Not Starting or Stopping IPT *(use codes below)* | 14 Isoniazid Preventive Therapy: 14a. Quantity dispensed (tablets/ ml) | 14 Isoniazid Preventive Therapy: 14b. % adherence |
|---|---|---|---|---|---|---|---|---|---|---|---|---|---|---|---|---|---|
| | | | | | | | | | | | | X | | | | | X |
| | 1/1/16 F | 68 | | N/A | C | W | 2 | Y | | | 90 | 100% | NE | | | | |
| | | | | | | | | | | | 90 | | | | | | |
| | | | | | | | | | | | 90 | | | | | | |
| | | | | | | | | | | | 90 | | | | | | |

**OI/ART NUMBER** P P - D D - S S - Y Y Y Y - A - S S S S S

| 15 Fluconazole: 15a. Quantity prescribed (tablets/ ml) | 15 Fluconazole: 15b. Quantity dispensed (tablets/ ml) | 16 ARV Status (see codes below) | 17 Adverse Events Status (see codes below) | 18 ARV Reason (see codes below) | 19 ARV combination regimen (see codes below) | 20 ARV medicine: 20a) Quantity prescribed (tablets/ ml) | 20 ARV medicine: 20b) Quantity dispensed (tablets/ ml) | 20 ARV medicine: 20c) % Adherence | 21 CD4 count/ % & Test Date | 22 Viral Load & Test Date | 23 a) ALT (Alanine Transaminase) | 23 b) Creatinine | 23 c) Other diagnostics test | 24 Referred To: (see codes below) | 25 Next Review date | 26 Visit Status (see codes below) | 27 Follow up Status (see codes below) | 28 Name: Of Clinician (Initials) | 28 Name: Of Pharmacy (Dispenser) (Initials) |
|---|---|---|---|---|---|---|---|---|---|---|---|---|---|---|---|---|---|---|---|
| | | | | | | | | X | | | | | | | | X | | | |
| | | 3 | | | IF | 90 | 90 | 100% | | | | | | | 25/3/16 | OT | TX | AA | BB |
| | | | | | | 90 | | | | | | | | | 17/6/16 | | | | |
| | | | | | | 90 | | | | | | | | | 9/9/16 | | | | |
| | | | | | | 90 | | | | | | | | | 2/12/16 | | | | |

#### Patient notebook documentation at the clinical visit

1/1/16

TDF/3TC/EFV (po) 3/12

CTX 960 mg (po) 3/12

Repeat above prescription on 25/3/16
17/6/16
9/9/16

TCB for clinical review and VL 2/12/16

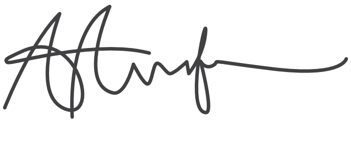

### Documentation at the refill visit

**OI/ART NUMBER** P P - D D - S S - Y Y Y Y - **A** - S S S S S

| 1 | 2 | 3 | 4 | 5 | 6 | 7 | 8 | 9 | | 10 | 11 | | 12 | | 13 | 14 | |
|---|---|---|---|---|---|---|---|---|---|---|---|---|---|---|---|---|---|
| **Visit No.** *(Includes visits made at other facility while the patient is away)* | **Date of review** | **Weight (kg)** | **Height for Children below 15 years (cm)** | **5a. Pregnancy/ Lactating Status** *(see codes below)* | **Family Planning Status** *(multiple response: use codes below)* | **Functional Status** *(use codes below)* | **WHO** *Clinical Stage (1-4) refer to Page 1* | **TB Status** | | **Opportunistic Infections and Other Problems** *(use codes below)* | **Cotrimoxazole Prophylaxis** | | **IPT Criteria** | | **Reason for Not Starting or Stopping IPT** *(use codes below)* | **Isoniazid Preventive Therapy** | |
| | **Visit Type** *(use codes below)* | | | 5b. **Date of 1st ANC booking and LNMP date** | | | | **9a. TB Screening** *(use codes below)* | **9b. TB Investigation Result** *(use codes below)* | | Quantity dispensed (tablets/ ml) | % Adherence | **12a-IPT Eligibility (Y/N)** | **12b - IPT Status** *(use codes below)* | | 14a. Quantity dispensed (tablets/ ml) | 14b. % adherence |
| | | | | | | | | | | | | X | | | | | X |
| | 1/1/16 F | 68 | | N/A | C | W | 2 | Y | | | 90 | 100% | NE | | | | |
| | 25/3/16 F | | | | | | | | | | 90 ✓ | | | | | | |
| | | | | | | | | | | | 90 | | | | | | |
| | | | | | | | | | | | 90 | | | | | | |

**OI/ART NUMBER** P P - D D - S S - Y Y Y Y - **A** - S S S S S

| 15 | | 16 | 17 | 18 | 19 | 20 | | | 21 | 22 | 23 | | | 24 | 25 | 26 | 27 | 28 | |
|---|---|---|---|---|---|---|---|---|---|---|---|---|---|---|---|---|---|---|---|
| **Fluconazole** | | **ARV Status** (see codes below) | **Adverse Events Status** (see codes below) | **ARV Reason** (see codes below) | **ARV combination regimen** (see codes below) | **ARV medicine** | | | **CD4 count/ % & Test Date** | **Viral Load & Test Date** | **a) ALT (Alanine Transaminase)** | **b) Creatinine** | **c) Other diagnostics test** | **Referred To:** (see codes below) | **Next Review date** | **Visit Status** (see codes below) | **Follow up Status** (see codes below) | **Name** | |
| 15a. Quantity prescribed (tablets/ ml) | 15b. Quantity dispensed (tablets/ ml) | | | | | 20a) Quantity prescribed (tablets/ ml) | 20b) Quantity dispensed (tablets/ ml) | 20c) % Adherence | | | | | | | | | | Of Clinician (Initials) | Of Pharmacy (Dispenser) (Initials) |
| | | | | | | | | X | | | | | | | | X | | | |
| | | 3 | | | 1F | 90 | 90 | 100% | | | | | | | 25/3/16 | OT | TX | AA | BB |
| | | | | | | 90 | 90 | | | | | | | | 17/6/16 | OT | TX | | BB |
| | | | | | | 90 | | | | | | | | | 9/9/16 | | | | |
| | | | | | | 90 | | | | | | | | | 2/12/16 | | | | |

### Patient notebook documentation at the refill visit

1/1/16

TDF/3TC/EFV (po) 3/12

CTX 960 mg (po) 3/12

Repeat above prescription on 25/3/16
17/6/16
9/9/16

TCB for clinical review and VL 2/12/16

### Appendix 6: M and E forms for ART clubs and CARGS

#### Facility club register

**Facility "club" group register**

Facility name: | Meeting time:

Club number:

| Club member number | ART number | Full name | Sex | Date of birth | Mobile number | Date ART initiation | Date joined club | Date per-manently left club | Reason left club* |
|---|---|---|---|---|---|---|---|---|---|
| 1 | | | | ../ .. /.... | | ../ .. /.... | ../ .. /.... | ../ .. /.... | |
| 2 | | | | ../ .. /.... | | ../ .. /.... | ../ .. /.... | ../ .. /.... | |
| 3 | | | | ../ .. /.... | | ../ .. /.... | ../ .. /.... | ../ .. /.... | |
| 4 | | | | ../ .. /.... | | ../ .. /.... | ../ .. /.... | ../ .. /.... | |
| 5 | | | | ../ .. /.... | | ../ .. /.... | ../ .. /.... | ../ .. /.... | |
| 6 | | | | ../ .. /.... | | ../ .. /.... | ../ .. /.... | ../ .. /.... | |
| 7 | | | | ../ .. /.... | | ../ .. /.... | ../ .. /.... | ../ .. /.... | |
| 8 | | | | ../ .. /.... | | ../ .. /.... | ../ .. /.... | ../ .. /.... | |
| 9 | | | | ../ .. /.... | | ../ .. /.... | ../ .. /.... | ../ .. /.... | |
| 10 | | | | ../ .. /.... | | ../ .. /.... | ../ .. /.... | ../ .. /.... | |
| 11 | | | | ../ .. /.... | | ../ .. /.... | ../ .. /.... | ../ .. /.... | |
| 12 | | | | ../ .. /.... | | ../ .. /.... | ../ .. /.... | ../ .. /.... | |
| 13 | | | | ../ .. /.... | | ../ .. /.... | ../ .. /.... | ../ .. /.... | |
| 14 | | | | ../ .. /.... | | ../ .. /.... | ../ .. /.... | ../ .. /.... | |
| 15 | | | | ../ .. /.... | | ../ .. /.... | ../ .. /.... | ../ .. /.... | |
| 16 | | | | ../ .. /.... | | ../ .. /.... | ../ .. /.... | ../ .. /.... | |
| 17 | | | | ../ .. /.... | | ../ .. /.... | ../ .. /.... | ../ .. /.... | |
| 18 | | | | ../ .. /.... | | ../ .. /.... | ../ .. /.... | ../ .. /.... | |
| 19 | | | | ../ .. /.... | | ../ .. /.... | ../ .. /.... | ../ .. /.... | |
| 20 | | | | ../ .. /.... | | ../ .. /.... | ../ .. /.... | ../ .. /.... | |

*Reason for leaving club: 1. TFO 2. Moved to another club 3. Permanently returned to clinic care 4. LTFU 5. Died 6. Other

Club appointment dates

| | | | | | | |
|---|---|---|---|---|---|---|
| ../ .. /.... | | ../ .. /.... | | ../ .. /.... | | ../ .. /.... |
| ../ .. /.... | | ../ .. /.... | | ../ .. /.... | | ../ .. /.... |
| ../ .. /.... | | ../ .. /.... | | ../ .. /.... | | ../ .. /.... |
| ../ .. /.... | | ../ .. /.... | | ../ .. /.... | | ../ .. /.... |

### Community ART group register

| CARG group register | | | | | | | | | |
|---|---|---|---|---|---|---|---|---|---|
| Facility name: | | | Group leader name: | | | | Meeting area: | | |
| CARG number: | | | Group leader contact number: | | | | | | |
| CARG member number | ART number | Full name | Sex | DOB | Mobile number | Date ART initiation | Date joined CARG | Date perma-nently left CARG | Reason left CARG* |
| 1 | | | | ../ .. /.... | | ../ .. /.... | ../ .. /.... | ../ .. /.... | |
| 2 | | | | ../ .. /.... | | ../ .. /.... | ../ .. /.... | ../ .. /.... | |
| 3 | | | | ../ .. /.... | | ../ .. /.... | ../ .. /.... | ../ .. /.... | |
| 4 | | | | ../ .. /.... | | ../ .. /.... | ../ .. /.... | ../ .. /.... | |
| 5 | | | | ../ .. /.... | | ../ .. /.... | ../ .. /.... | ../ .. /.... | |
| 6 | | | | ../ .. /.... | | ../ .. /.... | ../ .. /.... | ../ .. /.... | |
| 7 | | | | ../ .. /.... | | ../ .. /.... | ../ .. /.... | ../ .. /.... | |
| 8 | | | | ../ .. /.... | | ../ .. /.... | ../ .. /.... | ../ .. /.... | |
| 9 | | | | ../ .. /.... | | ../ .. /.... | ../ .. /.... | ../ .. /.... | |
| 10 | | | | ../ .. /.... | | ../ .. /.... | ../ .. /.... | ../ .. /.... | |
| 11 | | | | ../ .. /.... | | ../ .. /.... | ../ .. /.... | ../ .. /.... | |
| 12 | | | | ../ .. /.... | | ../ .. /.... | ../ .. /.... | ../ .. /.... | |
| *Reason for leaving CARG: 1. TFO 2. Moved to other CARG 3. Permanently returned to clinic care 4. LTFU 5. Died 6. Other | | | | | | | | | |
| CARG appointment dates | | | | | | | | | |
| ../ .. /.... | | ../ .. /.... | | | ../ .. /.... | | ../ .. /.... | | |
| ../ .. /.... | | ../ .. /.... | | | ../ .. /.... | | ../ .. /.... | | |
| ../ .. /.... | | ../ .. /.... | | | ../ .. /.... | | ../ .. /.... | | |
| ../ .. /.... | | ../ .. /.... | | | ../ .. /.... | | ../ .. /.... | | |

### Community ART group refill form

**Community ART group refill monitoring form**

| | |
|---|---|
| Facility name: | CARG number: |
| Date group meeting before refill ../ .. /.... | Signature of group leader: |
| Date ART prescribed by nurse ../ .. /.... | Signature of nurse: |
| Date ARVs distributed ../ .. /.... | |

| | To be completed By Group Leader | | | | | | To be completed by nurse | | | | To be completed By CAG member | | | |
|---|---|---|---|---|---|---|---|---|---|---|---|---|---|---|
| CARG member number | Full name | Pregnant (P) or on family planning (FP) | TB symptoms* Y/N | Other "alert" problems** | ARV tablets remaining | CTX tablets remaining | ARV regimen prescribed / quantity | CTX quantity prescribed | VL result (CD4 if not available) | Date VL | Full name | Signature of recipient | Date drugs received | Comments (include any reason for temporary clinic follow up) |
| 1 | | | | | | | | | | ../ .. /.... | | | ../ .. /.... | |
| 2 | | | | | | | | | | ../ .. /.... | | | ../ .. /.... | |
| 3 | | | | | | | | | | ../ .. /.... | | | ../ .. /.... | |
| 4 | | | | | | | | | | ../ .. /.... | | | ../ .. /.... | |
| 5 | | | | | | | | | | ../ .. /.... | | | ../ .. /.... | |
| 6 | | | | | | | | | | ../ .. /.... | | | ../ .. /.... | |
| 7 | | | | | | | | | | ../ .. /.... | | | ../ .. /.... | |
| 8 | | | | | | | | | | ../ .. /.... | | | ../ .. /.... | |
| 9 | | | | | | | | | | ../ .. /.... | | | ../ .. /.... | |
| 10 | | | | | | | | | | ../ .. /.... | | | ../ .. /.... | |
| 11 | | | | | | | | | | ../ .. /.... | | | ../ .. /.... | |
| 12 | | | | | | | | | | ../ .. /.... | | | ../ .. /.... | |

*TB symptoms: Ask if the member has a current cough of any duration, is losing weight, has night sweats or has had contact with TB patient in last month

**Alert problems: Ask if the member has any ankle swelling, puffiness of the face, breathlessness, diarrhea for more than 2 weeks, severe headache

### ACKNOWLEDGEMENTS

The Ministry of Health and Child Care acknowledges all stakeholders who participated in the development of the Operational and Service Delivery Manual (OSDM) for the Prevention, care and treatment of HIV in Zimbabwe 2017.

Financial support for the development of the manual was provided by the International AIDS Society (IAS) and FHI360 through the USAID-funded Zimbabwe HIV Care and Treatment (ZHCT) Project, CHAI, The Global Fund for HIV, TB & Malaria, UNICEF and EGPAF.

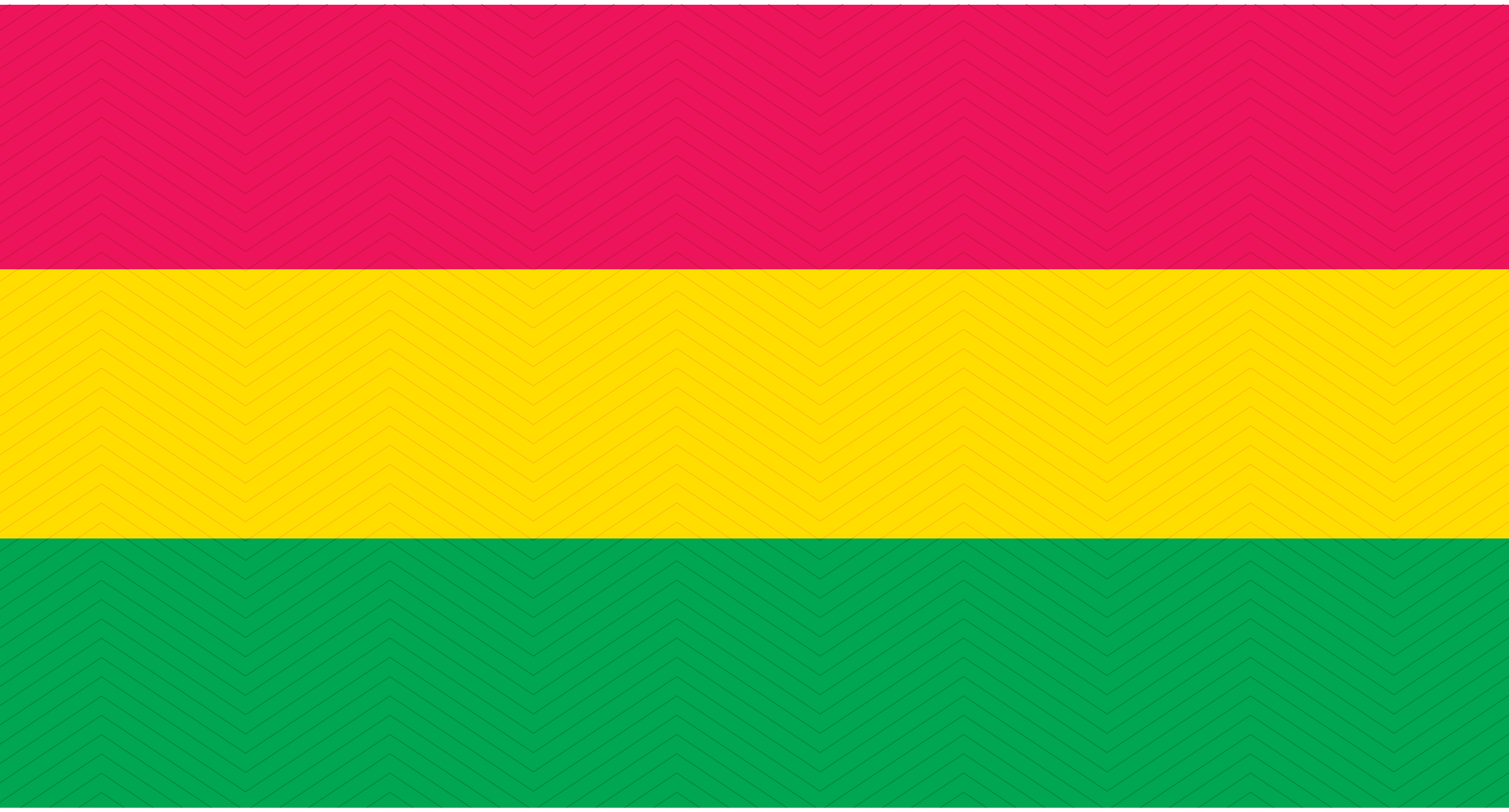
